# Effect of Telemedicine Support for Intraoperative Anaesthesia Care on Postoperative Outcomes: The TECTONICS Randomised Clinical Trial

**DOI:** 10.1101/2024.05.21.24307593

**Authors:** Christopher R King, Bradley A. Fritz, Stephen H. Gregory, Thaddeus P. Budelier, Arbi Ben Abdallah, Alex Kronzer, Daniel L. Helsten, Brian Torres, Sherry L. McKinnon, Sandhya Tripathi, Mohamed Abdelhack, Shreya Goswami, Arianna Montes de Oca, Divya Mehta, Miguel A. Valdez, Evangelos Karanikolas, Omokhaye Higo, Paul Kerby, Bernadette Henrichs, Troy S. Wildes, Mary C. Politi, Joanna Abraham, Michael S. Avidan, Thomas Kannampallil, the ACTFAST collaborator group

## Abstract

**Background:** Novel applications of telemedicine can improve care quality and patient outcomes. Telemedicine for intraoperative decision support has not been rigorously studied.

**Methods:** This single centre randomised clinical trial (RCT, clinicaltrials.gov NCT03923699) of unselected adult surgical patients was conducted between 2019-07-01 and 2023-01-31. Patients received usual-care or decision support from a telemedicine service, the Anesthesiology Control Tower (ACT). The ACT provided real-time recommendations to intraoperative anaesthesia clinicians based on case reviews and physiologic alerts. ORs were randomised 1:1. Co-primary outcomes of 30-day all-cause mortality, respiratory failure, acute kidney injury (AKI), and delirium in the Intensive Care Unit (ICU) were analysed as intention-to-treat.

**Results:** The trial completed with 71927 surgeries (35302 ACT; 36625 usual care). The ACT performed 11812 case reviews and communicated alerts regarding 2044 intervention-group patients. There was no significant effect of the ACT vs. usual care on 30-day mortality [630/35302 (1.8%) vs 649/36625 (1.8%), RR 1.01 (95% CI 0.87 to 1.16), p=0.98], respiratory failure [1071/33996 (3.2%) vs 1130/35236 (3.2%), RR 0.98 (95% CI 0.88 to 1.09), p=0.98], AKI [2316/33251 (7.0%) vs 2432/34441 (7.1%), RR 0.99 (95% CI 0.92 to 1.06), p=0.98] or delirium [1264/3873 (32.6%) vs 1298/4044 (32.1%), RR 1.02 (95% CI 0.94 to 1.10), p=0.98]. There were no significant differences in secondary outcomes or sensitivity analyses.

**Conclusions:** In this large RCT of intraoperative telemedicine decision support using real-time alerts and case reviews, we found no significant differences in postoperative outcomes. Large-scale intraoperative telemedicine is feasible, and we suggest avenues where it may be more impactful.

## Introduction

Tele-critical care (TCC), the use of remote monitoring and telemedicine to augment intensive care unit (ICU) services, ^1^ operates in most academic hospitals^2^ and in nearly 20% of hospitals with ICU beds.^3^ Although there are multiple TCC models, ^4^ contemporary TCC supports bedside teams through a combination of expert consultations, real-time review of emerging problems, continuous remote monitoring, encouragement of best practices, and facilitating communication. ^2^ TCC support aims to improve care quality and patient outcomes by reducing and overcoming distractions to bedside clinicians. ^2, 5^ Although effect size estimates from observational and quasi-experimental TCC studies have been heterogeneous,^6^ TCC has repeatedly been associated with improved patient outcomes. ^1, 2^ However, recent cluster-randomized RCTs have found no effect on patient outcomes. ^5, 7^

Although seemingly a natural extension of the adoption of TCC, there is limited research evaluating remote monitoring or telemedicine models for intraoperative care. Existing case reports focus on intraoperative one-on-one video calls to support low-resource or remote locations. ^8–13^ This is surprising because critical care and anaesthesia care are closely related fields, with similar physiologic changes and monitoring technology. Like critical care settings, anaesthesia clinicians experience high cognitive load and distractions from complex, high-frequency data combined with procedures and documentation. Decision support from telemedicine may therefore have a similar beneficial role. ^14^

Therefore, we hypothesized that remote monitoring and support via a telemedicine centre for anaesthesia clinicians similar to current tele-critical care delivery models would improve surgical patient outcomes. Towards this end, we developed the “Anesthesiology Control Tower” (ACT), ^15–17^ a telemedicine support service for anaesthesia clinicians in the OR. Key activities of the ACT included (a) reviewing the anaesthetic plans for higher-risk cases and communicating recommendations to the OR anaesthesia clinicians, (b) continuous monitoring of real-time data and alerts to assess patient deterioration (c) encouraging adherence to institutional protocols (e.g., hyperglycaemia management), and (d) assisting in crisis management and coordinating out-of-OR resources. A pilot and feasibility trial of the ACT (ACTFAST-3) showed the feasibility of using a real-time telemedicine system for intraoperative decision support and collaborative decision-making.^16^ Additional design and process changes were made based on studies assessing the workflow of ORs and ACT ^15^ and patient input. ^18^

The objective of this randomised clinical trial (RCT), Telemedicine Control Tower for the OR: Navigating Information, Care, and Safety (TECTONICS), was to evaluate the impact of remote monitoring and decision support by the ACT on postoperative patient outcomes.

## Methods

### Study design and ethics

TECTONICS was a pragmatic, randomised parallel, single-centre, superiority trial conducted at Barnes-Jewish Hospital (BJH) and Washington University School of Medicine in St. Louis, MO, USA. The institutional review board at Washington University approved the study with a waiver of informed consent. TECTONICS’s protocol and statistical analysis plan were registered at clinicaltrials.gov (NCT03923699). A data safety monitoring board (DSMB) met quarterly but conducted no interim efficacy analyses. Supplemental methods 13 contains a PRECIS-2 analysis of trial pragmatism.

### Setting and population

BJH is an adult urban tertiary centre. During the study (July 1, 2019 to January 30, 2023), 59 ORs were included, with at most 54 concurrent rooms. These excluded non-surgical procedure suites and obstetrics. The site used a “medical direction” anaesthesia care team model (supplemental methods 8). We enrolled all adult patients (>=18 years) with surgery starting between 06:15 and 16:00 weekdays with anaesthesia services, including emergency cases. The ACT was closed, and patients excluded, on days with clinical staff shortages (supplemental methods 6). Patients were followed for 30 days to ascertain outcomes.

### Intervention: Anesthesiology Control Tower (ACT)

Patients in the intervention group received ACT-enhanced care; we have previously described the development, preliminary evaluation and functioning of the ACT. ^15–17^ The ACT was a remote monitoring suite with real-time data feeds from the Electronic Health Record (EHR) and the AlertWatch:OR (Ann Arbor, Michigan) informatics platform. ACT screens displayed a customized AlertWatch:OR interface with a dashboard summarizing all active surgeries. The dashboard contained an overall patient complexity measure, current data-driven alerts, and links to patient-specific views of current data and trends (Supplemental Figure 1). To assist with prioritizing cases for review and quantifying patient risks for ACT-OR communication, a web application displayed machine-learning predictions of individual patient risk of several major adverse events (Supplemental Figure 2). ^19–22^ The implementation and validation of this web application is described elsewhere. ^23^ ACT staff also used the Epic EHR to access patient information.

The supplement (methods 10 and 14) contains the ACT alert criteria and manual of procedures, including the selection of cases and alerts to review, how the ACT analysed and communicated to the OR clinicians, and roles and responsibilities of ACT staff members. The minimal staffing for the ACT to operate was a research coordinator and an attending anaesthesiologist; however, it was possible to have up to 5 staff members in the ACT simultaneously (research coordinator, attending, resident, certified registered nurse anaesthetist, and a student nurse anaesthetist). The ACT had two main activities: (1) pre-emptively reviewing anaesthetic plans in active cases (2) addressing alerts generated by real-time data in the Alertwatch:OR dashboard. The selection of which cases to pre-emptively review was at the anaesthesiologist’s discretion, with the patient complexity and complication-risk tools mentioned above available to complement their assessment of procedural complexity. The ACT reviewed anaesthesia plans in both intervention and usual care group cases, however, a summary of risk-mitigation recommendations and applicable protocols was sent to the intraoperative anaesthesia clinicians only in the intervention group. Similarly, the ACT staff chose which alerts should be silenced and which should be communicated to the intraoperative clinicians. ACT rationales for contacting ORs were logged by the research coordinator. To improve consistency, the ACT research coordinators were trained to assist clinicians in following the guidelines for case selection and alert response. The ACT contacted clinicians via Epic’s Secure Chat messaging or hospital-issued phones.

Staffing and monitoring were unchanged in the usual care group. ACT staff reviewed anaesthesia plans and alerts in usual-care patients, but the results were not communicated to intraoperative clinicians. ACT staff contacted intraoperative clinicians in usual-care ORs only in situations with potential imminent danger to patients, such as the failure to deliver anaesthetic agents. Intraoperative clinicians in either group were allowed to contact the ACT for assistance if they felt it was necessary for patient care; however, in practice this was uncommon.

### Randomisation and Blinding

Each day, operating rooms (and all patients nested in each OR) were 1:1 randomised to intervention or usual care (cluster randomisation by operating room and day). Randomisation was calculated and displayed in the ACT dashboard at midnight without considering the number of cases scheduled in each room (including zero). Therefore, the realized randomisation ratio of patients varied. Only the first surgery for a patient in a 30-day window was analysed; cases 30-days after an index surgery were eligible for analysis. Outcome assessors and patients were blinded to assignments. Clinicians learned of their assignment only if the ACT contacted them; i.e., if contacted they were aware that they were in the intervention group, but if uncontacted they were unsure whether they were assigned to usual-care or intervention with no issues warranting ACT communication. Because group assignments were fixed for a day, this information carried over to subsequent cases.

### Patient Outcomes

Four co-primary outcomes were ascertained: 30-day all-cause mortality, postoperative respiratory failure, postoperative acute kidney injury (AKI), and postoperative delirium. Delirium was routinely measured and recorded only in patients admitted to the surgical and cardiothoracic ICUs. These outcomes were chosen to represent major patient-oriented complications. Secondary outcomes were a composite (sum of co-primary outcomes), hyperglycaemia, hypotension, normothermia, low peak airway pressures, avoidance of gaps in volatile anaesthetic delivery, and efficient use of volatile anaesthetics, which are operationalized in the supplement (methods 1). Alternative specifications are also defined in the supplement. All outcomes were incident. Definitions, missing data handling, and EHR measurements used for primary and secondary outcomes are contained in the Supplement (methods 2). Data was extracted from the EHR in two waves (2019-2020, 2021-2023).

### Sample size calculation

Sample size was calculated per the protocol with a planned enrolment of 40,000 over 4 years. ^24^ As a low-risk health system intervention, there was no clear minimally clinically important difference. Due to a larger than expected number of eligible patients, during the first year of the study, the principal investigator and DSMB agreed to expand the study population to 80,000 to allow detection of smaller but potentially meaningful intervention effects.

### Statistical analysis

Relative risks for all primary outcomes were calculated with a Poisson generalized estimating equation (GEE) model clustering on OR and day (the unit of randomisation) and HC1 adjustment to standard errors. ^25^ Risk differences were calculated with a linear-link GEE model using the same clustering. Bonferroni adjusted 95% and 99.5% confidence intervals were calculated. Two-sided p-values were adjusted for multiple testing ^26^ with an alpha of 0.005. 27 Secondary outcomes were analysed in the same manner; a linear link GEE model was used for non-binary outcomes. All analyses were conducted intention-to-treat (ignoring actual communication between the ACT and OR) among all patients with ascertainable outcomes. Planned subgroup and sensitivity analyses are described in the supplement (methods 4 and 9). A hypothesis-generating per-protocol analysis comparing intervention group patients with case reviews and alert recommendations communicated to the OR to usual-care patients matched on patient and case characteristics is described in the supplement (methods 7). Analysis used R version 4.4.0; a docker file with package versions and analysis code is available at https://github.com/cryanking/tectonics_deident.

### Intervention monitoring

ACT staff logged case reviews, alert interpretation, and OR communication (OR interventions) in the AlertWatch:OR web interface using discrete fields and free-text comments. Due to a database malfunction, ACT logs of case reviews, ACT to OR communication, and alert interpretations were lost for a block of 67/833 study days; we report contacts and alerts from discrete data on days with intact logs. A previous manuscript ^15^ describes workflow observations, recommendations, and adherence during a portion of the trial using simultaneous observation of ORs and the ACT. A random sample of contacted cases were manually reviewed for recommendation adherence. We did not characterize ACT recommendations and adherence for all case reviews as there are many possible anaesthesia plan modifications.

## Results

Figure 1 displays the included population. The trial completed its targeted enrolment with 71927 surgeries included in the primary analysis, with 35302 cases (17790 clusters) allocated to intervention and 36625 cases (18211 clusters) allocated to usual care. There were minimal differences in comorbidities, functional status, surgery class, or other characteristics (Table 1). The mean patient age was 57 ranging from 18 to 101; patients were 49% male and 51% female. Patient races were white (70%) or Black (20%), with 9% unrecorded or belonging to multiple groups. A wide variety of surgery types were included; orthopaedics was the most common high-level grouping (18%).

**Figure 1:**
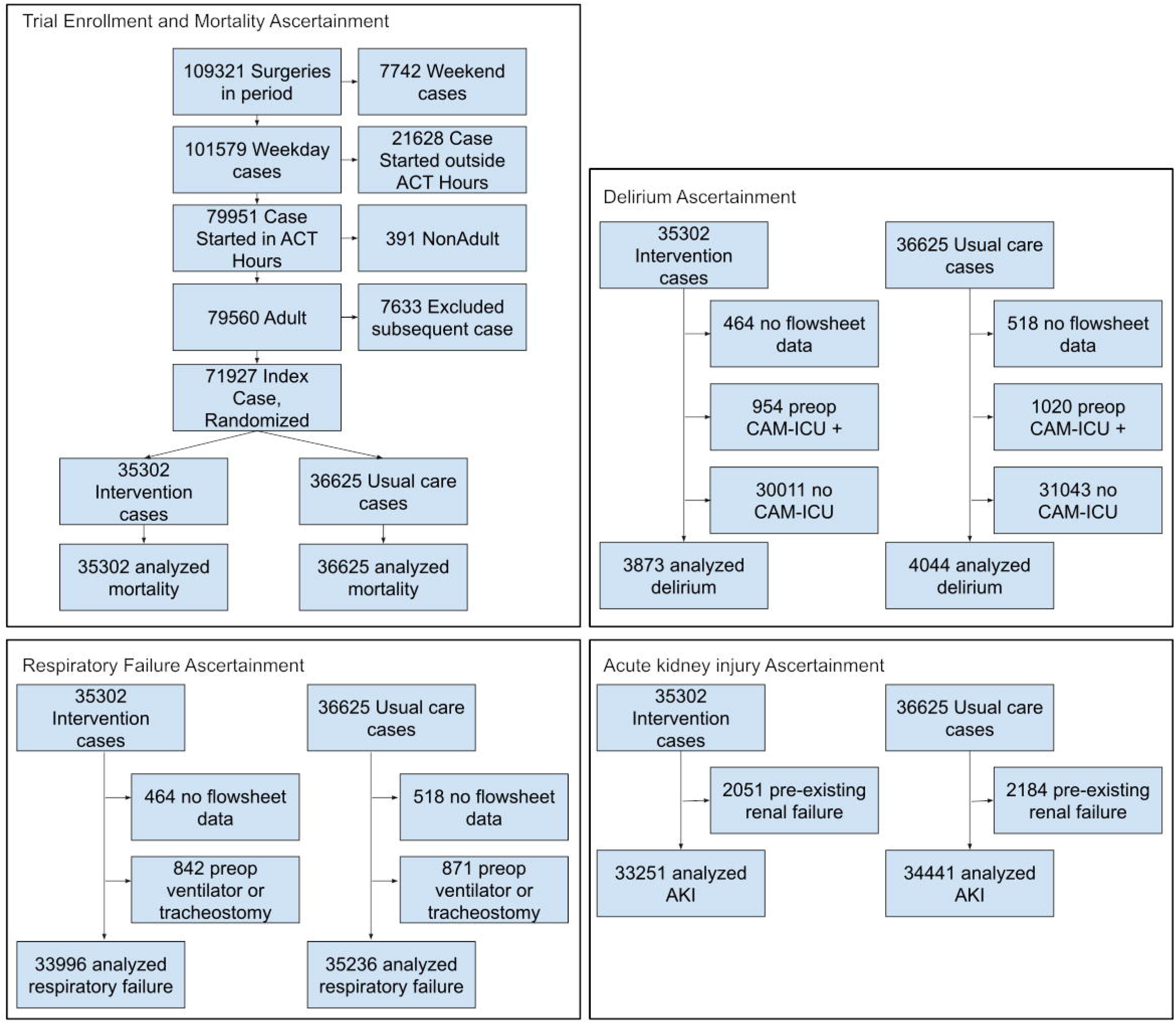
Patient flow and exclusion criteria. ACT hours = 0615 to 1600. AKI =acute kidney injury. CAM-ICU = confusion assessment method for the ICU. “no flowsheet data” = no data returned for specified medical record number.

**Figure 2:**
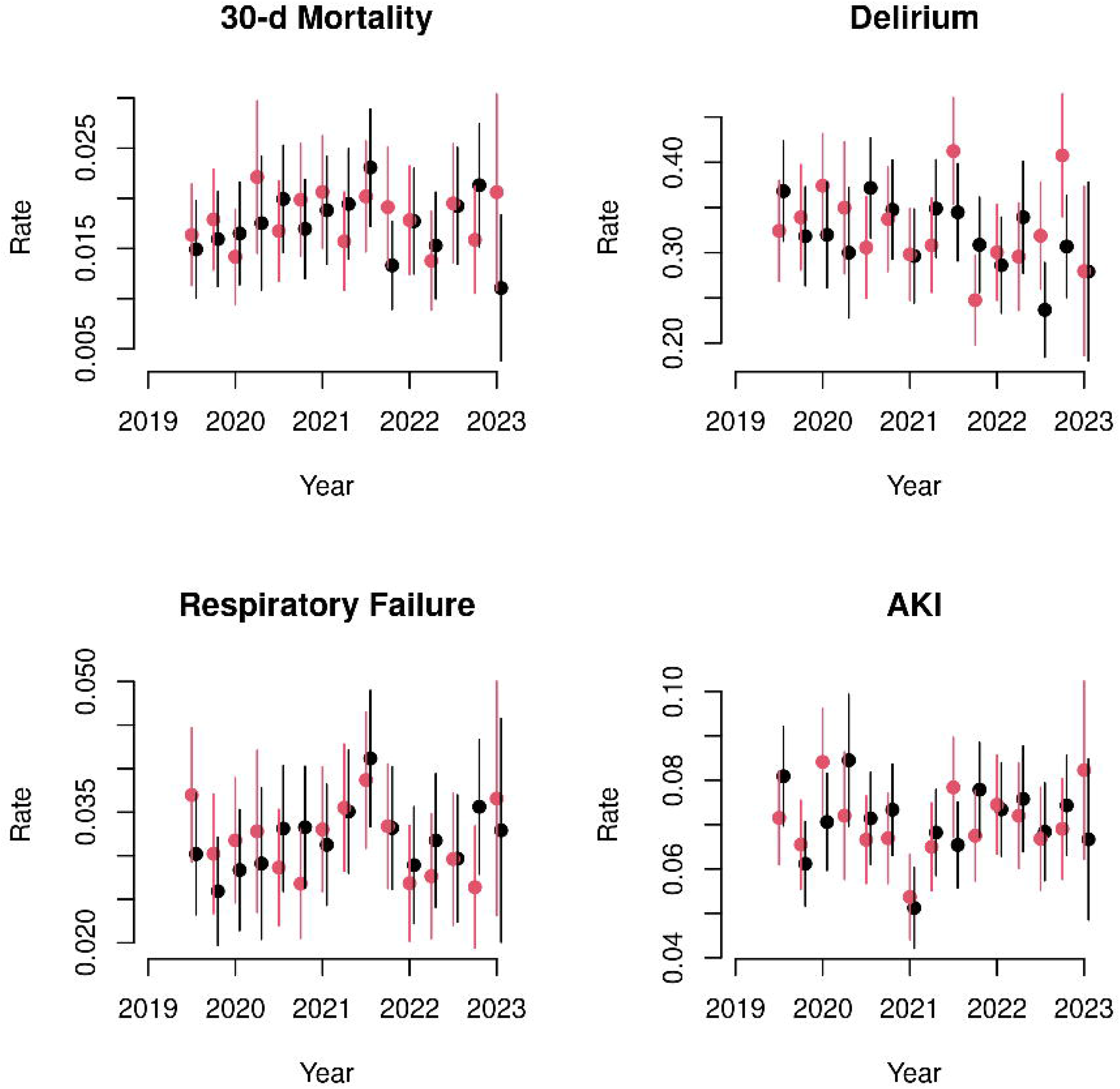
Primary outcome rates by treatment group over time. Bars = pointwise 95% confidence intervals from a linear GEE model using the same clustering as the primary analysis and HC1 adjustment. Data aggregated over 3-month intervals. Red dots = ACT, black = usual care.

**Table 1:** Baseline characteristics of patients. Age = mean (range), otherwise mean (SD) or k/N (%), “Standardized diff” = Cohen’s D for binary, Phi for categorical, standardized mean difference for continuous. Groups with less than 100 patients not shown (merged into “Other”). Atrial Fib = atrial fibrillation, CAD = coronary artery disease, CKD = chronic kidney disease, ESRD = end stage renal disease, COPD = chronic obstructive pulmonary disease, VTE = venous thromboembolic disease, OSA = obstructive sleep apnoea, HTN = hypertension, METs = metabolic equivalents, ASA-PS = American Society of Anesthesiologists physical status, GI = gastroenterology. Patients with more than one included surgery appear multiple times.

| variable | Intervention, n=35302 | Usual care, n=36625 | Standardized diff |
| --- | --- | --- | --- |
| Age | 57.2 (18-101) | 57.4 (18-101) | 0.01 (0.00 , 0.03) |
| Atrial Fib | 2897/32377 (8.9%) | 3116/33527 (9.3%) | 0.00 (0.00 , 0.01) |
| Anaemia | 9034/32377 (27.9%) | 9531/33527 (28.4%) | 0.00 (0.00 , 0.01) |
| Asthma | 3692/32377 (11.4%) | 3728/33527 (11.1%) | 0.00 (0.00 , 0.01) |
| CAD | 4248/32377 (13.1%) | 4328/33527 (12.9%) | 0.00 (0.00 , 0.01) |
| Cancer | 3690/32377 (11.4%) | 3793/33527 (11.3%) | 0.00 (0.00 , 0.01) |
| CKD | 4381/32377 (13.5%) | 4571/33527 (13.6%) | 0.00 (0.00 , 0.01) |
| ESRD | 1476/32377 (4.6%) | 1596/33527 (4.8%) | 0.00 (0.00 , 0.01) |
| COPD | 3407/32377 (10.5%) | 3519/33527 (10.5%) | 0.00 (0.00 , 0.01) |
| Dementia | 990/32377 (3.1%) | 1068/33527 (3.2%) | 0.00 (0.00 , 0.01) |
| Diabetes | 7675/32377 (23.7%) | 8054/33527 (24%) | 0.00 (0.00 , 0.01) |
| Stroke | 1907/32377 (5.9%) | 1946/33527 (5.8%) | 0.00 (0.00 , 0.01) |
| VTE | 3227/32377 (10%) | 3431/33527 (10.2%) | 0.00 (0.00 , 0.01) |
| OSA | 5787/32377 (17.9%) | 5991/33527 (17.9%) | 0.00 (0.00 , 0.01) |
| HTN | 16697/32377 (51.6%) | 17070/33527 (50.9%) | 0.01 (0.00 , 0.01) |
| Race |  |  | 0.00 (0.00 , 0.01) |
| American Indian or Alaska Native | 93/35302 (0.3%) | 106/36625 (0.3%) |  |
| Asian | 409/35302 (1.2%) | 399/36625 (1.1%) |  |
| Black or African American | 6849/35302 (19.4%) | 7158/36625 (19.5%) |  |
| Other Pacific Islander | 50/35302 (0.1%) | 65/36625 (0.2%) |  |
| Unknown or Other | 3234/35302 (9.2%) | 3368/36625 (9.2%) |  |
| White | 24667/35302 (69.9%) | 25529/36625 (69.7%) |  |
| Sex |  |  | 0.00 (0.00 , 0.01) |
| Female | 18047/35302 (51.1%) | 18589/36625 (50.8%) |  |
| Male | 17252/35302 (48.9%) | 18033/36625 (49.2%) |  |
| Unknown or Other | 3/35302 (0%) | 3/36625 (0%) |  |
| Functional Capacity |  |  | 0.01 (0.00 , 0.02) |
| <4 METs | 9763/35302 (27.7%) | 10274/36625 (28.1%) |  |
| 4-6 METs | 16417/35302 (46.5%) | 16672/36625 (45.5%) |  |
| 6-10 METs | 779/35302 (2.2%) | 748/36625 (2%) |  |
| >10 METs | 137/35302 (0.4%) | 128/36625 (0.3%) |  |
| Unknown or Unable to Assess | 8206/35302 (23.2%) | 8803/36625 (24%) |  |
| ASA-PS |  |  | 0.01 (0.00 , 0.02) |
| 1 | 1358/35302 (3.8%) | 1355/36625 (3.7%) |  |
| 2 | 11297/35302 (32%) | 11584/36625 (31.6%) |  |
| 3 | 15147/35302 (42.9%) | 15729/36625 (42.9%) |  |
| 4 | 3047/35302 (8.6%) | 3385/36625 (9.2%) |  |
| 5 | 93/35302 (0.3%) | 99/36625 (0.3%) |  |
| Unknown or Other | 4360/35302 (12.4%) | 4473/36625 (12.2%) |  |
| Surgical Service |  |  | 0.02 (0.00 , 0.03) |
| Cardiothoracic | 3377/35302 (9.6%) | 3569/36625 (9.7%) |  |
| Colorectal | 1062/35302 (3%) | 1202/36625 (3.3%) |  |
| General and Trauma | 4546/35302 (12.9%) | 4490/36625 (12.3%) |  |
| Gynaecology | 4106/35302 (11.6%) | 4153/36625 (11.3%) |  |
| Misc procedures, GI, Ophthalmology | 2183/35302 (6.2%) | 2615/36625 (7.1%) |  |
| Neurosurgery | 2624/35302 (7.4%) | 2716/36625 (7.4%) |  |
| Orthopaedics | 6377/35302 (18.1%) | 6392/36625 (17.5%) |  |
| Otolaryngology | 2964/35302 (8.4%) | 3021/36625 (8.2%) |  |
| Plastics | 784/35302 (2.2%) | 788/36625 (2.2%) |  |
| Transplant | 1154/35302 (3.3%) | 1224/36625 (3.3%) |  |
| Urology | 3736/35302 (10.6%) | 3987/36625 (10.9%) |  |
| Vascular | 2389/35302 (6.8%) | 2468/36625 (6.7%) |  |

Supplementary Figures 3, 4, and 5 show enrolment rates and ascertainment rates of primary and secondary outcomes over time. Aside from a period of reduced surgical volume and limited staff availability during the COVID-19 pandemic (Supplementary Figure 6), recruitment and outcome ascertainment were consistent and not differential by study group. The low rate of ascertainment of incident delirium was anticipated because routine CAM-ICU measurement was performed only in some ICUs. Intervention and usual care groups were well separated with respect to ACT-OR contact. ACT clinicians reviewed anaesthesia plans on 11812/34576 (34%) intervention cases and 1845/35919 (5%) usual care cases. 9246 (27%) and 81 (<1%) case reviews in intervention and usual care were sent with recommendations to OR clinicians. The ACT contacted 2044/31490 (6.5%) of intervention cases and 238/32671 (0.7%) of usual care cases regarding alerts. Denominators for ACT actions are smaller than the total number of participants because of a database malfunction affecting several months (Supplementary Methods 5). Supplementary Figures 7 and 8 show ACT activity by month. Supplementary Table 1 shows patient characteristics of reviewed vs unreviewed intervention group cases. Supplementary Table 2 shows the number of case reviews responded to by OR clinicians.

**Table 2:** Intention-to-treat effect estimates of the intervention. Missing due to either outcome data not present, or patient not eligible, such as outcome present at baseline. Usual care and intervention columns [yes/number measured (%)] or [mean (SD)] for continuous outcomes. GEE coefficients from clustered Poisson model (binary outcome) or linear model (continuous outcomes) rounded to 2 decimal places; -0.00 is a negative number rounding to 0 to two decimal places. 95% CI Bonferroni corrected for multiple testing. P-value by permutation (5000 permutations) corrected for multiple testing. 24 Identical adjusted p-values due to step-down procedure; unadjusted results in Supplemental Table 12. AKI = acute kidney injury, PIP = peak inspiratory pressure. Secondary outcomes defined in the supplement.

| Outcome | Intervention | Usual Care | GEE coef (95% CI) | p value |
| --- | --- | --- | --- | --- |
| Delirium | 1264/3873 (32.6) | 1298/4044 (32.1) | 1.02 (0.94, 1.10) | 0.98 |
| Respiratory Failure | 1071/33996 (3.2) | 1130/35236 (3.2) | 0.98 (0.88, 1.09) | 0.98 |
| AKI | 2316/33251 (7.0) | 2432/34441 (7.1) | 0.99 (0.92, 1.06) | 0.98 |
| 30-day mortality | 630/35302 (1.8) | 649/36625 (1.8) | 1.01 (0.87, 1.16) | 0.98 |
| *Secondary Outcomes* |  |  |  |  |
| On time Antibiotic redosing | 5140/5402 (95.1) | 5255/5509 (95.4) | 1.00 (0.99, 1.01) | 0.92 |
| Normothermia | 24482/31147 (78.6) | 24906/31877 (78.1) | 1.01 (0.99, 1.02) | 0.70 |
| Fraction time hypotensive | 0.03 (0.08) | 0.04 (0.08) | -0.00 (-0.00, 0.00) | 0.70 |
| Fraction time acceptable PIP | 0.92 (0.22) | 0.92 (0.21) | -0.00 (-0.01, 0.00) | 0.94 |
| Hyperglycaemia | 1669/28469 (5.9) | 1616/29599 (5.5) | 1.07 (0.98, 1.18) | 0.24 |
| Anaesthetic delivery without gaps | 26967/27278 (98.9) | 27688/27976 (99) | 1.00 (1.00, 1.00) | 0.70 |
| Efficient gas flow > 90% | 17968/24018 (74.8) | 18546/24750 (74.9) | 1.00 (0.98, 1.01) | 0.94 |

For the co-primary outcomes, there was no significant difference in 30-day mortality [ACT 630/35302 (1.8%) vs usual care 649/36625 (1.8%), risk difference 0.0% (95% CI -0.2, 0.3), RR 1.01 (95% CI 0.87 to 1.16), p=0.98], respiratory failure [1071/33996 (3.2%) vs 1130/35236 (3.2%), risk difference -0.1% (95% CI -0.4% to 0.3%), RR 0.98 (95% CI 0.88 to 1.09), p=0.98], AKI [2316/33251 (7.0%) vs 2432/34441 (7.1%), risk difference -0.1% (-0.6% to 0.4%), RR 0.99 (95% CI 0.92 to 1.06), p=0.98], or delirium [1264/3873 (32.6%) vs 1298/4044 (32.1%), risk difference 0.5% (-2.0% to 3.2%), RR 1.02 (95% CI 0.94 to 1.10), p=0.98]. Table 2 shows event rates and effect estimates for primary and secondary outcomes. Figure 2 shows time trends in unadjusted quarterly event rates in usual care and intervention groups.

After multiple testing correction, there were no significant differences by intervention group for any secondary outcome (Table 2). The results were consistent across alternative specifications, subgroup analysis, and sensitivity analyses (Supplement Tables 3-13).

**Table 3:** Reasons Why ACT Clinicians Contacted Intraoperative Clinicians. N = 2084 patients with contact, excluding pre-emptive case reviews.

| Reason for Contact | Intervention Group (N=1932) | Usual Care Group (N=152) | Action Recommended | Intervention Group | Usual Care Group |
| --- | --- | --- | --- | --- | --- |
| Glucose | 852 | 16 | Measure glucose value when feasible if not already done | 625/852 (73%) | 3/16 (19%) |
|  |  |  | Measure glucose value urgently if not already done | 100/852 (12%) | 6/16 (38%) |
|  |  |  | Initiate insulin infusion | 86/852 (10%) | 4/16 (25%) |
|  |  |  | Obtain new glucose value | 74/852 (9%) | 2/16 (12%) |
|  |  |  | Administer short-acting insulin bolus IV | 6/852 (1%) | 3/16 (19%) |
| Temperature | 502 | 7 | Initiate temperature documentation for case, including applying temperature monitor if not in place | 197/502 (39%) | 0/7 (0%) |
|  |  |  | Increase warming therapies | 176/502 (35%) | 1/7 (14%) |
|  |  |  | Maximize warming therapies (e.g., warm room) | 124/502 (25%) | 1/7 (14%) |
|  |  |  | Assess monitoring for accuracy and/or consider altering monitoring | 36/502 (7%) | 0/7 (0%) |
|  |  |  | Decrease warming therapies or introduce cooling therapies | 29/502 (6%) | 1/7 (14%) |
|  |  |  | 'Measure/document more recent patient temperature ' | 23/502 (5%) | 0/7 (0%) |
| Antibiotic | 254 | 1 | Administer (and document) antibiotics if not already done | 128/254 (50%) | 0/1 (0%) |
|  |  |  | Consider administering (and documenting) repeat dose of antibiotics if not already done | 84/254 (33%) | 1/1 (100%) |
| Hypotension | 103 | 49 | Administer additional volume | 55/103 (53%) | 14/49 (29%) |
|  |  |  | Administer vasopressor | 46/103 (45%) | 10/49 (20%) |
|  |  |  | Increase vasopressor dosing | 38/103 (37%) | 4/49 (8%) |
|  |  |  | Decrease anaesthetic dose | 17/103 (17%) | 2/49 (4%) |
|  |  |  | Administer additional blood products | 14/103 (14%) | 4/49 (8%) |
|  |  |  | Use different vasopressor | 10/103 (10%) | 5/49 (10%) |
| Volatile Anaesthetic Concentration | 64 | 16 | Decrease anaesthetic dose | 36/64 (56%) | 1/16 (6%) |
|  |  |  | Increase volatile anaesthetic dose or administer additional IV anaesthetics | 11/64 (17%) | 5/16 (31%) |
|  |  |  | Consider EEG monitoring | 7/64 (11%) | 2/16 (12%) |
| Train of Four | 69 | 0 | Measure (and document) TOF if not already done | 46/69 (67%) | 0 |
|  |  |  | Document pre-reversal TOF if not already done or consider post-reversal tetany assessment | 22/69 (32%) | 0 |
| Blood Pressure Monitoring | 38 | 15 | Obtain BP if not already done or attempting | 36/38 (95%) | 15/15 (100%) |
| Fresh Gas Flow | 45 | 0 | Reduce fresh gas flow if possible to conserve anaesthetic agent | 44/45 (98%) | 0 |
|  |  |  | Assess for possible cause of high gas flow requirement (e.g., leak, inspired CO2) | 6/45 (13%) | 0 |
| Potassium | 36 | 5 | Remeasure potassium | 18/36 (50%) | 2/5 (40%) |
|  |  |  | Treat with potassium and remeasure | 11/36 (31%) | 2/5 (40%) |
|  |  |  | View EKG | 9/36 (25%) | 1/5 (20%) |
| Oxygenation | 14 | 14 | Re-assess ETT position | 6/14 (43%) | 3/14 (21%) |
|  |  |  | Assess pulse oximeter waveform and replace sensor if necessary | 5/14 (36%) | 3/14 (21%) |
|  |  |  | Increase FiO2 | 4/14 (29%) | 3/14 (21%) |
|  |  |  | Increase PEEP | 3/14 (21%) | 2/14 (14%) |
|  |  |  | Assess for and/or treat bronchospasm | 3/14 (21%) | 2/14 (14%) |
| Tidal Volume | 22 | 1 | Adjust tidal volume | 19/22 (86%) | 1/1 (100%) |
| Tachycardia | 20 | 2 | Administer additional analgesics | 8/20 (40%) | 0/2 (0%) |
|  |  |  | 'Administer IV fluid in setting of hypovolemia ' | 8/20 (40%) | 0/2 (0%) |
|  |  |  | Administer beta-blocker | 6/20 (30%) | 0/2 (0%) |
|  |  |  | Increase anaesthetic dose | 5/20 (25%) | 0/2 (0%) |
| Lung Compliance | 12 | 2 | Re-assess circuit and ETT position and patency | 5/12 (42%) | 0/2 (0%) |
|  |  |  | Reduce tidal volume | 3/12 (25%) | 0/2 (0%) |
|  |  |  | Assess for and treat bronchospasm | 3/12 (25%) | 0/2 (0%) |
| Other Automated Alert | 56 | 11 |  |  |  |
| ANY AUTOMATED ALERT | 1883 | 130 |  |  |  |
| Issues other than Automated Alerts | 61 | 22 | New issue identified by ACT without an automated alert | 49/61 (80%) | 3/22 (14%) |
|  |  |  | OR clinicians contacted the ACT to request assistance | 12/61 (20%) | 19/22 (86%) |

Table 3 displays the frequency and reason for ACT interventions other than case reviews. Supplement Table 14 characterizes adherence to ACT recommendations, which were actionable in 69% (95% CI 60% to 78%) of cases and followed in 51% (95% CI 41% to 61%) of cases.

## Discussion

This large (N=71927) randomised trial tested the hypothesis that compared to usual care, remote monitoring and telemedicine-based support from a novel Anesthesiology Control Tower using real-time alerts, machine learning risk identification, and in-depth case reviews for intraoperative anaesthesia clinicians would improve 4 key postoperative patient outcomes: 30-day mortality, respiratory failure, AKI, and delirium. We found no significant effects of randomisation to ACT support on any of the primary or secondary outcomes, nor did we find discordant results in subgroup or sensitivity analyses. Because mortality and respiratory failure are rare (Table 2) and multiple testing decreases certainty, confidence intervals on the effect sizes included some clinically meaningful values. However, for the more common outcomes (AKI and delirium) and secondary outcomes, the results excluded meaningful effect sizes (Table 2).

### Comparison to prior trials

The only directly comparable study for synthesis is our pilot trial, ACTFAST-3, ^16^ which had similar findings regarding effects of the ACT on patient outcomes and process measures. Notably, ACTFAST-3 was smaller and designed to detect changes in process measures (temperature and glycaemic control). TECTONICS is also distinguished from ACTFAST-3 by having a stable intervention versus adaptation during implementation and the deployment of a machine learning risk-estimation tool. Our results should also be compared to Kheterpal et al., who quasi-experimentally studied the impact of the Alertwatch:OR application for intraoperative clinicians on anaesthesia process measures, finding that time with hypotension, inappropriately large tidal volumes, and costs improved. ^14^ Our results are therefore surprising given the most serious alerts were communicated to the intraoperative clinician, and that very few clinicians at the study site otherwise used Alertwatch:OR.

Our intervention is an adaptation of widely adopted strategies in tele-critical care. 4 Our results are therefore surprising given the positive association of TCC with ICU patient outcomes. 6 The study setting, an urban academic centre, also mirrors the settings in which TCC was most associated with improved patient outcomes. 6 However, there are no individual-level RCTs of TCC’s effect on patient outcomes, and a variety of biases may have led to over-estimation of effects in the before-after studies which dominated tele-critical care’s evidence base during the design and implementation of TECTONICS. A recent cluster-randomised trial of TCC with multidisciplinary rounds, quality feedback meetings, and expert-guided protocols found no effect on patient outcomes, 7 and an earlier RCT focused on recognized quality strategies and expert consultations found effects on protocol adherence but not patient outcomes. 5 Importantly, because of the (nearly) patient level randomisation and contamination from shared clinicians, our analysis of TECTONICS does not capture benefits to patient outcomes due to overall improvements in protocol adherence, discussed below.

Strengths of TECTONICS include the low crossover rate, large and diverse population, low loss to follow up, and pragmatic design. TECTONICS was preceded by iterative development and a substantial pilot, making the intervention relatively mature. Because of the long period of ACTFAST-3 and TECTONICS (2017-2023), our analysis avoids threats to generalization from sustainability and acceptance from clinical stakeholders. Our study design was pragmatic, meaning that study elements were designed to reflect how the intervention would work in routine practice rather than in a best-case highly controlled scenario. Our trial therefore does not allow us to conclude that no settings or patients benefit from ACT-like interventions, but that in a real-world setting there were no significant differences. An exploratory subgroup analysis of only intervention patients with case reviews performed also found no benefit (Supplement Table 13). One pragmatic element of the study was the discretion of the ACT staff regarding which cases to review and which alerts to contact the OR staff about.

Although the manual of procedures and research coordinators provided guidance on using patient-acuity tools for this purpose, differences in patient characteristics between reviewed and un-reviewed cases (Supplemental Table 1) provide some insight into how ACT staff selected cases. The reviewed population had more numerous comorbidities, but there was broad overlap in patient characteristics other than minor procedure classes. Supplemental Table 13 shows an increased mortality rate among intervention cases selected for review after matching on recorded case and patient characteristics, suggesting that ACT staff selected higher acuity cases and patients based on factors other than comorbidities and surgery type. We believe this residual selection bias to be a much more likely explanation than significant harm created by ACT intervention, consistent with the primary analysis. Routine tasks and protocol adherence were the most common reasons for contacting an OR other than a case review (Table 3).

Several limitations could have led to underestimating the effects of the ACT. Although the population was large, the rate of contact between the ACT and eligible ORs was moderate (6.3%), so even a highly efficacious intervention (conditional on contact) would have had modest power. Clinicians in the OR had incomplete adherence to ACT recommendations, which may also have reduced its impact. We observed a 51% adherence rate in a random sample of OR contacts, and a prior substudy conducted for workflow analysis observed a 42% adherence rate.^15^ Incomplete adherence is a fact of pragmatic trials, and faithfully reflects the real-world effect of the intervention, although it suggests that effects in a more adherent group could be larger. Hawthorne and contamination effects are also limitations. Clinicians at the study site were aware of ACT monitoring and received frequent messages regarding common issues. Although specific physiologic alerts did not affect usual care patients, the “audit and feedback” type effect on protocol adherence and optimization of common intraoperative issues likely spilled over, affecting both study arms. For example, an anaesthesiologist caring for diabetic patients in both arms of the study might improve dysglyceamia protocol adherence in both groups based on ACT messages. A large fraction of anaesthesia staff at the site rotated through the ACT, which may also have increased protocol adherence in usual-care group patients. We initially planned a pre-post intervention analysis to elucidate these effects, but changes in the EHR and the concurrent COVID-19 pandemic made that infeasible. Secondary analyses will explore changes after the study to address this question. Although the recorded crossover rate was low, the primary analysis does not reflect the effect of the most clinically-relevant interventions made by the ACT, because the study encouraged clinicians to intervene on usual care-group patients if serious time-sensitive alerts occurred (Table 3). A clustered design in which usual-care group clinicians are truly naïve to the intervention would be the best way to detect these indirect effects. Finally, measurement error may have mitigated observed effects of the intervention. Two of our primary outcomes, delirium in the ICU and respiratory failure, were derived from routinely collected bedside assessments, and inaccuracies 28 in those data may have distorted our results. Deaths were identified from the EHR rather than governmental statistics and may miss some events after discharge if no follow-up was planned. AKI is likely under-detected due to selective measurement of postoperative creatinine. In none of these cases do we believe these errors to be differential between groups.

The study site was a tertiary academic centre, which limits the generalizability of the findings. Several aspects of the study site may explain the null result. The site’s low supervision ratio allowed anaesthesiologists to closely monitor high-risk cases, reducing the impact of cognitive load and the opportunity to miss key data.

Anaesthesia clinicians at the site sub-specialize, which tends to make them familiar with relevant protocols and best practices, ^29^ mitigating the relevance of the ACT’s recommendations. The site’s pre-anaesthesia clinic evaluated roughly 90% of patients, pre-emptively facilitating appropriate risk mitigation plans. ^20^ In settings without these redundant checks, a similar intervention may improve outcomes. Additionally, we evaluated a single set of physiologic alerts. The most common alerts were on topics only weakly related to the primary outcomes (Table 3). Although we found these alerts to fit the needs of the ACT, alerts with more advanced detection of, for example, impending hypotension, may have larger effects.

### Summary

In this large RCT, we found that intraoperative telemedicine support for anaesthesia clinicians with case planning reviews and real-time alerts did not reduce 30-day mortality, respiratory failure, AKI, or delirium. Clinicians accepted telemedicine support, and we observed many safety and process-of-care interventions, which suggests that intraoperative telemedicine should be further explored in other settings. Future work on similar telemedicine models should use multi-centre designs to detect effects mediated by quality improvement, avoid contamination due to time-critical alerts that cannot ethically be withheld from the intraoperative clinician, and explore alternative practice settings.

## Supporting information

Merged Supplement

## Data Availability

The Washington University Human Research Protection Office did not permit sharing of patient level data due to enrolment with a waiver of consent. Summary data is available at the clinicaltrials.gov registration.

## Author Contributions

All authors had full access to all the data in the study and agreed to submit it for publication. All authors contributed to the revision of the manuscript. CRK wrote the manuscript draft, processed the EHR data, and did the statistical analysis. CRK, BAF, ST, and MA implemented the machine-learning web application. ABA created the original analysis plan, contributed to the conception of the work, and analysed data for and prepared DSMB reports. CRK and ABA directly accessed and verified the underlying patient data in the study and take responsibility for the integrity of the data and the accuracy of the data analysis. BAF and CRK directly accessed and verified the case-review, and alert data in the study and take responsibility for the integrity of the data and the accuracy of the data analysis. SHG, BT, OH, PK, BH, TSW, MCP, JA, and MSA contributed to the conception of the work, implementation of the intervention, and interpretation of the results. TB contributed to the implementation of the intervention, data acquisition, and quality control of case-reviews and alert data. SM contributed to the implementation of the intervention and regulatory oversight. SG, AM, DM, MAV, and EK contributed to the implementation of the intervention and data acquisition. AK conducted EHR data acquisition and intervention informatics implementation. MSA lead the conception, design, and implementation of the work. TB and EK conducted the recommendation adherence reviews. MSA, TK, BAF, ABA, and CRK contributed to the interpretation of the data.

## Acknowledgements

Members of the ACTFAST Study Group who contributed to data acquisition or other support of the implementation are provided in the Supplement.

## Non-author collaborators

the ACTFAST author group (see supplement)

