## Supplementary material for "Effect of Telemedicine Support for Intraoperative Anaesthesia Care on Postoperative Outcomes: The TECTONICS Randomised Clinical Trial": Merged Supplement

#### TECTONICS Supplement Table of Contents

| | |
|---|---|
| SUPPLEMENTAL METHODS 1: OUTCOME DEFINITIONS | 2 |
| SUPPLEMENTAL METHODS 2: Missing outcome data | 4 |
| SUPPLEMENTAL METHODS 3: Pre-registration | 5 |
| SUPPLEMENTAL METHODS 4: Pre-specified Subgroup analyses | 5 |
| SUPPLEMENTAL METHODS 5: Database Issues Discovered During the Trial | 5 |
| SUPPLEMENTAL METHODS 6: Detecting days with no staff | 5 |
| SUPPLEMENTAL METHODS 7: “Per-protocol” exploratory subgroup analysis | 6 |
| SUPPLEMENTAL METHODS 8: Site staffing model | 6 |
| SUPPLEMENTAL METHODS 9: Defining high-risk cases | 7 |
| SUPPLEMENTAL METHODS 10: Alert criteria | 10 |
| SUPPLEMENTAL METHODS 12: Data safety monitoring plan | 11 |
| SUPPLEMENTAL METHODS 13: PRECIS-2 analysis of trial pragmatism | 13 |
| SUPPLEMENTAL METHODS 14: ACT Manual of procedures and clinical protocol | 14 |
| Welcome / Background | 14 |
| Daily routine | 15 |
| Appendix and how to use AlertWatch:OR and Epic messaging | 19 |
| Getting Started / Schedule | 27 |
| Responsibilities by role | 32 |
| SUPPLEMENTAL RESULTS: Sup Figure 1: AlertWatch:OR interface | 35 |
| Sup Figure 2: Machine learning risk web-app | 35 |
| Sup Fig 3: Enrollment by month. | 36 |
| Sup Fig 4: Primary outcomes ascertainment rate by Month | 37 |
| Sup Fig 5: Secondary outcomes ascertainment rate by Month. | 38 |
| Sup Fig 6: Covid-19 infection rates per 100k in St Louis County, Missouri | 39 |
| Sup Fig 7: Case reviews by month | 40 |
| Sup Fig 8: Alerts communicated to the operating room by month. | 41 |
| Sup Table 1: Characteristics of reviewed and unreviewed intervention group cases | 42 |
| Sup Table 2: Replies to case reviews | 44 |
| Sup Table 3: Alternative specification analysis | 45 |
| Sup Table 4: Subgroup analysis: age >=65 | 46 |
| Sup Table 5: Subgroup analysis: ASA >= 3 | 47 |
| Sup Table 6: Subgroup analysis: overlap >= 50% of case | 48 |
| Sup Table 7: Subgroup analysis: top quartile of baseline mortality risk | 49 |
| Sup Table 8: Exploratory analysis: interaction with surgical service | 50 |
| Sup Table 9: Sensitivity analysis: include subsequent cases on patients. | 51 |
| Sup Table 10: Exploratory subgroup analysis stratifying by emergency status. | 52 |
| Sup Table 11: Sensitivity analysis: un-clustered analysis | 53 |
| Sup Table 12: Main Analysis before correction for multiple testing | 54 |
| Sup Table 13: Exploratory per-protocol analysis | 55 |
| Sup Table 14: Recommendation actionability and adherence. | 56 |
| Consort Checklist | 70 |

| Nonauthor contributors | 73 |
|---|---|

#### SUPPLEMENTAL METHODS

#### OUTCOME DEFINITIONS

| Measurement | Definition |
|---|---|
| Thirty-day postoperative mortality | Death of any cause occurring in or out of the hospital, within 30 days of the index surgery. Captured by hospital vital statistics, which includes in-hospital events, routine clinical and billing follow-up. Deaths outside of hospital are incompletely captured. |
| Postoperative delirium | Defined as an acute change in consciousness or cognition. It has a fluctuating course, and is characterized by inattention, disorganized thinking and altered level of consciousness. Nursing staff in the surgical intensive care and cardiac-thoracic intensive care units regularly assess all patients using the Confusion Assessment Method for the Intensive Care Unit (CAM-ICU) instrument.[64] It is administered every 12-24 hours depending on clinical context while in the ICU. All delirium assessments within 7 days will be included, with any positive evaluations yielding a positive event. CAM-ICU scores within 30 minutes of a documented deep level of sedation (Richmond Agitation Sedation Scale < -3) are excluded. Measurements taken from nursing flowsheets in the EHR. |
| Postoperative respiratory failure | Defined as mechanical ventilation for greater than 24 hours after surgery, or re-intubation and mechanical ventilation within 30 days of surgery. Patients with tracheostomies and preoperative ventilation are excluded. Airway events within 24 hours after surgeries within 30 days of the index operation are excluded. That is, a 24-hour window of ventilation is allowed after all surgeries without triggering a positive event. Measurements taken from ventilator data and airway events documented in the EHR. |
| Postoperative acute kidney injury | Modified KDIGO stage-1 AKI using creatinine criteria: (i) an increase in serum creatinine by 50% maximum within 7 days or 0.3 mg / dL absolute increase compared with preoperative maximum within 2 days. This is a modification from the protocol due to unreliable postoperative urine output records[33,66,67] Patients with evidence of renal failure (serum creatinine > 4 mg/dL, ESRD, dialysis in the last week) are excluded. Measurements taken from laboratory values included in the EHR. |

**Secondary outcome measures and definitions.**

| Outcome | Definition |
|---|---|
| Composite outcome | The sum of the 4 primary outcomes, treating missing as “0”. |
| Temperature management | Temperature ≥ 36°C at end of surgery (maximum temperature in the final 10 minutes of intraoperative record). Measurements taken from data recorded in the EHR. |

<table>
<tr><td>Antibiotic redosing</td><td>Antibiotic redosing compliant with guidelines developed by the institutional pharmacy and therapeutics committee. Measurements taken from medication administration record in the EHR.
<table>
<tr><th>Antibiotic</th><th>Adult Dose</th><th>Redosing Interval[1,2]</th></tr>
<tr><td>Ampicillin/sulbactam</td><td>3 g</td><td>q2h</td></tr>
<tr><td>Aztreonam</td><td>2 g</td><td>q4h</td></tr>
<tr><td>Cefazolin</td><td>< 120 kg: 2 g<br>≥ 120 kg: 3 g</td><td>q4h</td></tr>
<tr><td>Cefepime</td><td>2 g</td><td>q4h</td></tr>
<tr><td>Cefoxitin</td><td>2 g</td><td>q2h</td></tr>
<tr><td>Ceftriaxone</td><td>2 g</td><td>q12h</td></tr>
<tr><td>Ciprofloxacin</td><td>400 mg</td><td>q8h</td></tr>
<tr><td>Clindamycin</td><td>900 mg</td><td>q6h</td></tr>
<tr><td>Ertapenem</td><td>1 g</td><td>q24h</td></tr>
<tr><td>Gentamicin (traditional)</td><td>1.5 mg/kg</td><td>q8h</td></tr>
<tr><td>Gentamicin (extended interval)</td><td>5 mg/kg</td><td>q24h</td></tr>
<tr><td>Linezolid</td><td>600 mg</td><td>q10h</td></tr>
<tr><td>Meropenem</td><td>1000 mg</td><td>q2h</td></tr>
<tr><td>Metronidazole</td><td>500 mg</td><td>q8h</td></tr>
<tr><td>Piperacillin/tazobactam</td><td>3.375 g</td><td>q2h</td></tr>
<tr><td>Vancomycin</td><td>< 80 kg: 1 g<br>≥ 80 kg: 1.5 g</td><td>q12h</td></tr>
</table>
</td></tr>
<tr><td>Mean arterial pressure management</td><td>Fraction of time during surgery with mean arterial pressure ≥ 60 mmHg. Measurements taken from data captured in the EHR.</td></tr>
<tr><td>Acceptable peak inspiratory pressure with mechanical ventilation</td><td>Fraction of time during surgery with peak inspiratory pressure ≤ 30 cmH2O. Only cases with mechanical ventilation are included. Mechanical ventilation is detected with sustained EtCO2 > 20 mmHg, tidal volume > 150 mL, driving pressure > 6 cmH2O, PEEP > 2 cmH2O. The first and last 8 minutes are excluded to account for activities around intubation and extubation. Measurements taken from data captured in the EHR.</td></tr>
<tr><td>Hyperglycemia</td><td>Blood glucose > 200 mg/dL at end of surgery. Data taken from laboratories included in the EHR.</td></tr>
<tr><td>Anesthetic delivery without gaps</td><td>Patients without ≥ 15 consecutive min of volatile anesthetic concentration ≤ 0.3 MAC during mechanical ventilation, defined as in “Peak inspiratory pressure” above. Patients with infusions of sedatives at greater than the following doses are excluded: propofol > 40 mcg/kg/min, ketamine > 0.3 mg/kg/hr. Measurements taken from data captured in the EHR.</td></tr>
<tr><td>Fresh gas flow rates</td><td>Proportion of patients with efficient fresh gas flow for ≥90% of the mechanical ventilation period defined above. “Efficient” fresh gas flow is defined as <= 2 L/minute for sevoflurane and <= 1 L/minute for isoflurane and desflurane. Only times with > 0.3 age adjusted MAC are included. Measurements taken from data captured in the EHR.</td></tr>
</table>

**Alternative specifications**

| Outcome | Definition |
|---|---|
| ICU utilization | Total hours admitted to the ICU in the first 7 postoperative days |

| | |
|---|---|
| Average intraoperative temperature | Average of measured temperatures intraop. Temperatures below 34 degrees C excluded (most likely either cardio-pulmonary bypass or measurement errors), averaged within surgery |
| Time-weighted hypotension | Sum over intraop measurements of arterial and NIBP of Maximum( 65-MAP, 0) * (time until next measurement). Where Systolic and diastolic recorded but MAP not, MAP approximated by 1/3 SBP + 2/3 DBP. |
| Change in serum creatinine | Ratio of postoperative maximum creatinine in 7 days to last preop creatinine. Analyzed on log scale. |
| Average peak inspiratory pressure | Defined on same time window as "Peak inspiratory pressure with mechanical ventilation" , averaged within surgery |
| Average time with efficient gas flow | Defined on same time window as, "Fresh gas flow rates", averaged within surgery |
| Missing delirium | Coding patients with no measured CAM-ICU as "negative" instead of "missing". |

**Missing outcome data**

Missingness in all the primary endpoints is informative, and therefore usual imputation methods for missing-at-random data are not applicable.

- Mortality: Patients discharged alive without subsequent medical system contact are assumed to be alive at 30 days.
- Respiratory failure: Patients without documented mechanical ventilation events are assumed to have no event. Patients discharged from hospital in less than the at-risk window are assumed to have no event.
- Delirium: Patients without CAM-ICU scores are not included in the analysis. No other imputation is used.
- Acute kidney injury: Patients without baseline creatinine measurements are assumed to have normal values (single imputation) for their age, sex, weight, race, and CKD variable using an XGBoost (Chen and Guestrin 2016) imputation model from all patients with measured baseline creatinine. Patients without postoperative creatinine measures are assumed to not have acute kidney injury.

Patients with missing secondary outcomes are not included in the analysis of those outcomes.

Chen, Tianqi, and Carlos Guestrin. 2016. "XGBoost: A Scalable Tree Boosting System." In *Proceedings of the 22nd ACM SIGKDD International Conference on Knowledge Discovery and Data Mining*, KDD '16, New York, NY, USA: Association for Computing Machinery, 785–94. doi:10.1145/2939672.2939785.

**PRE-REGISTRATION**

The protocol for the study including an analysis plan was finalized with the Washington University IRB prior to the enrollment of the first participant. During the initial registration of the study with clinicaltrials.gov (NCT03923699), administrative staff were unclear on how to register co-primary outcomes, and input the four primary outcomes as secondary outcomes with a general statement ("complications") as the primary outcome. An update to the clinicaltrials.gov registration was made to bring it into agreement with the IRB-approved protocol.

**PRE-SPECIFIED SUBGROUP ANALYSES**

- ASA >= 3
- Age >= 65
- Cases with >50% overlap with the hours of operation of the ACT
- Top quartile in procedure-based mortality risk.
- Excluding patients without diabetes from blood glucose analyses
- Stratification by Surgical service

Multiple-comparisons corrections are performed within each subgroup in the same manner as the primary analysis. No additional correction to applied to account for multiple subgroup analyses.

A non-prespecified subgroup analysis according to ACT interventions (approximating a per-protocol analysis) was requested by reviewers and is described in a subsequent section.

**DATABASE ISSUES DISCOVERED DURING THE TRIAL**

In May 2020, the sever hosting the database containing ACT reviews and alert information was upgraded due to hardware end-of-life. This resulted in a block of 67 days between the last backup and the new server being operational where study records were not stored or could not be retrieved. During the upgrade, it was found that the existing database for case reviews used a 255 character field to store review data, which was much shorter than the actual data, resulting in most of the information being lost, including the specific recommendations, whether the review was sent, and whether the OR clinician responded.

**DETECTING DAYS WITH NO STAFF.**

Study assistants and coordinators did not log the days with no medical staff available. It was planned to detect these days by periods with no interaction recorded in the ACT dashboard interface. However, we found that on days where medical staff were pulled to direct clinical work, they (or study assistants) may have logged into the interface before having to leave. Days without study personnel were therefore detected using a requirement for at least 2 interface interactions separated by 2 hours, and for activity after 10:00. This requirement also excludes days where the database storing trial records was non-functional.

**"PER-PROTOCOL" EXPLORATORY SUBGROUP ANALYSIS**

Reviewers hypothesized that the patients whom the ACT felt contact was warranted in may represent a subgroup in which greater benefit of the intervention was likely. Unfortunately, we do not know which usual care group patients the ACT would have initiated communication about had they been in the intervention group. Our best response is to approximate this group using a propensity matching method.

Within the intervention group, we used baseline characteristics (demographics, diagnoses, pre-procedure evaluation variables such as ASA status, procedure words, surgical service) likely related to the decision to perform a case review to predict whether or not a case review was sent to the intraoperative team. The predictive model was an XGBoost model, using hyperparameters and word tokenization selected for identification of high-risk cases in the pilot-study dataset (see "DEFINING HIGH-RISK CASES" below). Several alternative sets of hyperparameters were tested, with negligible differences. Linear models were also tested, with the same set of conclusions.

We then used the fitted model to obtain propensity scores in the usual-care group and 1:1 matched each intervention group patient with an ACT case review to a usual care group patient using the optmatch package version 10.7. Matching was stratified on surgical service and sex for computation reasons. The matched set was analyzed using the same methods as the primary analysis without considering the matching strata.

While we can match baseline characteristics for case reviews, the decision to communicate alerts to the OR likely depends heavily on intraoperative data. These data are post-randomization, and potentially affected by earlier communication (such as a case review or earlier alerts). Additionally, intraoperative data is very high dimensional, including all medication events, monitor data, and the alerts themselves. Preliminary attempts at representation learning and forecasting methods to predict alert events was not successful, and we therefore present results using baseline characteristics to predict the propensity to case review or alert communication from the ACT to the OR. Cases where the OR initiated contact are included as positive events.

Because of confounding due to variables not included in the propensity model, we expect that both groups will be somewhat anti-conservative, failing to identify usual-care group patients with the same degree of procedural risk and intervention group patients. This bias is likely stronger in the alert group.

**SITE STAFFING MODEL**

At the study site, the anesthesia model is "medical direction." This requires anaesthesiologist's involvement at multiple portions of a case, including induction / intubation and emergence. Supervised residents are expected to discuss their anaesthetic plans for scheduled cases with the anesthesiologist on the previous day. CRNAs are not expected to have reviewed cases before the day of surgery, but they are expected to have some discussion (proportional to the complexity of the case) with the anaesthesiologist prior to the start of the case. The number of rooms assigned to an anaesthesiologist varied among locations within the study site, with cases with higher rates of planned ICU admission, invasive lines, and other complexities having lower room counts (usually 2-3 rooms) and up to four rooms per anaesthesiologist in cases with lower risks of complications and staffing needs. A senior anaesthesiologist is assigned "pod leader" in each surgical area (from a rota), who creates daily assignment lists, addresses add-on and urgent cases, and is the first-line resource for anaesthesiologist questions and advice in most circumstances.

**DEFINING HIGH-RISK CASES**

An xgboost model was fitted to data from before TECTONICS (including ACTFAST-3, as no outcome effect was found in that study), using words in case descriptions occurring greater than 100 times (683 most common words) as prediction features and death within 30 days as the outcome. Hyper-parameters were selected by cross-validation in the training data and are listed below. The top quartile in predicted mortality risk (threshold = 0.0189) was used as the "high risk" group.

XGB parameters:

Depth=5, eta=0.71875, nrounds=12, min_child_weight=3.266351688, gamma=3.4375, subsample=0.871875, lambda=4.003113252, num_parallel_tree=1

Input Tokens used:

| | | | | |
|---|---|---|---|---|
| bleeding | lobe | room | polypectomy | dynamic |
| long | cyclophotocoagulation | imas | moma | penis |
| interpedicullary | stryker | instrumented | minimal | lengthening |
| vim | bleed | expander | middle | cholangio |
| common | pancreatography | cystocele | illeal | pso |
| at | chiari | exenteration | peroneal | plif |
| c7 | antrostomy | ercp | saphenous | metrx |
| unilateral | pyeloplasty | nail | thalamic | thymectomy |
| heartware | medial | transthoracic | laminar | index |
| circumcision | pneumonectomy | svg | drive | excisional |
| intraocular | reimplant | adjustment | proctocolectomy | saturation |
| taping | transvaginal | ventriculostomy | rib | colectomy |
| flank | vaporization | microsurgery | line | olecranon |
| balloon | mastoidectomy | inflatable | ethmoidectomy | lateral |
| completion | zygomatic | hepatectomy | transperineal | halo |
| from | add | adhesions | hidradenitis | third |
| ktp | nephroureterectomy | bile | lima | flexible |
| gas | parastomal | bifemoral | dislocation | botox |
| maxillectomy | morphogenetic | hydrocelectomy | thumb | defect |
| axillo | lysis | blue | enterocele | patch |
| pouch | anastomosis | c6 | simple | gastrojejunostomy |
| myectomy | vocal | cpa | heel | calcaneous |
| manipulation | nephew | ligament | thyroplasty | l2 |
| aorto | resurfacing | the | c3 | hemorrhoidectomy |
| rhizotomy | thoracoabdominal | trans | esophagus | smith |
| wide | peel | urethrotomy | acoustic | malformation |
| thorocotomy | rectus | bso | decortication | lymphoscintigraphy |
| ring | c4 | condyloma | orchiectomy | dental |
| ebus | glaucoma | venous | colpopexy | wean |
| subthalmic | globe | adhesiolysis | body | control |
| facial | orbital | transanal | unicompartment | change |
| depuy | heller | omentectomy | proctectomy | autolitt |
| monteris | transposition | poudrage | ruptured | colporrhaphy |
| endoscopy | on | root | c5 | mucosal |
| pleuroscopy | ileocolic | level | washout | found |
| talc | guyons | neuroma | proctoscopy | localization |
| tibioperoneal | replant | fat | surgical | retroperitoneal |
| bronchus | septoplasty | duct | arteriovenous | lead |
| pilon | endoluminal | pleurodesis | canal | end |
| sphincter | via | suprapubic | atrial | foraminotomy |
| complex | stone | pregnancy | ectopic | buccal |
| patella | ivor | lewis | adrenalectomy | prosthetic |

| | | | | |
|---|---|---|---|---|
| cervix | pericardial | endolaser | fasciotomy | micro |
| ischium | not | scan | sleeve | target |
| fossa | interstim | ileal | laminoplasty | l3 |
| parathyroidectomy | si | vacuum | mandibulectomy | laryngectomy |
| cath | eyelid | urethroplasty | arthrodesis | screw |
| embolectomy | septal | intramedullary | jejunostomy | myotomy |
| plate | ear | port | ascending | s1 |
| air | artificial | glossectomy | aicd | anus |
| axillary | ileo | novasure | sling | trigger |
| shaft | window | corpectomy | clipping | re |
| pinning | colonoscopy | mini | metacarpal | aorta |
| clavicular | l5 | ovarian | remote | subclavian |
| tracheal | nasal | median | dilation | leep |
| ulnar | autograft | forearm | mastectomy | irrigation |
| sacral | sacrum | fulguration | splenectomy | minimally |
| turp | endo | invasive | acetabulum | gauge |
| zimmer | pancreatectomy | perineal | axilla | l4 |
| fingers | cavity | duodenoscopy | low | vulvectomy |
| joint | pacemaker | perirectal | craniectomy | allograft |
| urethral | xi | temporal | filter | donor |
| egd | leads | corporeal | eswl | uterus |
| redo | vitrectomy | endometrial | colposcopy | prosthesis |
| extra | scrotum | oral | takedown | nissen |
| esophagectomy | frequency | labral | radio | suction |
| holes | stealth | gastrectomy | guided | circulation |
| burr | tissue | extradural | fluid | toe |
| veins | wedge | vulva | for | conduit |
| dialysis | closed | paraesophageal | mesh | tricuspid |
| post | generator | electrode | reexploration | saver |
| oxygenation | tonsillectomy | transfer | mandible | cell |
| abscess | vena | cava | buttock | fundoplication |
| bead | parotidectomy | periacetabular | plateau | epidural |
| heartmate | maze | omniguide | oopherectomy | tubal |
| fibula | cholangiogram | penile | crest | hemiarthroplasty |
| sigmoid | antibiotic | spacer | angioplasty | intraoperative |
| intrauterine | debulking | thyroidectomy | peg | esophageal |
| radial | umbilical | trauma | muscle | needle |
| ecmo | major | throat | cystectomy | thrombectomy |
| incisional | rigid | interbody | anal | injection |
| endobronchial | navigation | drain | wall | approach |
| ulna | tibial | back | ultrasound | ii |
| preoperative | membrane | stage | above | tube |
| neurogenic | access | below | to | uterine |
| sentinel | fess | mouth | loop | hysteroscopic |
| popliteal | gu | myomectomy | mediastinoscopy | wrist |
| pancreatoduodenectomy | appendectomy | stripping | hiatal | minor |
| ureter | vaginal | tendon | perineum | application |
| mandibular | tenotomy | osteoplasty | possible | salpingectomy |
| radius | vascular | proximal | subdural | battery |
| endarterectomy | gastrostomy | sternotomy | deep | face |
| nephrolithotomy | elbow | ventral | esophagogastroduodenoscopy | phacoemulsification |
| prostatectomy | vein | cataract | carotid | local |
| vagina | vessel | peritoneal | hematoma | internal |
| osteotomy | shunt | sinus | turbt | lens |

| | | | | |
|---|---|---|---|---|
| carpal | groin | colostomy | harvest | assist |
| angiogram | evacuation | iliac | microscope | outlet |
| humerus | external | tunnel | heart | prostate |
| gastric | microlaryngoscopy | pectoralis | radical | rectum |
| reconstruction | urethra | free | co2 | lung |
| inguinal | hypophysotomy | transsphenoidal | pelvic | ligation |
| foot | antegrade | pelvis | catheter | stimulator |
| ablation | ventricular | brain | ball | aneurysm |
| socket | pyelogram | finger | sternal | kidney |
| reduction | video | arthroscopy | thoracotomy | hepatic |
| ileostomy | procedure | thigh | laminectomy | nerve |
| lobectomy | morphogenic | bowel | laryngoscopy | partial |
| multiple | small | teeth | transurethral | mitral |
| creation | protein | vac | exploration | nephrectomy |
| implant | retrograde | split | lithotripsy | breast |
| operative | upper | release | liver | imri |
| fixation | thickness | percutaneous | ankle | tibia |
| hardware | head | im | skin | distal |
| gyn | colon | coronary | hand | vats |
| amputation | monitoring | nailing | hysteroscopy | tracheostomy |
| closure | bladder | device | surgery | pump |
| fistula | spine | femoral | laser | esophagoscopy |
| cholecystectomy | fracture | curettage | diagnostic | ureteroscopy |
| cord | extremity | aortic | extraction | exchange |
| femur | transplant | av | reverse | debridement |
| flap | bronchoscopy | fiberoptic | node | chest |
| lymph | leg | open | or | discectomy |
| mass | cyst | lower | craniotomy | exam |
| under | hernia | oophorectomy | shoulder | anterior |
| decompression | eye | valve | anesthesia | dilatation |
| bypass | laparoscopy | salpingo | artery | insertion |
| wound | dissection | lesion | neck | with |
| revision | ureteral | endoscopic | tumor | arm |
| robotic | hysterectomy | bilateral | biopsy | bone |
| thoracic | of | cervical | spinal | abdominal |
| assisted | orif | abdomen | exploratory | Lumbar |
| laparotomy | removal | instrumentation | cystoscopy | Posterior |
| stent | knee | resection | repair | Excision |
| placement | incision | fusion | drainage | Hip |
| graft | replacement | arthroplasty | left | Total |
| right | laparoscopic | and | | |

**ALERT CRITERIA**

Definitions for common alerts (Table 3). Where multiple definitions exist on the same parameter, a “yellow” and “red” alert existed.

| **Alert Group** | **Triggers** |
|---|---|
| Glucose | Glucose measurement > 2 hours old (T2DM) |
| | Glucose measurement > 1 hours old (T1DM) |
| | Glucose measurement > 1 hours old (insulin administered) |
| | Glucose Outside 70 to 200 mg/dL |
| | Glucose Outside 60 to 300 mg/dL |
| Temperature | No temperature monitoring 45 minutes |
| | Core Temperature Outside 36.0 to 37.5 C |
| | Core Temperature Outside 35.0 to 38.0 C |
| Antibiotic | No antibiotic detected 10 minutes after incision |
| | Institutional redosing table (see secondary outcome definition) |
| Hypotension | MAP < 65 mmHg |
| | MAP < 60 mmHg |
| | MAP low time > 10 minutes |
| Volatile Anesthetic Concentration | < 0.7 MAC |
| | < 0.5 MAC |
| Train of Four | NMB administered, no TOF in 1 hour |
| Blood Pressure Monitoring | no BP measured 10 minutes |
| Fresh Gas Flow | If desflurane: FGF > 1.2 L/min |
| | If isoflurane or sevoflurane: FGF > 2.2 L/min |
| Potassium | Outside 3.5 to 5.0 mmol |
| Oxygenation | SpO2 < 93 |
| | SpO2 < 90 |
| Tidal Volume | Tidal volume Outside 6-8 mL/kg ideal body weight (4-6 ml/kg with one-lung ventilation) |
| Tachycardia | HR > 100 |
| Lung Compliance | PIP > 30 |
| | PIP > 40 |

**TECTONICS Trial - Data and Safety Monitoring Plan**

We shall implement a Safety Monitoring Committee (SMC) in compliance with the NINR DSM policy. The monitoring plan for this study is appropriate for a minimal risk pragmatic trial.

The SMC will consist of:

- The Principal Investigator (PI)
- Two other members of the study team (Bernadette Henrichs and Troy Wildes)
- Two additional members, who have expertise in anesthesia and are independent of the study
- A biostatistician (Arbi Ben Abdallah) will also participate as a member of the SMC

The SMC will review all adverse events, compliance with IRB requirements, investigator compliance. The SMC will advise on approaches to minimize risks to participants and to protect the confidentiality of participants' data. Adverse events and unanticipated problems will be identified through review of data from the electronic medical record. The PI together with two designated members of the study team (Bernadette Henrichs and Troy Wildes) will assess if a risk or adverse event is potentially related to the conduct of the study. The opinion of **<u>any</u>** of these three that a risk or an event is potentially related to the conduct of the TECTONICS study will be sufficient grounds for reporting to the IRB and to the NINR. As such, all adverse events determined to be potentially related to the study and all unanticipated problems involving risks to subjects or others will be reviewed by the study team, and will be reported to the IRB at Washington University and to the NINR, according to NINR stipulations.

- Reporting to the NINR of any intra-operative death will be within 24 hours.
- Intraoperative serious adverse events (SAEs) and adverse events (AEs) will be reported to the NINR within 7 days. These serious adverse events and adverse events will include:
    - Seizures
    - Anaphylaxis
    - Myocardial infarction
    - Cardiac arrest
    - Unexpected serious cardiac arrhythmias (e.g. ventricular tachycardia, ventricular fibrillation) – In some surgeries, like cardiac surgeries, these arrhythmias can be anticipated
    - Pulmonary edema
    - Pneumothorax
    - Hypoglycemia
    - Breech of patient confidentiality
    - Information on the wrong patient communicated from the Anesthesiology Control Tower to operating room clinicians
    - Factually incorrect information communicated from the Anesthesiology Control Tower to operating room clinicians
    - Undesirable distraction of operating room clinicians attributable to communications from the Anesthesiology Control Tower

<u>The Principle Investigator will be responsible for reporting these (and other) adverse events to the NINR</u>. In reporting these events to the NINR, the Principal Investigator and study team members will provide an informed opinion to the NINR regarding whether or not these events are potentially related to the conduct of the study. We anticipate that these adverse events will occur very rarely, and when they do occur, will not be likely to be related to the conduct of the study.

The study team will prepare reports for the SMC, NINR and the Institutional Review Board (IRB). The SMC will review the study data for safety every six months. The study team will monitor the study for compliance with IRB requirements on an ongoing basis and review aggregate data at least annually. If new information becomes know from external factors that may have an impact on the safety of participants or on the ethics of the TECTONICS trial, the information will be reviewed by the PI, the SMC and the IRB at Washington University.

The SMC and PI, in consultation with the IRB, might recommend halting the trial if there are unexpected safety events that are potentially attributable to the conduct of the TECTONICS trial. If negative outcomes are more prevalent in the intervention group for the tracked study outcomes, or if clinicians in the operating room report that communications from the Anesthesiology Control Tower prevent them from providing high quality care to patients in the operating room, the study may be stopped. The following are the negative outcomes that will be specifically tracked for this purpose:

- Postoperative delirium
- Postoperative acute kidney injury
- Postoperative respiratory failure
- Postoperative 30-day mortality

Focus groups will be conducted with operating room clinicians by members of the study team on an annual basis to ascertain whether the conduct of the TECTONICS trial is having a negative impact on their ability to provide high quality clinical care. We can think of no obvious mechanism by which an Anesthesiology Control Tower that communicates appropriately and thoughtfully with members of the operating room anesthesia care team could jeopardize the safety of surgical patients. Summaries from the SMC will be provided annually to the IRB and NINR.

Risks of physical harm to the patients attributable to the conduct of the TECTONICS trial are extremely unlikely, since the intervention by clinicians in the Anesthesiology Control Tower is communication with clinicians in the operating room, who will continue to be responsible for decision making in relation to the surgical patients. Anesthesiology Control Tower communications increase awareness of the operating room team regarding important patient factors, impending risks, and best management practices. It is possible that incorrect information could be communicated in these messages, however information is entirely channeled through the active operating room care team which inherently involves verification of patient details. It is also possible that discussion of important patient issues could result in attention being deviated away from other ongoing patient issues. However, communications can be postponed at the discretion of operating room team to attend to competing care issues, and this concern has not been realized with extensive experience to date in the Anesthesiology Control Tower.

Since the clinicians in the Anesthesiology Control Tower will be investigating and discussing patients who are in the operating room, there is a minimal risk of breach of confidentiality. However, all the participants in the Anesthesiology Control Tower are trained in good clinical practice and are bound to respect Health Insurance Portability and Accountability Act (HIPAA) regulations.

Monitoring of quality of the TECTONICS study interventions (i.e. communications between the Anesthesiology Control Tower clinicians and the operating room clinicians) will be conducted annually on a randomly selected sample of the comprehensive review summaries and the alert reviews, which will be generated by the clinicians in the Anesthesiology Control Tower.

**PRECIS-2 ANALYSIS OF TRIAL PRAGMATISM**

| Domain | Score | Rationale |
|---|---|---|
| Eligibility | 5 | All adult patients undergoing surgery in regular hours. |
| Recruitment | 5 | All patients at the study site were included with a waiver of consent |
| Setting | 1 | The study is single-center. The study site is a large academic medical center with high anesthesiologist ratios and a highly used pre-anesthesia clinic. The study site has low rates of non-Black non-White race. |
| Organization | 3 | Clinicians received a manual of procedures to review and a study coordinator (a non-practicing MD) participated in the ACT. |
| Flexibility (delivery) | 4 | Clinicians had discretion on which cases to review and which alerts to respond to. The ACT system ranked patient comorbidity burden and procedure complexity to encourage reviewing patients with greater risks. Alert setting were fixed after the pilot trial. |
| Flexibility (adherence) | 5 | Intraoperative clinicians were not required to follow ACT recommendations, nor were any actions taken to suggest that they ought. ACT participants did not have specific requirements for case reviews or alert responses, although coordinators were provided goals. |
| Follow up | 5 | Follow up captured automatically from the EHR |
| Primary Outcome | 5 | Outcomes represent major adverse events after surgery (death, respiratory failure, kidney injury, delirium) |
| Primary Analysis | 5 | Intention to treat with all available data was used in the analysis. |

#### Welcome to the Anesthesiology Control Tower (ACT)!

We are excited to have you work with us in the ACT! In this document, you will find a brief summary of the ACT concept and technology, and the studies that our team is carrying out. Subsequently, there is a list of ACT daily expectations, as well as a glossary of terms. Throughout the document, practical tips and important information are provided.

#### BACKGROUND

##### THE ACT

The ACT is modeled after an air traffic control tower for a busy airport. Just as a control tower monitors each aircraft and delivers additional information and alerts to the pilot and co-pilot, the ACTors will engage with each team of OR anesthesia clinicians in a similar fashion to assist them in providing safe, effective, and efficient care for their patients. ACT will use the AlertWatch system to continually monitor the 58 ORs (focusing on contact rooms) and will obtain additional data from our existing electronic records (i.e. Epic). It is expected that ACT clinicians will filter and prioritize information in order to provide high quality alerts with minimal false alarms. Over time, we will incorporate machine learning algorithms that we are developing into the ACT platform. *This conceptualization of the ACT will allow us to decrease the burden of false and intrusive alarms in the OR and instead provide empowering and unobtrusive IT-based support to our colleagues.*

##### ALERTWATCH

AlertWatch was developed to provide visual summaries and alerts based on the physiologic state of patients, taking into account each individual patient's comorbidities. AlertWatch captures information from both patient monitors and electronic medical records and analyzes the data to determine the current patient condition within the confines of pre-existing comorbidities (please see the figures at the end of this document). A composite view of all ORs indicates the real-time status of each operating suite, including which, if any, ORs have actual or potential safety or quality issues. On the composite view, patient-specific alerts cause notifications to appear in the OR in which they are triggered. These alerts are based on accepted anesthesia practices and serve to prompt the anesthesia practitioner to provide a treatment or perform a specified duty. *ACT will use AlertWatch to review most active alerts and will critically evaluate each alert for significance. ACT clinicians will subsequently communicate supplementary notifications to OR clinicians based on the ACT clinical judgment.*

##### OUR STUDY

Our group is conducting three studies to achieve the following objectives: (i) develop, refine and validate forecasting algorithms for adverse perioperative outcomes; (ii) assess the usability of an ACT for the operating suite; and (iii) assess whether the ACT improves clinician compliance with standards of care and patient outcomes. <u>The design for the current study is a one year randomized controlled evaluation</u>, in which all ORs will be randomized on a daily basis to either a control group (28 ORs) or to an experimental group (28 ORs). ORs in the experimental arm will receive supplemental alerts from the ACT based on the judgment of the physicians in the ACT.

#### DAILY ROUTINE

***The overall objective as an ACT clinician is to monitor the 58 South Campus and Parkview Tower ORs. ACT will use AlertWatch, with additional information from Epic, to complete case reviews and address alerts that appear (as indicated by checkmarks). ACT clinicians will document their assessment of each alert in the AlertWatch platform, as described below. The ACT will be staffed on weekdays from 0700-1600 (0800 start on Wednesdays).***

***Help documents and videos are available on the department intranet, under the "ACT" link. Videos cover a general introduction to the ACT in addition to help with specific features.***

##### Prior to OR start

1. Log in to required software programs
    a. AlertWatch: Alert Watch Link
        i. Username and password are same as EPIC credentials.
        ii. By right-clicking on an AlertWatch tab and clicking "duplicate," multiple AlertWatch tabs can be opened and dragged to separate windows
        iii. Drop down menus at top left corner of census allow you to change the view between the grid view and alert view
        iv. The grid view provides a summary of all active ORs, along with active alerts (Figure 1)
            1. Room color is determined by status of case
        v. Grid view arranges ORs in numerical order and shows alert icons with no additional informationThe Alert view lists details for all active alerts as well as Tower Notes.  ORs arranged in order of alert urgency (Figure 2)
        vi. Clicking on an individual OR pulls up the **Patient Display** (Figure 3a, 3b)
            1. Clicking on specific organ systems will allow you to view vital sign trends
            2. Clicking on the labs will allow to trend intra-op lab values
            3. The "handoff" button on the bottom left of this screen provides a summary of patient comorbidities, and the case vital signs. Comorbidities should be verified against the Epic record (Figure 3b, 4)
    b. Epic: Access Epic via desktop icon or via department Intranet (When obtaining information from Epic, *ensure that the patient you have selected is the same patient you are viewing in AlertWatch)*
    c. Identify cases and patients which have the potential to require a higher level of monitoring/intervention
2. Additional useful resources that may be helpful at times:
    a. Department protocols (log in: your WUDA ID and password)
    b. ACT specific protocol library (under development)

c. Becker Medical Library-Full access to all resources if on campus network
d. AlertWatch full User Guide
e. Roles and responsibilities documents to ensure understanding of scope of work in the tower

##### After OR start

Monitor all cases on AlertWatch-ACT census (South Campus and Parkview Tower), complete case reviews (indicated by grey diamond), and address alerts (indicated by red/black checkmarks).

1. **Case Reviews:** Clinicians should prioritize patients with higher risk scores, and ideally earlier in case progression, that have been randomized to a contact room. All cases with an orange note pad alert should be reviewed as early as possible, though occasionally the orange note pad alert factor may be discovered to be less worrisome.
    a. <u>Clinicians will prepare cases as “Tower Notes” by completing case assessments (grey diamond in patient display).</u>
    b.
    c.
    d. <u>Every case review must be discussed with the attending, on average, in three minutes or less prior to sending a message to the OR clinician.</u>
    e. <u>Notepads</u>
        i. 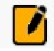 (orange): Identifies patients with specific increased risk (Ex: HOCM, pulmonary hypertension, type 1 diabetes, etc…). Having an orange notepad increases your risk score by 100, and shifts these patients to the top of the review view.
        ii. 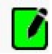 (green): Indicates intraoperative quicknotes documented in Epic.
        iii. 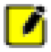 (yellow): Indicates a patient specific note/reminder in the alert assessment written by the telemedicine team [shows in Alert and Review view].
2. **Alert Reviews: Clinicians should address all alerts, irrespective of randomization status.**
    a. Each organ system is associated with pre-specified alerts that will be triggered when patient parameters fall outside of an accepted range (see glossary)
        i. Red usually indicates a higher priority alert than a black alert.
        ii. Squares indicate that a text alert is currently present within the patient display for that room (on right hand side of screen display).
        iii. Triangles indicate that a laboratory or physiologic parameter is out of range
    b. MAP goals can be changed as appropriate for each patient (Figure 3a) using the drop down in the bottom left hand corner of the “Patient View”
    ***c.*** Certain alerts will require ACT response or action. **These actionable alerts, when present, will trigger a red or black checkmark to appear in the OR in which they arise.** These checkmarks will remain until the alert is addressed by the ACT. **Note that not all alerts (triangles and squares) will necessarily trigger a checkmark.** Therefore, a room may have alarms that are not currently associated with a checkmark. If there is a clinical situation

that you think warrants an intervention, you may add an alert yourself by clicking on the green checkmark on the patient display for that room.

d. ACT will assess each alert and **will address it** by documenting in AlertWatch. To do this, ACT should **navigate to the patient display and click on the checkmark in the lower left corner of the screen (Figure 3b) and click on “Add Assessment”** next to the relevant alert (Figure 6a)

  i. This opens a **Case Review** (Figure 6b) dialogue in which clinicians will document assessments and actions for each alert, with the following definitions:

    1. Assessment: ACT evaluation of the severity of the problem (i.e. significant, possibly significant, etc)
    2. Action: What action should be taken in response to the specific alert
    3. Comment: Any other information that the ACT clinician wishes to include
    4. Reaction: Only available for study ORs- wait to fill this out until you hear back from OR staff (until it is filled out checkmark will appear as yellow)
    5. “Irrelevant alert, disable for entire case ”

      a. If you feel that an alert is entirely insignificant, clicking this will let you bypass having to enter an assessment and will suppress the alert for the remainder of the case.

  ii. Once all components of the alert have been addressed, click “Save”

    1. The checkmark in the far left column becomes the timestamp for your work.
    2. If you address all active alerts within a room, the checkmark on the census view will turn green, indicating that the ACT have documented at least one response to each alert, and there have been no new alerts. The checkmark will become red or black again if another alert is triggered within that room.

3. Contact vs Non-contact

  a. The information on a particular OR’s assignment (control or experimental) on a given day is found in the bottom left corner of Patient View or Grid view, and just before the procedure description in the Alert View (Figure 2)

  b. Providers assigned to control group ORs will not be contacted; however, the ACT will **otherwise perform the same documentation in AlertWatch**. Alerts or communications that would have otherwise been sent will be recorded.

4. ACT clinicians will select interesting, difficult, or long cases in which likelihood of ACT impact is higher for full case presentations regardless of alert status

  a. ACT clinicians will prepare cases and make notes in the “Tower Note View” for each of these patients. To do this open the Patient View and click on the note pad in the bottom left corner (Figure 7). This opens the “Tower Note View” (Figure 8). Click on “*Add*” or “*Edit*” to create a tower note or modify an existing one. Tower notes appear on the “Alert View” below the associated OR and any alerts.

   i. The Tower Notes can also be edited and created by clicking on the checkmark view as one would to address alerts and clicking "Add" or "Edit" in the top left corner of the pop-up (Figure 6a).
   ii. Note pad may be clear, yellow, or orange (Figure 7)
   iii. Tower Notes (Figure 8) should include the following:
      1. Patient age, sex, and procedure
      2. Comorbidities
      3. Potential problems (ie. myocardial infarction, stroke, dysglycemia)
      4. Strategies for risk mitigation
      5. Links to any articles relevant to the patient's care

b. After a tower note is prepared, the individual will present the patient to the group for discussion. Tower notes can then be edited as necessary to reflect the discussion and the attending's final assessment and recommendations for the patient

5. Contacting Rooms – Rooms can be contacted through Epic Secure Chat, Epic Staff Messaging System or by directly calling the clinicians
   a. To send an Epic Secure Chat message (the primary mode of communication between the tower and OR clinicians) open the patient chart…
      i.
      ii.
      iii.
      iv. Smart-phrases can be activated within a temporary staff message to leverage auto-generated content, and then pasted into a SecureChat conversation (which is more user friendly to read/respond)
   b. To send an Epic Message regarding a specific patient, first open the patient chart.
      i. In the bottom left hand corner of the Navigator, click "*More*"→ "*Send Message*"→"*Staff*" (Figure 9a). This opens the messaging platform with a link to the patient being discussed (Figure 9b). **This message is NOT part of the patient chart**
      ii. There are several smart texts available to expedite messaging. These all begin with **.actow**__
      iii. There are specific smart texts for glucose and temperature alerts as well as a general template for patients with general recommendations. Use F2 to quickly move between and complete the *** fields
      iv. If you free text a note instead of using a smart text please include a statement indicating that you would like a response from the OR clinician affirming they have received the message
      v. Messages can be sent regarding specific alerts or for general case discussion and management
         1. For cases that were discussed fully, copy and paste the "Tower Note" into the either the 'basic' smart text or into a free note (Figure 10).

**c.** After a message has been sent please addend the tower note to indicate that a message was sent.

**i.** If no response is received within 15-20 minutes call one of the OR clinicians to confirm that they received they message.

**d.** If the attending prefers to forgo the messaging system and just call the OR clinicians that is fine as well.

#### Education

Education is a vital part of the ACT.  While case based learning should occur throughout the day as a part of patient discussions residents, attendings, SRNAs, and CRNAs should all prepare and conduct a discussion on an educational topic.

- o Attending will prepare and conduct one core topic discussion (5-10 minutes) from the core topic library (under development).

- o CRNA, SRNA, and Resident will prepare and conduct one mini topic discussion (1-2 minutes) from the mini topic library (under development).
  - 

#### APPENDIX

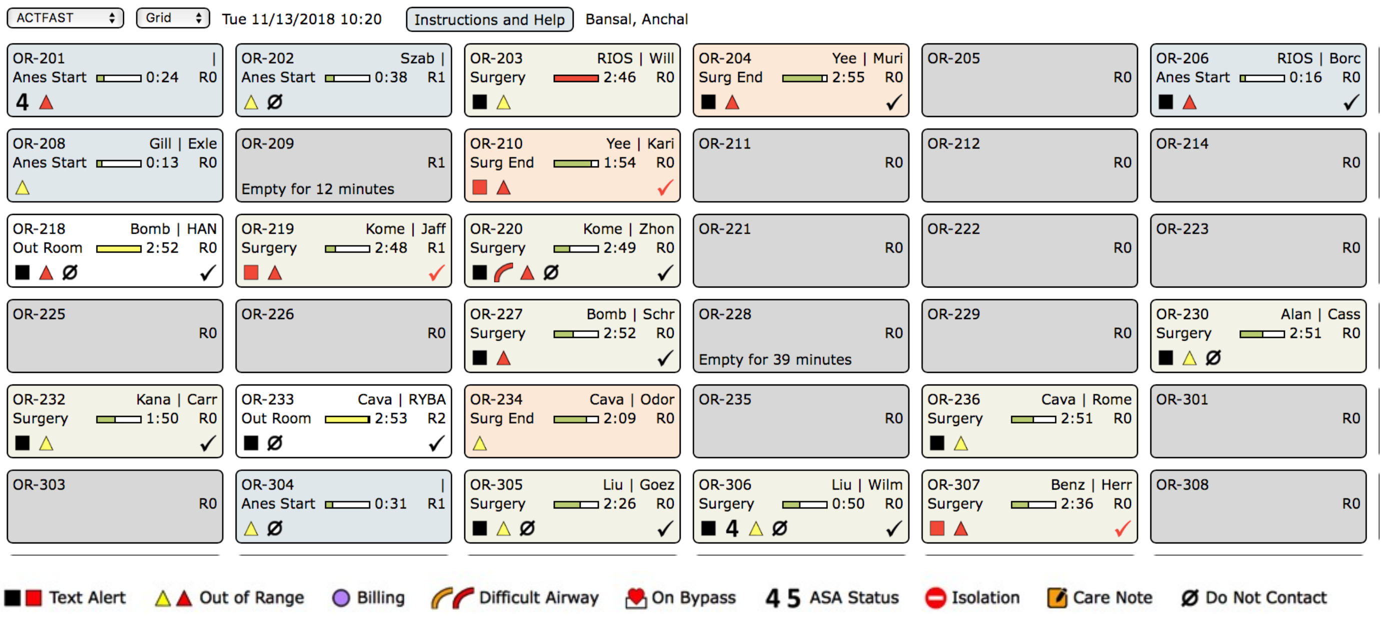

**Figure 1. Census view of AlertWatch dashboard/grid view:** This is a visual summary of all active cases. Alerts are indicated by squares and triangles. Triangles indicate that a lab or physiologic parameter is out of range. Squares indicate that a text alert is active within the patient display on the right hand side of the screen. Any time a new, actionable alert appears in a room, a ***red or black checkmark will appear in that OR.*** In the top left corner, a drop down box allows you to change the view from “Grid” to “Alert.” The Alert view provides details of alerts (Figure 2).

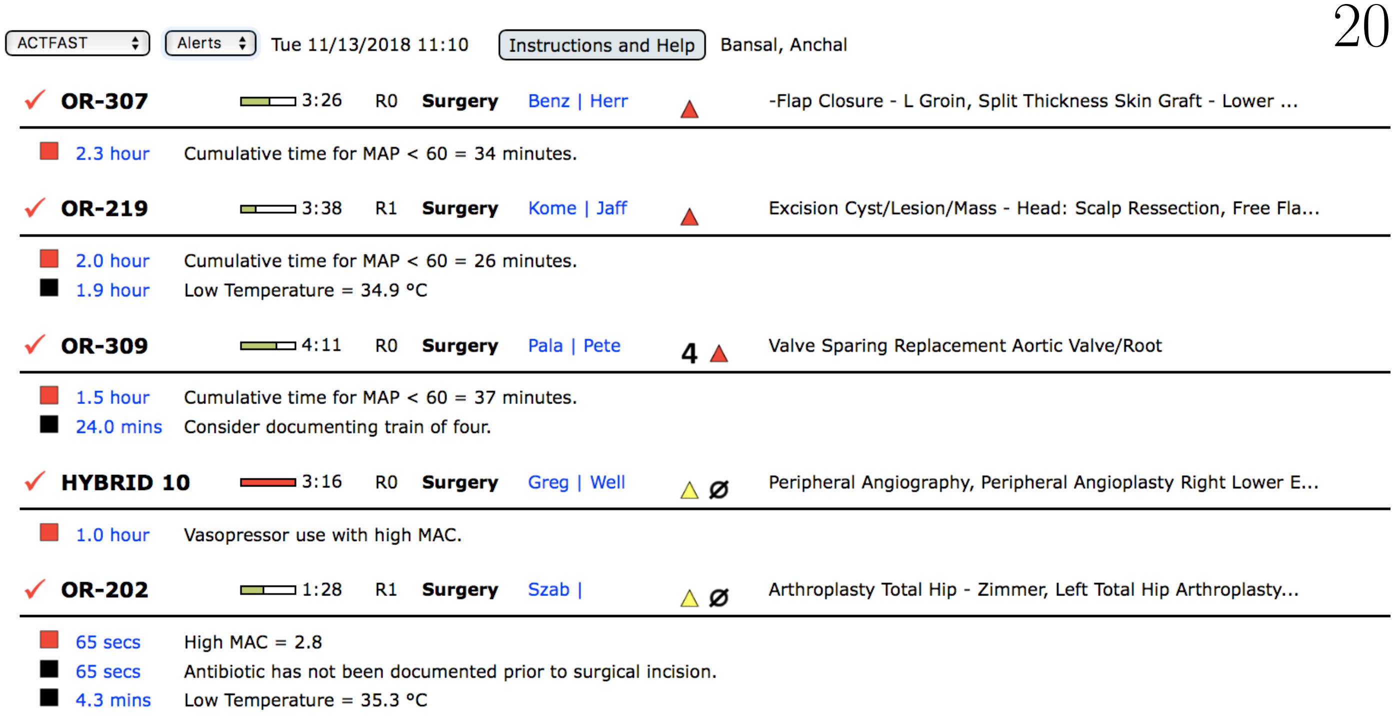

**Figure 2. "Alert" view of AlertWatch dashboard:** This view lists all current active alerts by OR. It is obtained by selecting "Alerts" from the drop down box in the top left corner of the Census view.

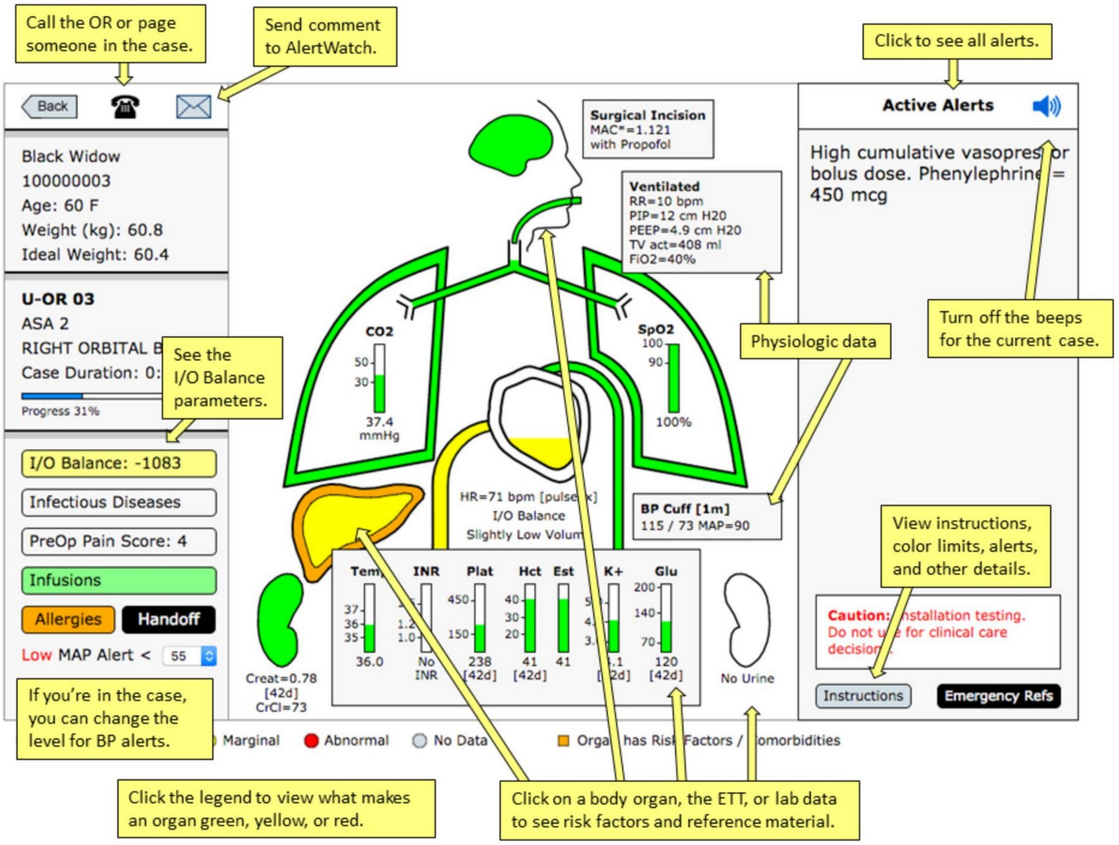

**Figure 3a.**

**Patient Display Overview**

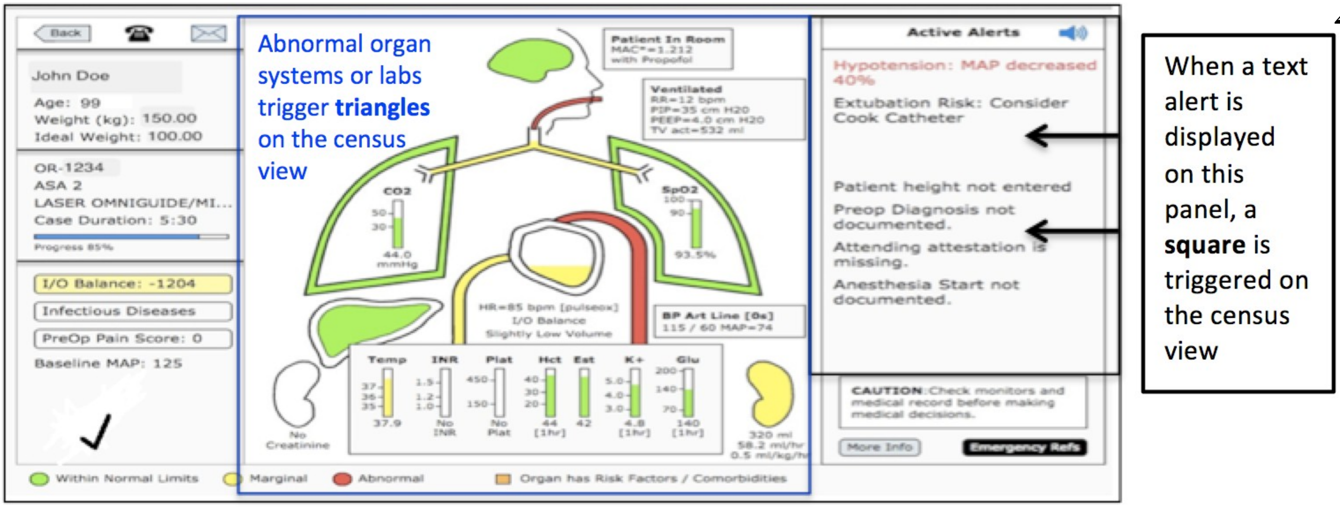

**Figure 3b. Patient Display:** Clicking on an OR accesses this screen. The case information is listed on the left side of the screen and active alerts on the right. Information on comorbidities is conveyed by the *organ outline*, which is orange if there are significant risk factors. The current organ status is conveyed by the *fill of the organ*. Any abnormal lab or physiologic parameters *will trigger a black or red triangle* on the census view. Any active text alerts on the right hand panel *will trigger a black or red square* on the census view.

| | | | |
|---|---|---|---|
| **Room** | OR-201 | **Diagnosis** | Stenosis,Cervical,; |
| **Age** | | **Procedure** | Spinal Fusion Posterior Lumbar / Thoracic W/ Instrumentation/Cervical Posterior Decompression / Laminectomy/Bone Graft W/ Bone Morphogenic Protein/Spinal Cord Monitoring/Cervical Posterior Decompression / Laminectomy |
| **Gender** | F | **ASA** | ASA 3 |
| **Weight** | 75.0 | **Surgeon** | |
| **BMI** | 31.2 | **Anesthesiologist** | |
| **Surgery Date** | | **Resident/CRNA** | |

| **History / Comorbidities** | | | |
|---|---|---|---|
| Asthma | COPD | Pulmonary Embolism | Sleep Apnea |
| Type 2 Diabetes | AdmitPatient | Outpatient | Dysrhythmia |
| Mitral Valve Prolapse | Murmur | | |
| **Allergies** | | | |
| No known allergies | | | |
| **Lab Results** | | | |
| **INR** No INR | **Platelet** No Plat | **Hct** No Hct | **K+** No K+ |
| **Glucose** 140 [2hr] | **Creatinine** No Creatinine | **CrCl** N/A | **Albumin** N/A |
| **Lipase** N/A | **Amylase** N/A | **ALT** N/A | **SGOT** N/A |
| **ALP** N/A | **GGT** N/A | | |
| **Medications / Infusions** | | | |
| Phenylephrine 0.9 mcg/kg/min | | | |
| **Alerts** | | | |
| H&P incomplete. ASA Status, Airway Exam [inactive, count=1] | Attending has not signed in. [inactive, count=1] | Hypotension: MAP = 56 [active, count=4] | High cumulative vasopressor bolus dose. Phenylephrine = 1400 mcg [active, count=1] |
| Hypotension: MAP = 49 [inactive, count=1] | Hypotension: MAP = 54 [inactive, count=2] | Cumulative time for MAP < 60 = 26 minutes. [active, count=1] | |

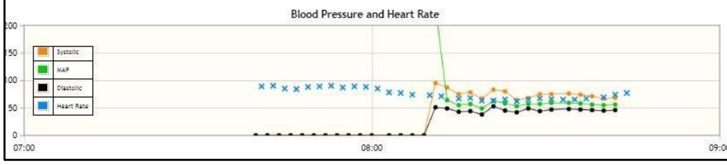

**Figure 4. Handoff View:** This view gives a summary of patient demographics, co-morbidities, recent labs, infusions, ins and outs, active and inactive alerts, and trend of vital signs throughout the case

**ACTFAST: Case Review** — **Treatment Patient: Contact Providers** — Close

| When ▼ | Organ ▼ | Alert/Color ▼ | Assessment/Action | |
|---|---|---|---|---|
| ✔ | Lungs | **Alert**. PEEP = 3 | + Add Assessment | No Worries |
| ✔ | Lungs | **Level**. SpO2 = 89.7 | + Add Assessment | No Worries |
| 09:09 | Heart | **Alert**. Hypotension: MAP = 56 | **Assessment:** Significant **Action:** Add fluid if patient is dry, Treat with vasopressor (bolus or infusion) **Comment:** | |

**Figure 6a. Case Review:** This view is obtained by clicking on the checkmark at the bottom left of the Patient Display. New alerts are indicated by red and black checkmarks. Clicking on “add assessment” brings up a check box menu.

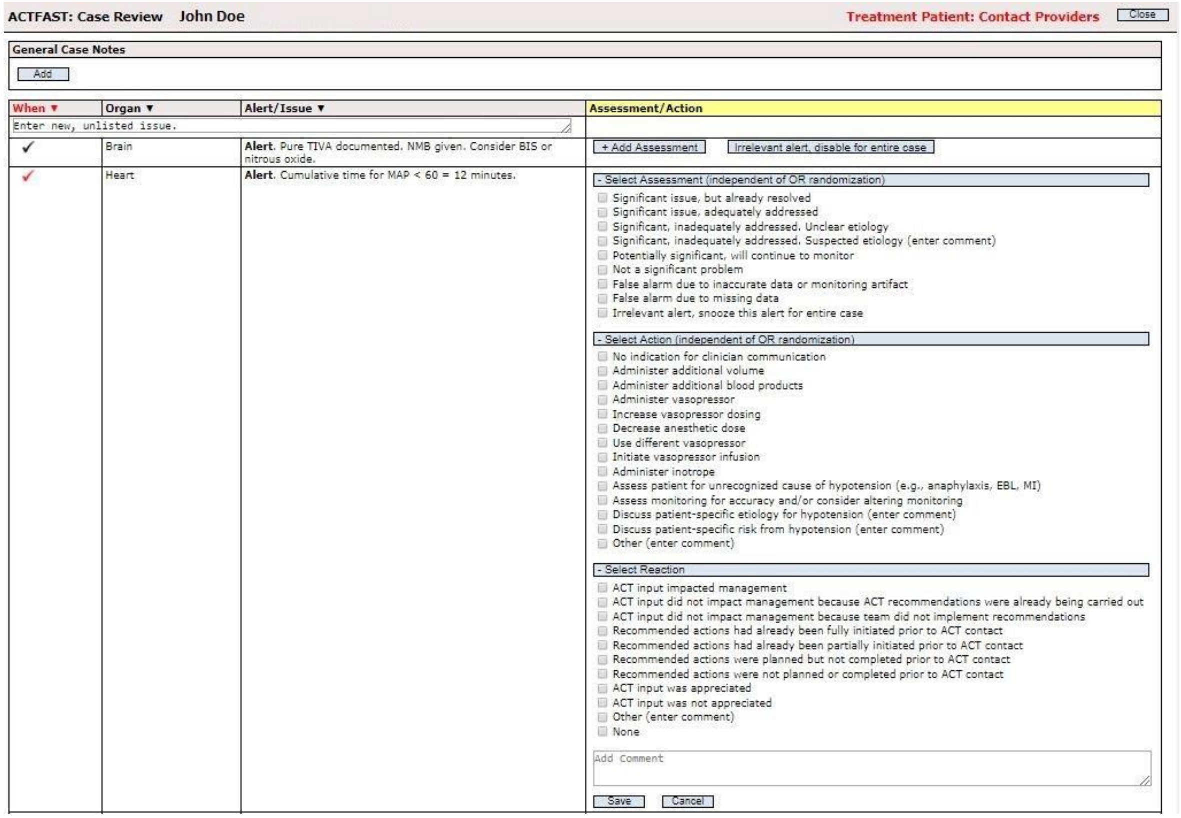

**Figure 6b. Case Review alert menu:** This is where you will document your assessments, actions, and comments. **Once you save your work, the checkmark in the far left column will become a timestamp. If you save an assessment for all active alerts for a room, the checkmark on the census view will turn green.** Otherwise the red or black checkmark will remain. It will also reappear any time there is a new alert. Orange checkmarks indicate which rooms have alerts that require additional follow up (i.e. assessment/action has been entered, but reaction is pending).

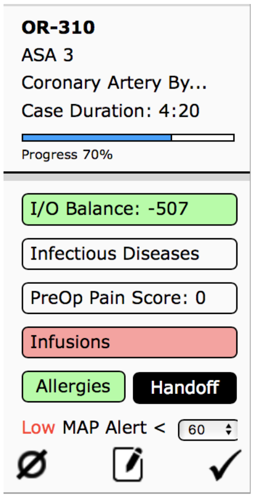

**Figure 7. Note pads**: Note pad may be clear, yellow, green or orange. Clear note pad indicates no note is written, yellow indicates a patient specific note/reminder in the alert assessment written by the telemedicine team [shows in Alert and Review view]., green indicates intraoperative quicknotes documented in Epic, and orange identifies patients with specific increased risk (Ex: HOCM, pulmonary hypertension, type 1 diabetes, etc…). Having an orange notepad increases your risk score by 100, and shifts these patients to

the top of the review view. . Clicking on the note brings up the tower note view where a new note can be started or an existing note can be viewed or modified.

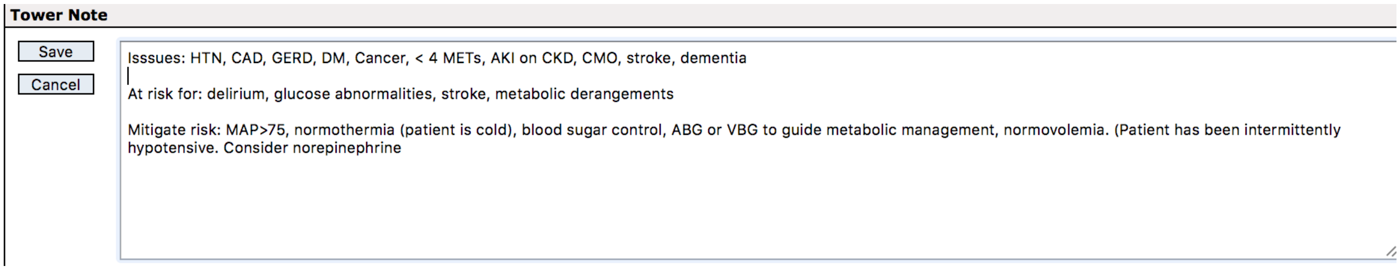

**Figure 8. Tower Note:** This view is found by clicking on "Add" in the top left corner of "Case Review" View (fig 4b) or by clicking on the notepad in the bottom left corner of the "Patient View" (fig 5) A short note should be written outlining Issues, potential problems, and strategies for risk mitigation. Make sure to click "Save" before exiting the view or your note will disappear. This note will then appear on the "Alert View" below any alerts.

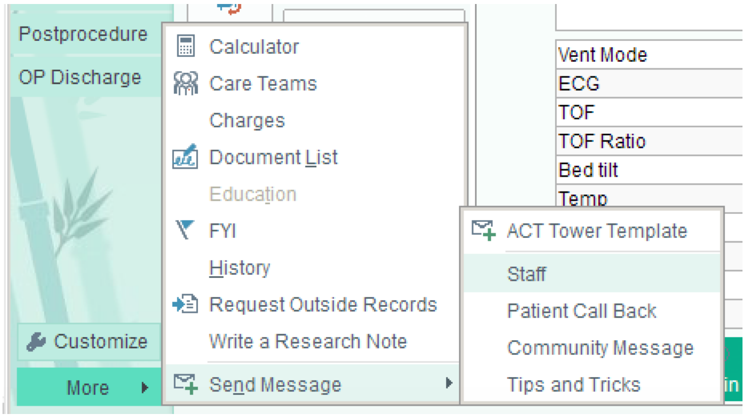

**Figure 9a. Accessing messaging in Epic:** Epic messages are NOT part of the medical record

Epic Secure Messaging

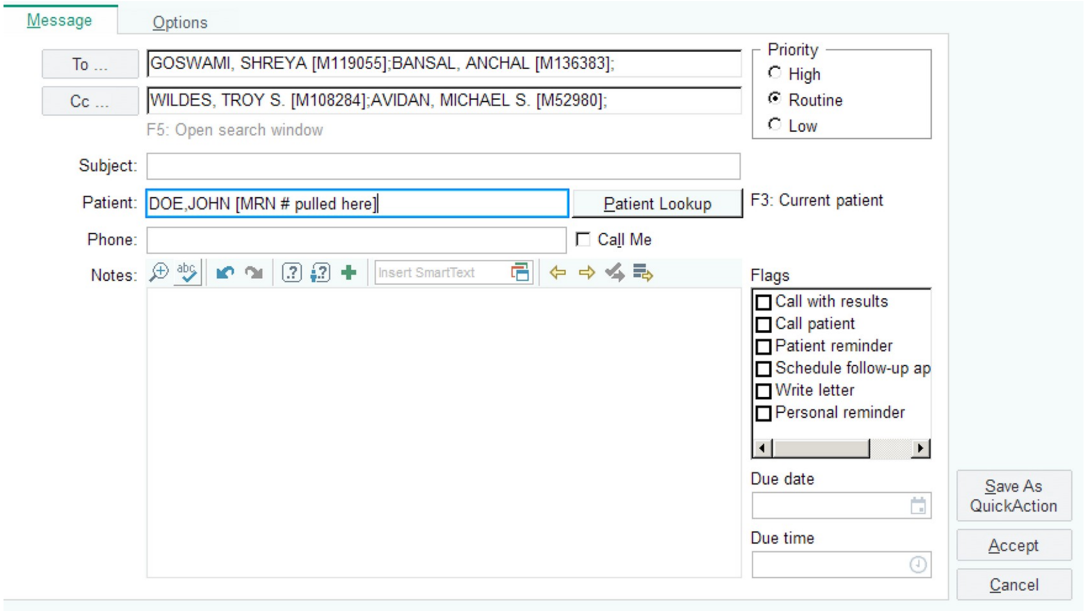

**Figure 9b. Epic Messaging Platform:** This is what the messaging platform will look like. Smart text can be used to populate the note using the phrase .actow__ if desired, or a free text can be entered. Send message to OR clinicians and cc ACT clinicians. Click “Accept” to send the message

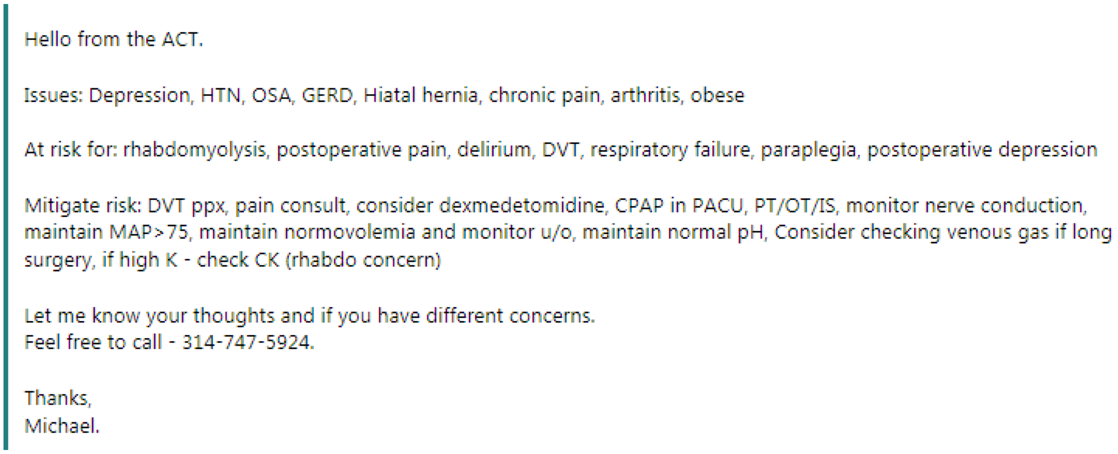
Hello from the ACT.

Issues: Depression, HTN, OSA, GERD, Hiatal hernia, chronic pain, arthritis, obese

At risk for: rhabdomyolysis, postoperative pain, delirium, DVT, respiratory failure, paraplegia, postoperative depression

Mitigate risk: DVT ppx, pain consult, consider dexmedetomidine, CPAP in PACU, PT/OT/IS, monitor nerve conduction, maintain MAP>75, maintain normovolemia and monitor u/o, maintain normal pH, Consider checking venous gas if long surgery, if high K - check CK (rhabdo concern)

Let me know your thoughts and if you have different concerns.
Feel free to call - 314-747-5924.

Thanks,
Michael.

**Figure 10. Example Epic Note:** This is an example of a free text note sent in the epic environment addressing general areas of concern for the patient. Make sure to address the note to everyone in the OR as well as the ACT clinicians and to indicate at the bottom that you would like a response. If no response in received in ~15 minutes, call one of the OR clinicians and ask them to address the message when they have time.

| **Glossary of symbols** | |
|---|---|
| | Isolation |
| | High-risk comorbidity (ie. VonWillebrandt’s, history of malignant hyperthermia, etc) |
| | On bypass |

| | |
|---|---|
| 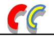 | Difficult airway, or risk factors for difficult airway |
| **4/5** | ASA 4/5 |
| □ | Organ system/lab is yellow |
| □ | Organ system/lab is red |
| ■ | Black text alert active in right hand panel of Patient Display |
| ■ | Red text alert in right hand panel of Patient Display |
| ✓ | New black alert since last alert addressed or since beginning of case |
| ✓ | Alert has been partially addressed, but “Reaction” is pending |
| ✓ | New red alert since last alert addressed or since beginning of case |
| ✓ | Room has had at least one alert that has been addressed, without subsequent alerts |
| Ø | Do not contact |

| **OR Colors** | |
|---|---|
| Blue | Patient is in the room |
| Tan | Case is in progress (incision has been made) |
| Dark gray | Room is empty |

#### GETTING STARTED

The purpose of the Anesthesiology Control Tower is to ***use AlertWatch and supplementary data sources (Epic) to monitor active ORs on South Campus and Parkview Tower to proactively address foreseeable issues and respond to AlertWatch alerts.*** *There are 58 ORs total randomized every morning to be either contact (29 rooms) or non-contact (29 rooms).*

Daily outline (ACT runs 0700-1600)

- Start of day: 0700
    - Load all programs.
        - **AlertWatch:** Username and password are same as Epic credentials. https://bjcalwaweb01t.bjc-nt.bjc.org/#/login
        - **Epic:** Epic storefront link on desktop or through intranet.
    - Refer to the roles and responsibilities documents to ensure understanding of scope of work in the tower.
    - Refer to protocol library for questions concerning alert triggers, thresholds for response, appropriate timing and modes of response, and other best practices (under development).
- 0700-1600
    - Monitor all cases on AlertWatch-ACT census (South Campus and Parkview Tower), complete case reviews (indicated by grey diamond), and address alerts (indicated by red/black checkmarks).
    - **Case Reviews:** Clinicians should prioritize patients with higher risk scores, and ideally earlier in case progression, that have been randomized to a contact room. All cases with an orange note pad alert should be reviewed as early as possible, though occasionally the orange note pad alert factor may be discovered to be less worrisome.
        - Clinicians will prepare cases as "Tower Notes" by completing case assessments (grey diamond in patient display).
        - After quick discussion/huddle of review with attending is complete, tower clinicians can message OR clinicians a short summary of pre-emptive recommendations.
        - When relevant and deemed to be helpful, identify useful protocols/literature and provide this information to OR clinicians.
        - <u>Notepads</u>
            - 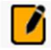 (orange): Identifies patients with specific increased risk (Ex: HOCM, pulmonary hypertension, type 1 diabetes, etc…). Having an orange notepad increases your risk score by 100, and shifts these patients to the top of the review view.
            - 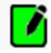 (green): Indicates intraoperative quicknotes documented in Epic.

            - 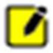 (yellow): Indicates a patient specific note/reminder in the alert assessment written by the telemedicine team [shows in Alert and Review view].

    - **Alerts:** Clinicians should address all alerts, irrespective of randomization status.
        - Any time that a new actionable alert is triggered, a red or black checkmark appears for the OR in which it is triggered. These checkmarks will remain until the alert is addressed by the ACT or resolve on their own. Once addressed, the red or black checkmark will be replaced by a green checkmark.
        - The alert is "addressed" by clicking on the check mark while in the Patient Display (bottom left-hand corner in patient display) and completing and saving a response for the specific alert.

    - **Contacting Rooms:** Epic Secure Chat, Epic Staff Messaging system, phone calls.
        - Rooms are primarily contacted through Epic Secure Chat (not part of the medical record).
            - If there is no response from OR clinicians or indication they have read the message after ~15mins, call to confirm receipt of message.
        - Epic messaging system (not part of medical record).
            - Smart-phrases are available for full discussion recommendations and common alerts. Smart-phrase titles begin with ' .actow__ '.
            - If there is no response from OR clinicians after ~15mins call to confirm receipt of message.
            - Smart-phrases can be activated within a temporary staff message to leverage auto-generated content, and then pasted into a SecureChat conversation (which is more user friendly to read/respond)
        - Okay to use calling as primary mode of contact if preferred. Calling is always necessary for urgent messages.
        - Non-contact rooms should not be contacted unless marked concern for patient safety that justifies a protocol deviation. If a non-contact room is contacted, follow tower instructions for documentation of protocol break.
- Education
    - Attending will prepare and conduct one core topic discussion (5-10 minutes) from the core topic library (under development).

    - CRNA, SRNA, and Resident will prepare and conduct one mini topic discussion (1-2 minutes) from the mini topic library (under development).

#### Figures:

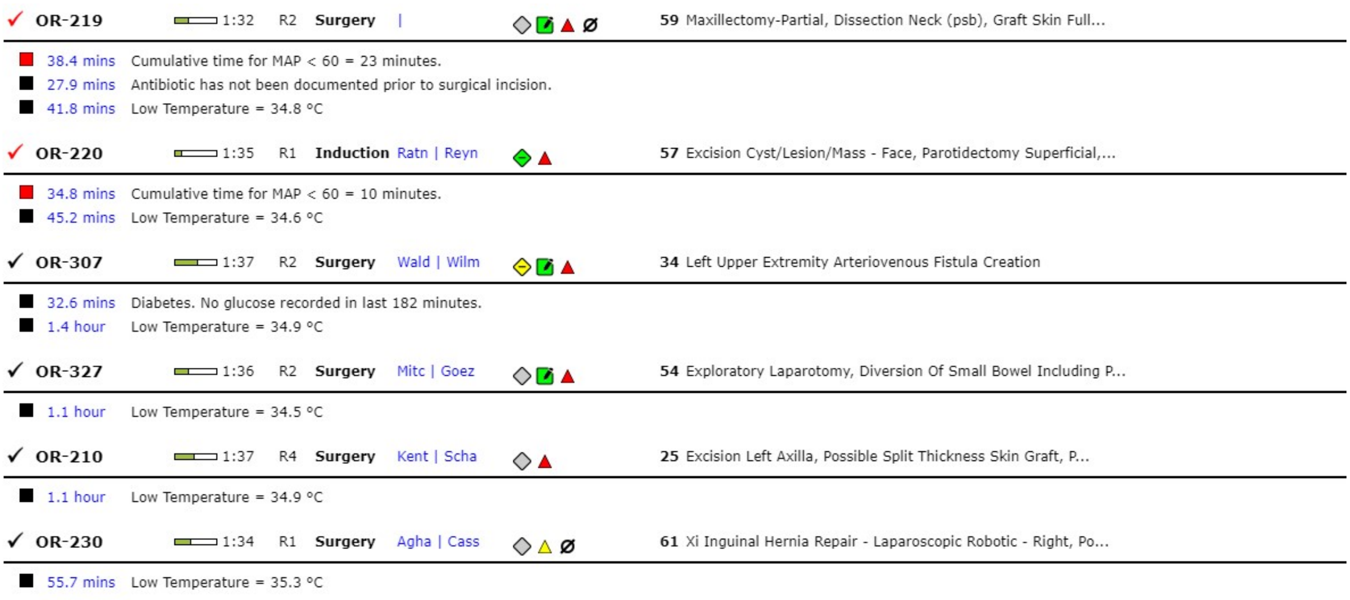

**Figure 1.** "Alert" view of AlertWatch dashboard displaying rooms in order of alert urgency. The room assignment as contact/non-contact is also displayed here just before the name of the case.

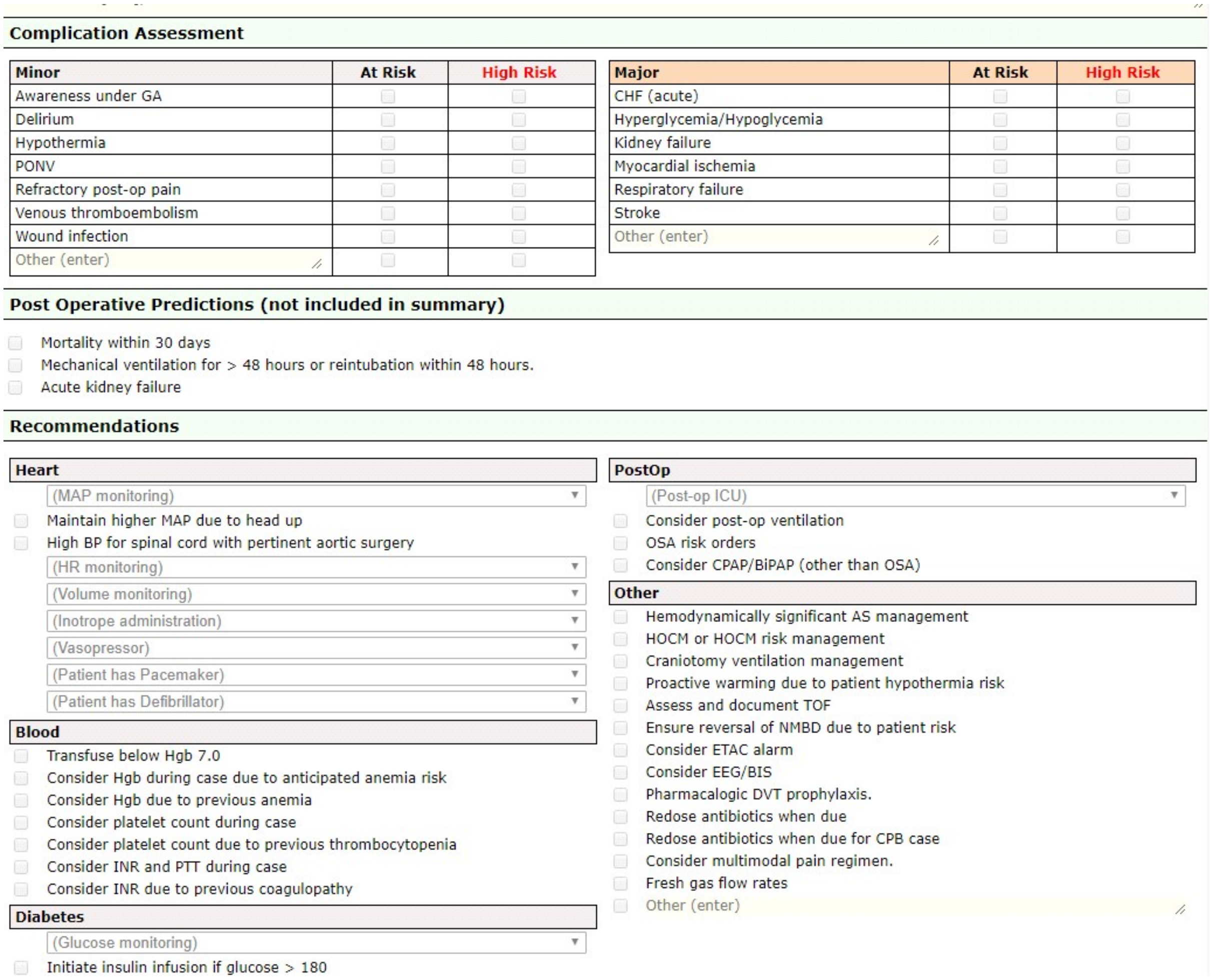

**Complication Assessment**

| Minor | At Risk | High Risk |
|---|---|---|
| Awareness under GA | | |
| Delirium | | |
| Hypothermia | | |
| PONV | | |
| Refractory post-op pain | | |
| Venous thromboembolism | | |
| Wound infection | | |
| Other (enter) | | |

| Major | At Risk | High Risk |
|---|---|---|
| CHF (acute) | | |
| Hyperglycemia/Hypoglycemia | | |
| Kidney failure | | |
| Myocardial ischemia | | |
| Respiratory failure | | |
| Stroke | | |
| Other (enter) | | |

**Post Operative Predictions (not included in summary)**

- Mortality within 30 days
- Mechanical ventilation for > 48 hours or reintubation within 48 hours.
- Acute kidney failure

**Recommendations**

**Heart**

(MAP monitoring)
- Maintain higher MAP due to head up
- High BP for spinal cord with pertinent aortic surgery

(HR monitoring)
(Volume monitoring)
(Inotrope administration)
(Vasopressor)
(Patient has Pacemaker)
(Patient has Defibrillator)

**Blood**

- Transfuse below Hgb 7.0
- Consider Hgb during case due to anticipated anemia risk
- Consider Hgb due to previous anemia
- Consider platelet count during case
- Consider platelet count due to previous thrombocytopenia
- Consider INR and PTT during case
- Consider INR due to previous coagulopathy

**Diabetes**

(Glucose monitoring)
- Initiate insulin infusion if glucose > 180

**PostOp**

(Post-op ICU)
- Consider post-op ventilation
- OSA risk orders
- Consider CPAP/BiPAP (other than OSA)

**Other**

- Hemodynamically significant AS management
- HOCM or HOCM risk management
- Craniotomy ventilation management
- Proactive warming due to patient hypothermia risk
- Assess and document TOF
- Ensure reversal of NMBD due to patient risk
- Consider ETAC alarm
- Consider EEG/BIS
- Pharmacalogic DVT prophylaxis.
- Redose antibiotics when due
- Redose antibiotics when due for CPB case
- Consider multimodal pain regimen.
- Fresh gas flow rates
- Other (enter)

**Figure 2:** Case Review form in Alert Watch. Case reviews can be edited to reflect when a room has been contacted. Case Review tower notes can be sent using Epic Secure Chat, or Epic messaging system with smart phrases starting with ‘ .actow__ ‘.

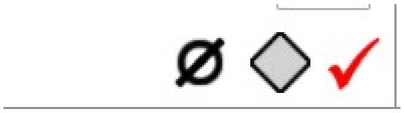

**Figure 3:** Symbols in Patient Display showing a non-contact room (circle with line thru), case review that has not been completed (grey diamond), and active alert (red or black checkmark).

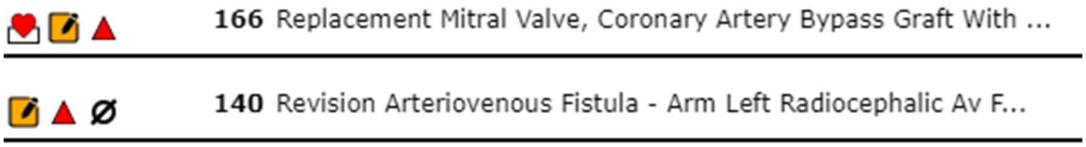

**Figure 4:** Showing numerical “risk score,” located in Alert and Review views.

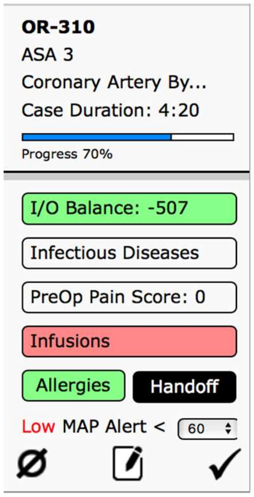

**Figure 5:** Notepad icon in the patient display view

### ACT Attending Responsibilities

#### Primary Clinical

- Keeps track of all important alerts or clinical situations that require follow-up, determines an appropriate follow up time, and ensures follow-up takes place. Best practice is to track pending follow-ups on the white board. White board notations can be delegated or personally carried out.
- If there are only two clinicians in the ACT (including the attending), the attending will routinely complete comprehensive case reviews like all other ACT clinicians. If there are 3 or more total clinicians in the ACT, the attending will generally not complete many or any comprehensive reviews themselves (due to oversight needs).

#### Oversight

- Ensures every comprehensive case review is vetted by the attending before it is sent to the OR clinician. In particular, risk categorizations and recommendations should be verified. Discussions about case reviews should, on average, take three minutes or less.

#### Vigilance

- Monitors “Alert view” for urgent or important alerts. Ensures that urgent alerts are addressed in a timely fashion by assigning to a control tower clinician or personally reviewing.

#### Teaching

- Prepares and conducts educational core topic presentation and discussion (Until the core topic library has been developed, a “mini topic” will be presented)
- Complete evaluations of Residents (under development)

#### Community

- Promotes a stimulating educational environment
- Promotes a collaborative culture of safety with OR clinicians. Ensures communications with OR clinicians are collaborative, clear, and contain sound recommendations. Personally carries out conversations with OR clinicians whenever necessary to promote quality patient care.

### ACT Clinician Responsibilities

#### Primary Clincial

- Completes seven or more comprehensive case reviews per shift. Presents every comprehensive case review to the attending. Discussions about case reviews should, on average, take three minutes or less.
- Completes alert reviews and follow-up in appropriate intervals (appropriate intervals determined by attending)
- Utilizes protocols to ensure appropriate steps are taken for each alert

#### Teaching

- Prepares and conducts educational mini topic presentation and discussion (1-2 minutes)
- CRNA completes evaluations of SRNA (under development)

#### Community

- Promotes a stimulating educational environment
- Promotes a collaborative culture of safety with OR clinicians. Ensures communications with OR clinicians are collaborative, clear, and contain sound recommendations.

*****Includes all clinicians aside from the attending and researcher (i.e. CRNA, Resident, SRNA, Researcher II, etc.)***

Approved by Dr. Troy Wildes, Dr. Mark Willingham, Dr. Daniel Helsten, Bernadette Henrichs, Brian Torres, and Melissa Milbrandt

### ACT Research Responsibilities

#### Team Support

- Provides tech and educational support as needed
- Utilizes protocols to make recommendations to other clinicians on modes of response and timing of response to alerts.
- Posts and updates protocols and ACT team metrics (under-development)
- Escalates any issues identified during the end of day debrief

#### Primary Clinical

- Completes comprehensive case reviews. Presents every comprehensive case review to the attending. Discussions about case reviews should, on average, take three minutes or less.
- Completes alert reviews and follow-up in appropriate intervals (appropriate intervals determined by attending)
- Utilizes protocols to ensure appropriate steps are taken for each alert

#### Community

- Promotes a stimulating educational environment
- Promotes a collaborative culture of safety with OR clinicians. Ensures communications with OR clinicians are collaborative, clear, and contain sound recommendations.

***** If there are two or more researchers, one researcher will take on this role and all others will default to the clinician role.***

#### SUPPLEMENTAL RESULTS

**Sup Figure 1: AlertWatch:OR interface.** Additional examples in manual of procedures.

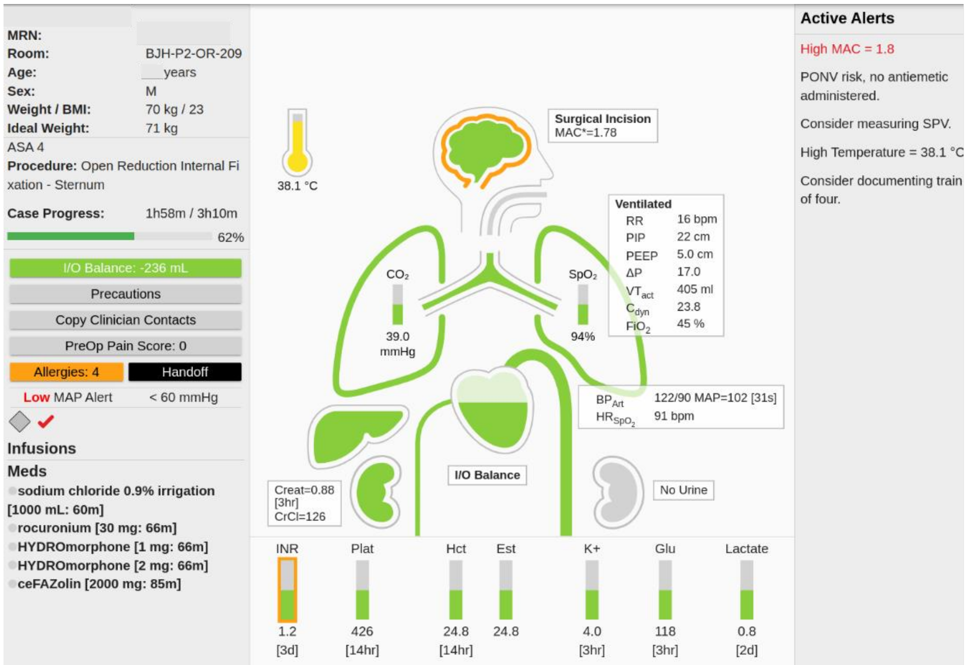

**Sup Figure 2: Machine learning risk web-app interface example with identifying information removed.**

| Room Name | Description | akigr1 | cardiac | death_in_30 | DVT_PE | ICU | post_dialysis |
|---|---|---|---|---|---|---|---|
| BJH OR POD 2 ROOM 209 | ###LAPAROSCOPIC CHOLECYSTECTOMY | 2.90% | 1.60% | 0.10% | 8.80% | 5.20% | 0.10% |
| BJH OR POD 3 ROOM 301 | EXPLORATION LIVER TRANSPLANT WITH WASHOUT LIVER BIOPSY AND BILE DUCT REVISION | 67.10% | 6.90% | 7.60% | 1.20% | 93.40% | 0.90% |
| BJH OR POD 2 ROOM 211 | TRACHEOSTOMY PERCUTANEOUS | 40.30% | 2.70% | 3.50% | 7.40% | 94.50% | 0.20% |

**Sup Fig 3: Enrollment by month.** Treatment 0 = usual care, 1 = ACT

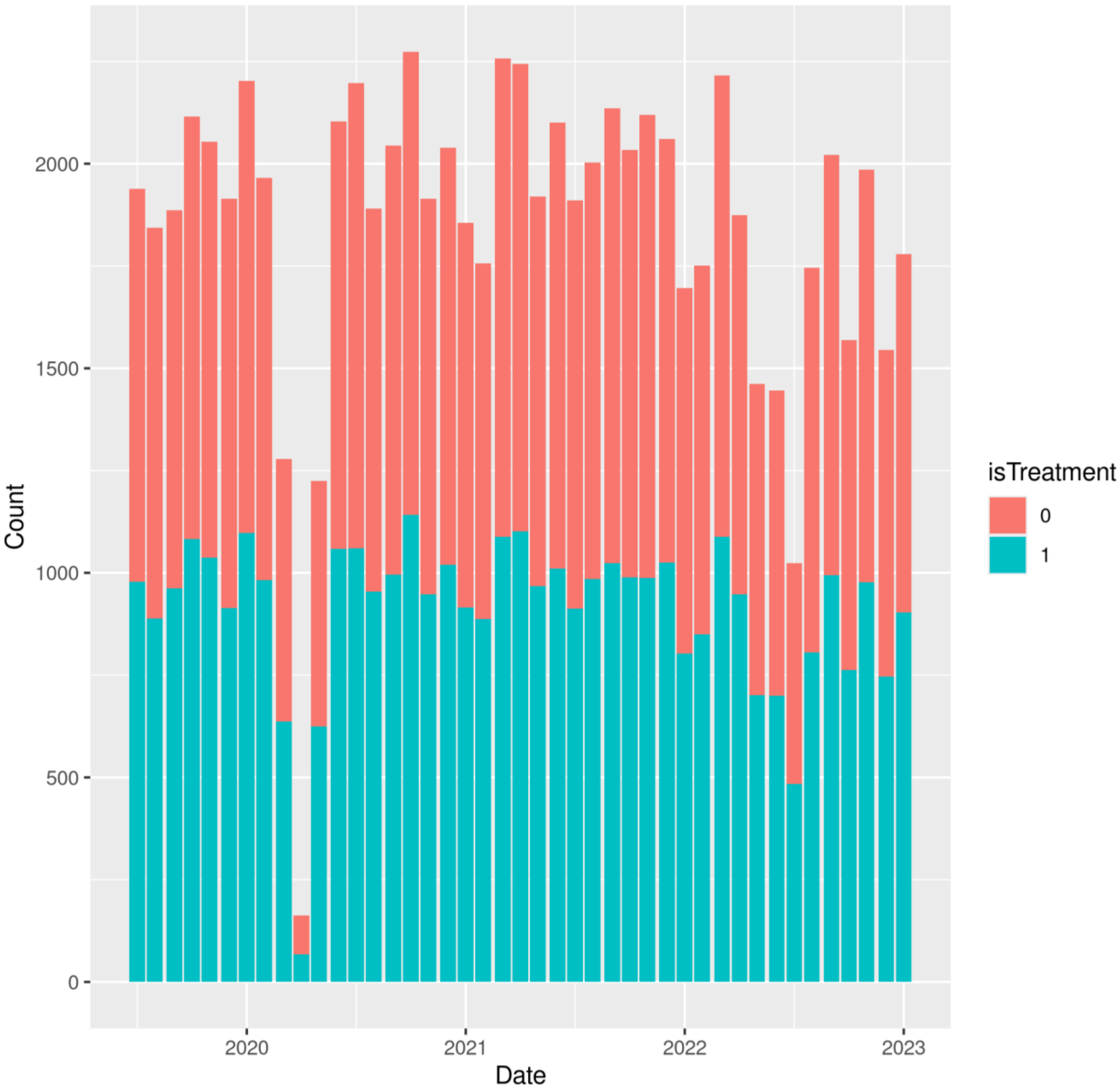

**Sup Fig 4: Primary outcomes ascertainment rate by Month.** Green = overlap, red or blue = additional ascertainment in usual care or ACT group. Ascertainment of mortality 100% by assumption (missingness is unknown). Treatment 0 = usual care, 1 = ACT

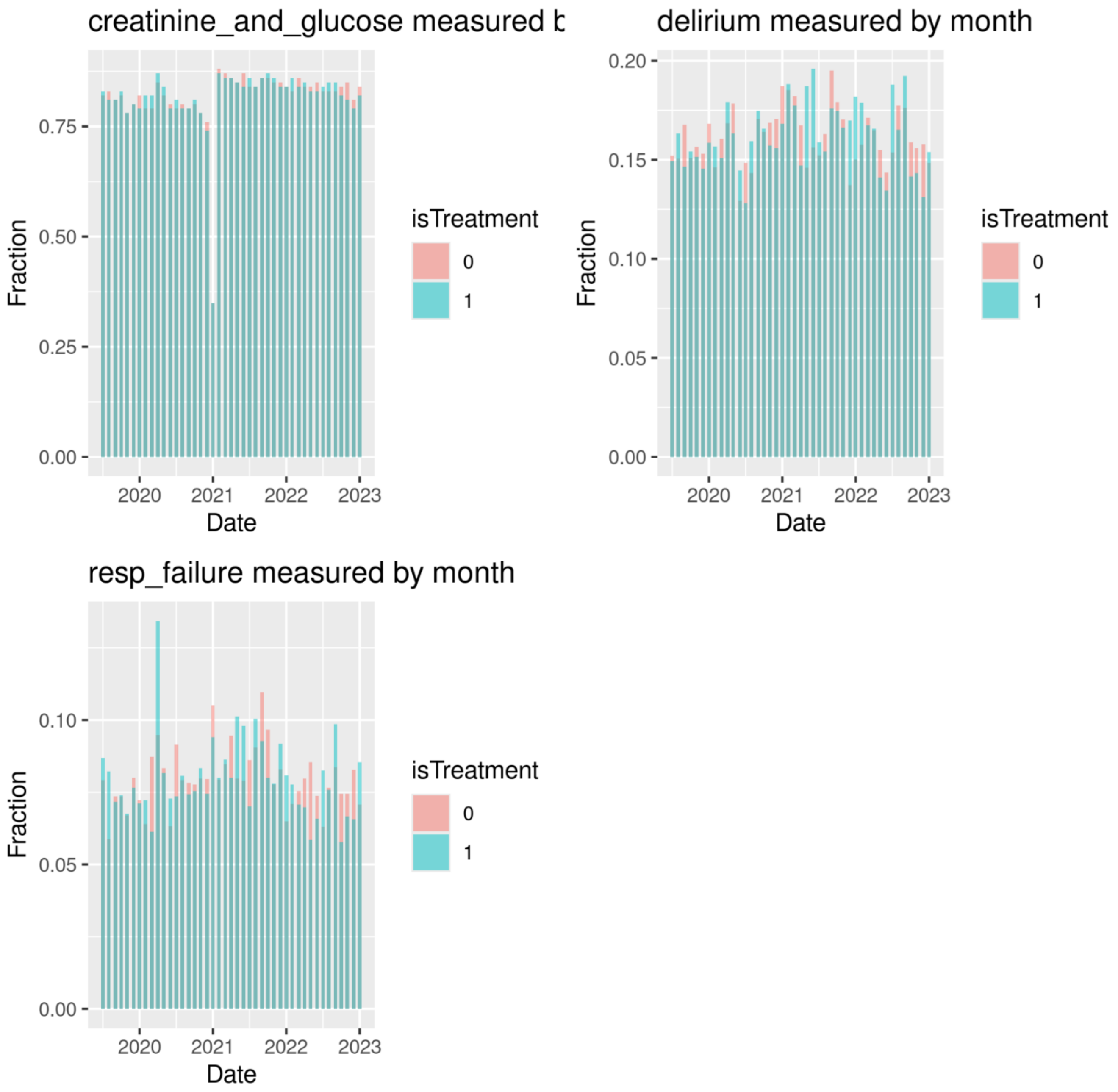

**Sup Fig 5: Secondary outcomes ascertainment rate by Month.** Green = overlap, red or blue = additional ascertainment in usual care or ACT group. Antibiotic redosing ascertainment among all cases, missing when no antibiotic redosing indicated. Efficient fresh gas flow missing when no volatile anesthetic administered. PIP missing when no ventilated time. Treatment 0 = usual care, 1 = ACT

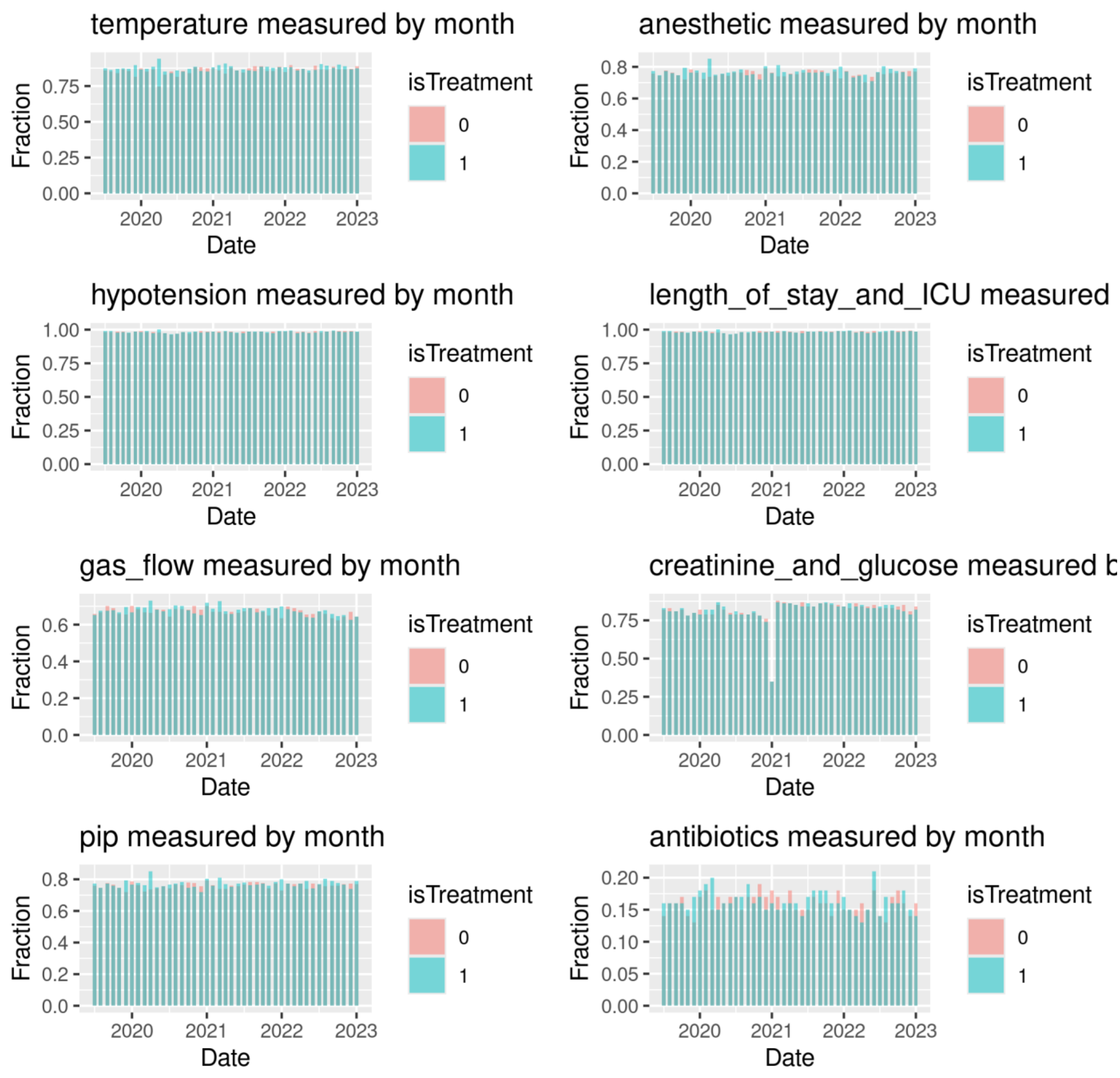

**Sup Fig 6: Covid-19 infection rates per 100k in St Louis County, Missouri (the study site) during the study**

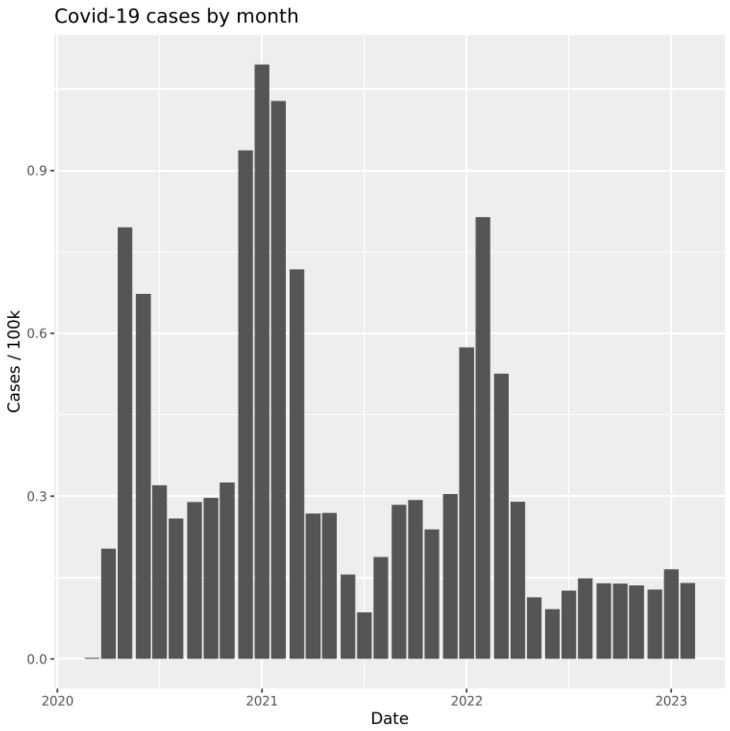

**Sup Fig 7. Case reviews sent by month.** Including only the "late period" (see Sup Table 12), the number of case reviews documented as sent to the OR per month. Mid-year (July) decreases align with reduced staff availability as new residents are on-boarded. Case reviews notably decline in the final year of the study.

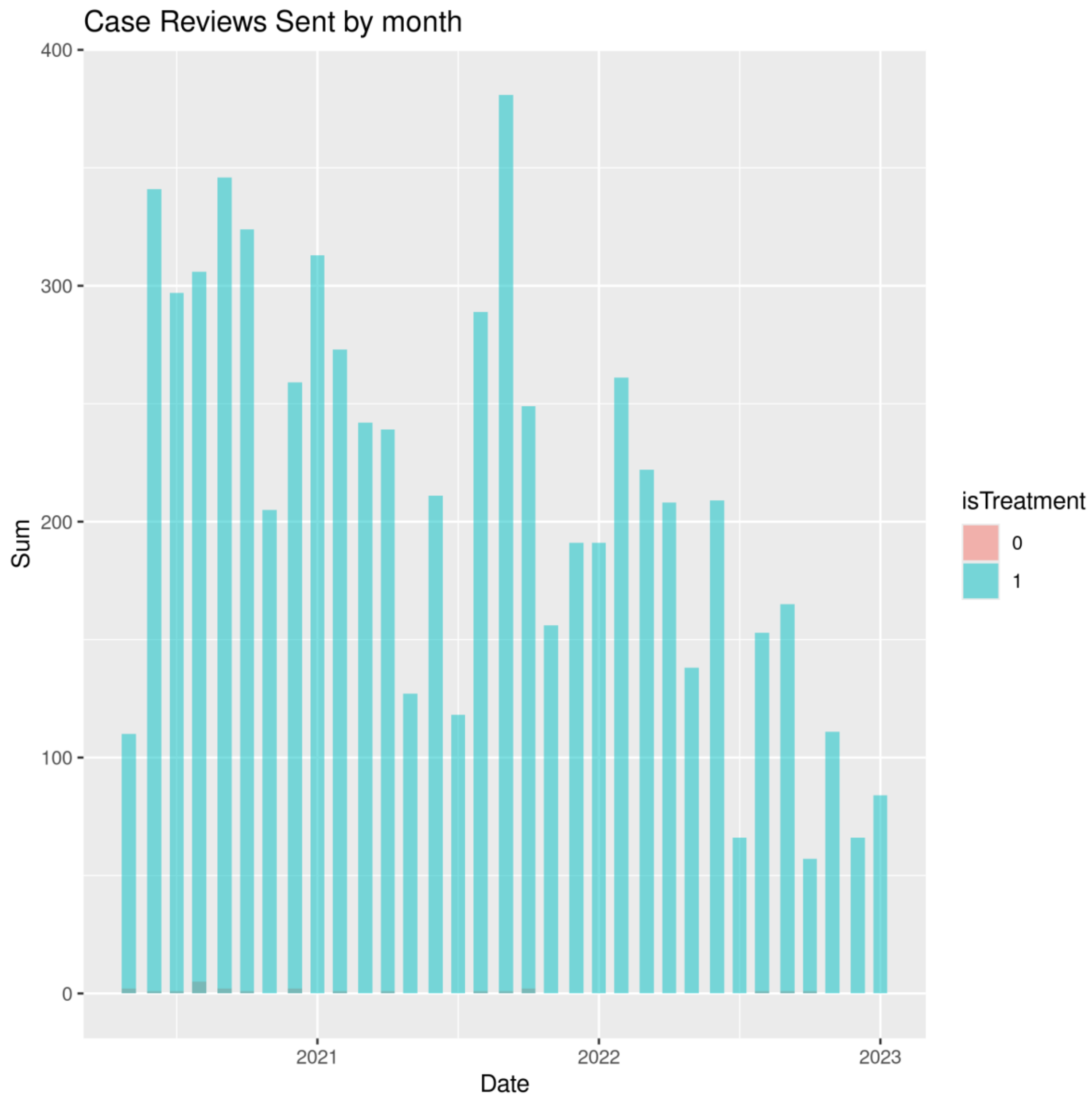

**Sup Fig 8. Alerts communicated to the operating room by month as a fraction of all acknowledged alerts.**

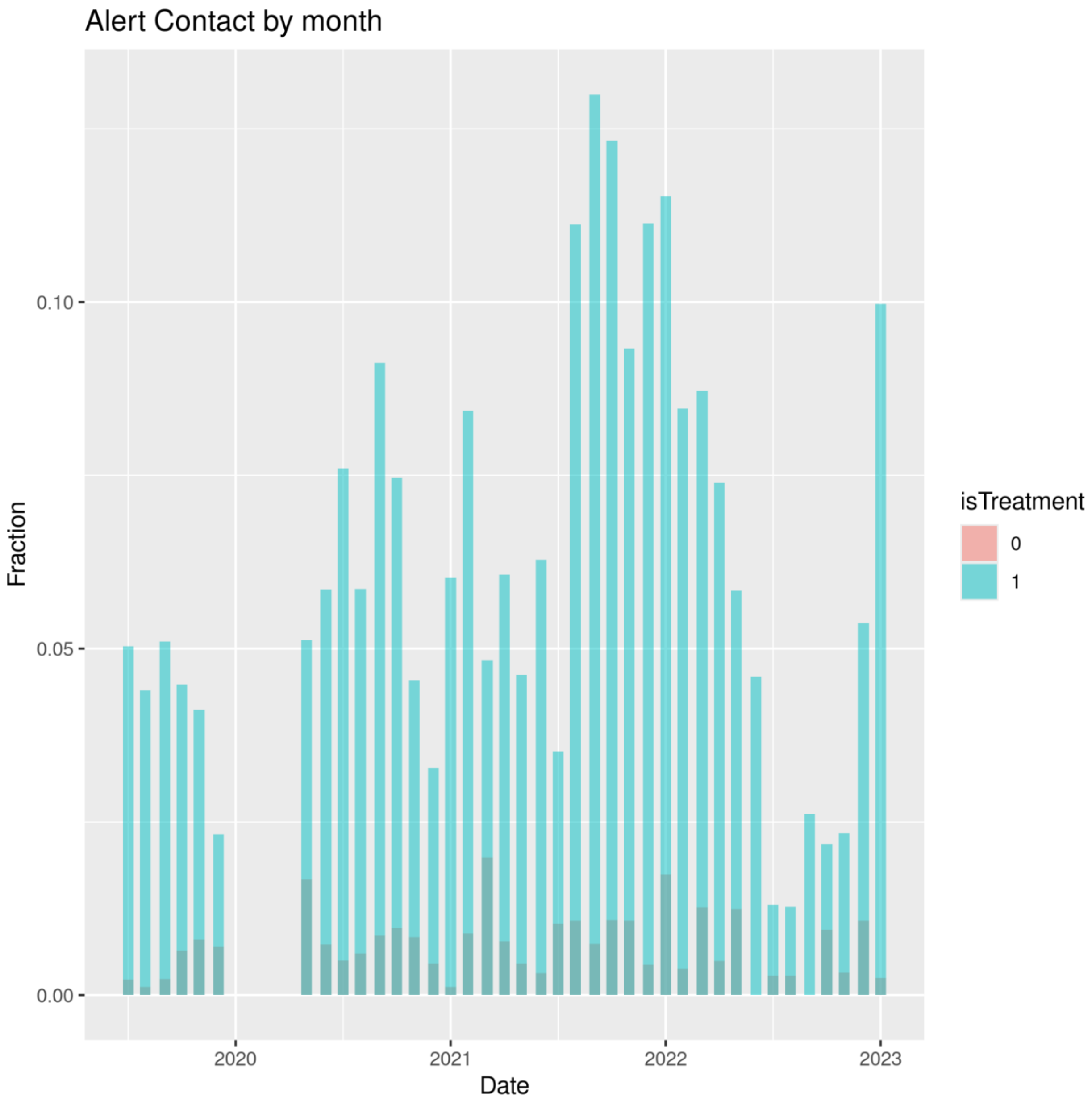

**Sup Table 1: Characteristics of reviewed vs un-reviewed intervention group cases.** Standardized diff as in Table 1

| variable | Reviewed | Unreviewed | Standardized diff |
|---|---|---|---|
| Age | 58.2 (16.2) | 56.7 (17.2) | -0.09 (-0.12 , 0.01) |
| Atrial Fibrillation | 625/6393 (9.8%) | 1541/17885 (8.6%) | 0.02 (0.00 , 0.03) |
| **Anemia** | **2020/6393 (31.6%)** | **4903/17885 (27.4%)** | **0.04 (0.03 , 0.05)** |
| **Asthma** | **858/6393 (13.4%)** | **2016/17885 (11.3%)** | **0.03 (0.02 , 0.04)** |
| **Coronary Artery Disease** | **951/6393 (14.9%)** | **2252/17885 (12.6%)** | **0.03 (0.02 , 0.04)** |
| **Cancer** | **977/6393 (15.3%)** | **1803/17885 (10.1%)** | **0.07 (0.06 , 0.08)** |
| Chronic kidney disease | 940/6393 (14.7%) | 2372/17885 (13.3%) | 0.02 (0.00 , 0.03) |
| End-stage renal disease | 293/6393 (4.6%) | 843/17885 (4.7%) | 0.00 (0.00 , 0.01) |
| **Chronic obstructive pulmonary disease** | **795/6393 (12.4%)** | **1851/17885 (10.3%)** | **0.03 (0.02 , 0.04)** |
| Dementia | 218/6393 (3.4%) | 536/17885 (3%) | 0.01 (0.00 , 0.02) |
| **Diabetes** | **1863/6393 (29.1%)** | **3968/17885 (22.2%)** | **0.07 (0.06 , 0.08)** |
| Stroke | 423/6393 (6.6%) | 1044/17885 (5.8%) | 0.01 (0.00 , 0.03) |
| **Venous thrombosis history** | **760/6393 (11.9%)** | **1751/17885 (9.8%)** | **0.03 (0.02 , 0.04)** |
| **Sleep apnea** | **1367/6393 (21.4%)** | **3024/17885 (16.9%)** | **0.05 (0.04 , 0.06)** |
| **Hypertension** | **3787/6393 (59.2%)** | **8886/17885 (49.7%)** | **0.08 (0.07 , 0.10)** |
| **Emergency status** | **79/6393 (1.2%)** | **342/17885 (1.9%)** | **0.02 (0.01 , 0.03)** |
| **Race** | | | **0.07 (0.05 , 0.08)** |
| Asian | 72/6908 (1%) | 230/20139 (1.1%) | |
| Black or African American | 1209/6908 (17.5%) | 4045/20139 (20.1%) | |
| Unknown or Other | 609/6908 (8.8%) | 2521/20139 (12.5%) | |
| White | 5018/6908 (72.6%) | 13343/20139 (66.3%) | |
| **Sex** | | | **0.03 (0.02 , 0.04)** |
| Female | 3716/6908 (53.8%) | 10167/20139 (50.5%) | |
| Male | 3190/6908 (46.2%) | 9971/20139 (49.5%) | |
| Unknown or Other | 2/6908 (0%) | 1/20139 (0%) | |
| **Functional Capacity (metabolic equivalents)** | | | **0.11 (0.10 , 0.12)** |
| <4 METs | 2160/6908 (31.3%) | 5217/20139 (25.9%) | |

| | | | |
|---|---|---|---|
| 4-6 METs | 3462/6908 (50.1%) | 9142/20139 (45.4%) | |
| 6-10 METs | 174/6908 (2.5%) | 429/20139 (2.1%) | |
| >10 METs | 34/6908 (0.5%) | 74/20139 (0.4%) | |
| Unknown or Unable to Assess | 1078/6908 (15.6%) | 5277/20139 (26.2%) | |
| **American Society of Anesthesiologists Physical Status** | | | **0.05 (0.04 , 0.06)** |
| 1 | 179/6908 (2.6%) | 816/20139 (4.1%) | |
| 2 | 2133/6908 (30.9%) | 6214/20139 (30.9%) | |
| 3 | 3205/6908 (46.4%) | 8288/20139 (41.2%) | |
| 4 | 532/6908 (7.7%) | 1747/20139 (8.7%) | |
| Unknown or Other | 859/6908 (12.4%) | 3074/20139 (15.3%) | |
| **Surgical Service** | | | **0.17 (0.16 , 0.18)** |
| Cardiothoracic | 503/6908 (7.3%) | 2113/20139 (10.5%) | |
| Colorectal | 265/6908 (3.8%) | 553/20139 (2.7%) | |
| General and Trauma | 1172/6908 (17%) | 2389/20139 (11.9%) | |
| Gynecology | 863/6908 (12.5%) | 2386/20139 (11.8%) | |
| Misc procedures, endoscopy, ophthalmology | 26/6908 (0.4%) | 1646/20139 (8.2%) | |
| Neurosurgery | 682/6908 (9.9%) | 1320/20139 (6.6%) | |
| Orthopaedics | 1302/6908 (18.8%) | 3258/20139 (16.2%) | |
| Otolaryngology | 576/6908 (8.3%) | 1732/20139 (8.6%) | |
| Plastics | 135/6908 (2%) | 536/20139 (2.7%) | |
| Transplant | 263/6908 (3.8%) | 646/20139 (3.2%) | |
| Urology | 633/6908 (9.2%) | 2205/20139 (10.9%) | |
| Vascular | 488/6908 (7.1%) | 1355/20139 (6.7%) | |

**Sup Table 2: Replies to case reviews.** The data is divided by a back-end server upgrade in May 2020. Prior to the upgrade, the case review forms were not stored with sufficient length to include whether or not the case review was sent or acknowledged by the intraoperative team; a default 256 character length in an intermediate transformation truncated the information. After the upgrade, the communication information was fully available. Based on the post-upgrade data, pre-upgrade case reviews were marked as "likely sent" if edits were saved more than twice with a greater than 5-minute separation. This additional edit and time seems to reflect the time to send a message to the intraoperative clinician and receive their response. Totals are less than the total in the study because of server downtime. Given the typical 13-15 case reviews per day, patients on days with no case reviews performed were excluded from the denominator.

| **Early period** | N in study | N reviewed | N likely sent |
|---|---|---|---|
| Telemedicine Group | 7529 | 2615 | 2333 |
| Usual Care Group | 7640 | 294 | 58 |

| **Late Period** | N in study | N reviewed | N sent | OR reply recorded |
|---|---|---|---|---|
| Telemedicine Group | 27047 | 9197 | 6913 | 3379 |
| Usual Care Group | 28279 | 1551 | 23 | 7 |

**Sup Table 3: Alternative specification analysis.** ICU length of stay truncated at 7 days; patients dying in the first 7 days assigned 7 days length of stay. Time weighted hypotension unit = mmHg * minute. Change in creatinine is ratio. PIP units cmH2O. See supplemental methods for outcome definitions.

| Outcome | Intervention | Usual care | GEE coef | p value | N total | GEE coef (99.5% CI) |
|---|---|---|---|---|---|---|
| Composite | 0.15 (0.48) | 0.15 (0.47) | -0.00 (-0.01, 0.01) | 0.82 | 71927 | -0.00 (-0.01, 0.01) |
| ICU length of stay | 1.21 (4.92) | 1.21 (4.94) | 0.01 (-0.10, 0.11) | 0.89 | 71927 | 0.01 (-0.13, 0.14) |
| Mean temperature | 36.16 (0.60) | 36.16 (0.60) | 0.00 (-0.01, 0.02) | 0.57 | 63024 | 0.00 (-0.01, 0.02) |
| Time weighted hypotension | 100.89 (342.90) | 102.85 (342.19) | -1.96 (-9.30, 5.38) | 0.46 | 70939 | -1.96 (-11.10, 7.18) |
| Change in creatinine | 1.05 (0.46) | 1.05 (0.44) | 1.00 (0.99, 1.01) | 0.86 | 42805 | 1.00 (0.99, 1.01) |
| Mean PIP | 20.55 (5.12) | 20.53 (5.13) | 0.02 (-0.12, 0.15) | 0.7 | 55479 | 0.02 (-0.15, 0.19) |
| Delirium (missing as false) | 1264/35302 (3.6) | 1298/36625 (3.5) | 1.01 (0.91, 1.13) | 0.8 | 71927 | 1.01 (0.88, 1.16) |
| Hyperglycemia: DM only | 938/7937 (11.8) | 914/8341 (11.0) | 1.08 (0.96, 1.22) | 0.08 | 16278 | 1.08 (0.93, 1.25) |
| Fraction time efficient gas flow | 0.75 (0.43) | 0.75 (0.43) | -0.00 (-0.01, 0.01) | 0.78 | 48768 | -0.00 (-0.02, 0.01) |

**Sup Table 4: Subgroup analysis: age >=65.** All other factors as in Table 2.

| Outcome | Intervention | Usual care | GEE coef (95% CI) | p value | N total | GEE coef (99.5% CI) | uncorrected p |
|---|---|---|---|---|---|---|---|
| Delirium | 621/1698 (36.6) | 634/1795 (35.3) | 1.04 (0.92, 1.16) | 0.66 | 3493 | 1.04 (0.89, 1.20) | 0.44 |
| Respiratory Failure | 476/12777 (3.7) | 523/13268 (3.9) | 0.95 (0.81, 1.10) | 0.66 | 26045 | 0.95 (0.77, 1.16) | 0.37 |
| AKI | 1173/12582 (9.3) | 1123/13020 (8.6) | 1.08 (0.98, 1.20) | 0.2 | 25602 | 1.08 (0.95, 1.23) | 0.05 |
| 30-day mortality | 374/13282 (2.8) | 360/13790 (2.6) | 1.08 (0.90, 1.30) | 0.66 | 27072 | 1.08 (0.85, 1.37) | 0.31 |
| Antibiotic redosing | 2048/2171 (94.3) | 2088/2219 (94.1) | 1.00 (0.98, 1.02) | 0.92 | 4390 | 1.00 (0.98, 1.03) | 0.74 |
| Normothermia | 8702/11605 (75.0) | 8880/11889 (74.7) | 1.00 (0.98, 1.03) | 0.92 | 23494 | 1.00 (0.98, 1.03) | 0.62 |
| Fraction time hypotensive | 0.03 (0.07) | 0.04 (0.07) | -0.00 (-0.00, 0.00) | 0.89 | 26640 | -0.00 (-0.00, 0.00) | 0.38 |
| Fraction time low PIP | 0.94 (0.18) | 0.94 (0.18) | 0.00 (-0.01, 0.01) | 0.92 | 20424 | 0.00 (-0.01, 0.01) | 0.57 |
| Hyperglycemia | 689/11574 (6.0) | 676/11937 (5.7) | 1.05 (0.91, 1.21) | 0.89 | 23511 | 1.05 (0.88, 1.26) | 0.34 |
| Anesthetic delivery | 9891/10043 (98.5) | 10158/10294 (98.7) | 1.00 (0.99, 1.00) | 0.87 | 20337 | 1.00 (0.99, 1.00) | 0.25 |
| Efficient gas flow > 90% | 6661/8805 (75.7) | 6768/9029 (75.0) | 1.01 (0.98, 1.03) | 0.89 | 17834 | 1.01 (0.98, 1.04) | 0.31 |

**Sub Table 5: Subgroup analysis: ASA >= 3.** All other factors as in Table 2.

| Outcome | Intervention | Usual care | GEE coef (95% CI) | p value | N | GEE coef (99.5% CI) | uncorrected p |
|---|---|---|---|---|---|---|---|
| Delirium | 1053/3016 (34.9) | 1116/3198 (34.9) | 1.00 (0.92, 1.09) | >0.99 | 6214 | 1.00 (0.89, 1.12) | 0.99 |
| Respiratory Failure | 898/17556 (5.1) | 964/18428 (5.2) | 0.98 (0.87, 1.10) | 0.94 | 35984 | 0.98 (0.84, 1.13) | 0.62 |
| AKI | 1611/16710 (9.6) | 1754/17535 (10.0) | 0.96 (0.89, 1.05) | 0.7 | 34245 | 0.96 (0.87, 1.07) | 0.27 |
| 30-day mortality | 513/18287 (2.8) | 541/19213 (2.8) | 1.00 (0.85, 1.16) | >0.99 | 37500 | 1.00 (0.82, 1.22) | 0.95 |
| Antibiotic redosing | 3046/3233 (94.2) | 3155/3335 (94.6) | 1.00 (0.98, 1.01) | 0.89 | 6568 | 1.00 (0.98, 1.02) | 0.49 |
| Normothermia | 12686/15951 (79.5) | 12980/16485 (78.7) | 1.01 (0.99, 1.03) | 0.49 | 32436 | 1.01 (0.99, 1.03) | 0.09 |
| Fraction time hypotensive | 0.04 (0.08) | 0.04 (0.08) | -0.00 (-0.00, 0.00) | 0.53 | 37453 | -0.00 (-0.00, 0.00) | 0.12 |
| Fraction time low PIP | 0.90 (0.23) | 0.91 (0.22) | -0.00 (-0.01, 0.01) | 0.89 | 28848 | -0.00 (-0.01, 0.01) | 0.51 |
| Hyperglycemia | 1121/16446 (6.8) | 1111/17274 (6.4) | 1.06 (0.95, 1.18) | 0.57 | 33720 | 1.06 (0.92, 1.22) | 0.16 |
| Anesthetic delivery | 13859/14073 (98.5) | 14439/14638 (98.6) | 1.00 (0.99, 1.00) | 0.7 | 28711 | 1.00 (0.99, 1.00) | 0.26 |
| Efficient gas flow > 90% | 8877/12447 (71.3) | 9220/12981 (71.0) | 1.00 (0.98, 1.03) | 0.89 | 25428 | 1.00 (0.98, 1.03) | 0.63 |

**Sup Table 6: Subgroup analysis: overlap >= 50% of case.** All other factors as in Table 2.

| Outcome | Intervention | Usual care | GEE coef (95% CI) | p value | N | GEE coef (99.5% CI) | uncorrected p |
|---|---|---|---|---|---|---|---|
| Delirium | 996/3193 (31.2) | 1075/3409 (31.5) | 0.99 (0.90, 1.08) | 0.95 | 6602 | 0.99 (0.88, 1.11) | 0.77 |
| Respiratory Failure | 842/30523 (2.8) | 918/31519 (2.9) | 0.95 (0.84, 1.07) | 0.66 | 62042 | 0.95 (0.81, 1.10) | 0.25 |
| AKI | 1941/29735 (6.5) | 2079/30714 (6.8) | 0.96 (0.89, 1.04) | 0.66 | 60449 | 0.96 (0.87, 1.07) | 0.24 |
| 30-day mortality | 515/31551 (1.6) | 533/32660 (1.6) | 1.00 (0.86, 1.17) | >0.99 | 64211 | 1.00 (0.82, 1.22) | >0.99 |
| Antibiotic redosing | 4342/4565 (95.1) | 4399/4623 (95.2) | 1.00 (0.99, 1.01) | 0.99 | 9188 | 1.00 (0.98, 1.02) | 0.93 |
| Normothermia | 21630/27749 (77.9) | 21937/28299 (77.5) | 1.01 (0.99, 1.02) | 0.68 | 56048 | 1.01 (0.99, 1.02) | 0.25 |
| Fraction time hypotensive | 0.03 (0.08) | 0.04 (0.08) | -0.00 (-0.00, 0.00) | 0.62 | 63362 | -0.00 (-0.00, 0.00) | 0.17 |
| Fraction time low PIP | 0.92 (0.21) | 0.92 (0.21) | -0.00 (-0.01, 0.00) | 0.99 | 48955 | -0.00 (-0.01, 0.01) | 0.77 |
| Hyperglycemia | 1441/25224 (5.7) | 1365/26181 (5.2) | 1.10 (0.99, 1.21) | 0.09 | 51405 | 1.10 (0.97, 1.24) | 0.01 |
| Anesthetic delivery | 23879/24142 (98.9) | 24399/24625 (99.1) | 1.00 (1.00, 1.00) | 0.31 | 48767 | 1.00 (1.00, 1.00) | 0.06 |
| Efficient gas flow > 90% | 15970/21319 (74.9) | 16377/21829 (75.0) | 1.00 (0.98, 1.02) | 0.99 | 43148 | 1.00 (0.98, 1.02) | 0.80 |

**Sub Table 7: Subgroup analysis: top quartile of baseline mortality risk.** Mortality risk defined in supplemental methods: an xgboost model using common words in the surgical procedure. All other factors as in Table 2.

| Outcome | Intervention | Usual care | GEE coef (95% CI) | p value | N | GEE coef (99.5% CI) | uncorrected p |
|---|---|---|---|---|---|---|---|
| Delirium | 759/1979 (38.4) | 772/2053 (37.6) | 1.02 (0.92, 1.13) | 0.88 | 4032 | 1.02 (0.90, 1.16) | 0.63 |
| Respiratory Failure | 667/6056 (11.0) | 723/6295 (11.5) | 0.96 (0.84, 1.09) | 0.85 | 12351 | 0.96 (0.81, 1.13) | 0.41 |
| AKI | 792/6614 (12.0) | 821/6889 (11.9) | 1.00 (0.89, 1.13) | 0.93 | 13503 | 1.00 (0.86, 1.17) | 0.92 |
| 30-day mortality | 300/6870 (4.4) | 291/7166 (4.1) | 1.08 (0.88, 1.32) | 0.85 | 14036 | 1.08 (0.83, 1.40) | 0.37 |
| Antibiotic redosing | 1526/1642 (92.9) | 1565/1678 (93.3) | 1.00 (0.97, 1.02) | >0.99 | 3320 | 1.00 (0.97, 1.03) | 0.71 |
| Normothermia | 4928/6290 (78.3) | 5135/6526 (78.7) | 1.00 (0.97, 1.02) | >0.99 | 12816 | 1.00 (0.96, 1.03) | 0.65 |
| Fraction time hypotensive | 0.05 (0.09) | 0.06 (0.09) | -0.00 (-0.01, 0.00) | 0.25 | 13605 | -0.00 (-0.01, 0.00) | 0.03 |
| Fraction time low PIP | 0.93 (0.19) | 0.93 (0.19) | 0.00 (-0.01, 0.01) | >0.99 | 12163 | 0.00 (-0.01, 0.01) | 0.65 |
| Hyperglycemia | 412/6140 (6.7) | 426/6388 (6.7) | 1.01 (0.84, 1.20) | >0.99 | 12528 | 1.01 (0.80, 1.26) | 0.93 |
| Anesthetic delivery | 5815/5914 (98.3) | 6106/6207 (98.4) | 1.00 (0.99, 1.01) | >0.99 | 12121 | 1.00 (0.99, 1.01) | 0.84 |
| Efficient gas flow > 90% | 3061/5342 (57.3) | 3258/5648 (57.7) | 0.99 (0.95, 1.04) | >0.99 | 10990 | 0.99 (0.94, 1.05) | 0.70 |

**Sub Table 8: Exploratory analysis: interaction with surgical service**

| Outcome | F | p value | P uncorrected |
|---|---|---|---|
| Delirium | 0.58 | 0.84 | 0.85 |
| Respiratory Failure | 1.26 | 0.48 | 0.24 |
| AKI | 1.48 | 0.38 | 0.13 |
| 30-day mortality | 1.3 | 0.48 | 0.22 |
| Antibiotic redosing | 0.95 | 0.97 | 0.49 |
| Normothermia | 1.27 | 0.84 | 0.24 |
| Fraction time hypotensive | 0.94 | 0.97 | 0.50 |
| Fraction time low PIP | 0.43 | 0.97 | 0.94 |
| Hyperglycemia | 0.82 | 0.97 | 0.62 |
| Anesthetic delivery | 0.91 | 0.97 | 0.53 |
| Efficient gas flow > 90% | 1.05 | 0.95 | 0.40 |

**Sup Table 9: Sensitivity analysis: include subsequent cases on patients.** All other factors as in Table 2.

| Outcome | Intervention | Usual care | GEE coef (95% CI) | p value | N | GEE coef (99.5% CI) | uncorrected p |
|---|---|---|---|---|---|---|---|
| Delirium | 1483/4615 (32.1) | 1525/4777 (31.9) | 1.01 (0.93, 1.09) | >0.99 | 9392 | 1.01 (0.91, 1.11) | 0.83 |
| Respiratory Failure | 1273/36964 (3.4) | 1310/38288 (3.4) | 1.01 (0.91, 1.11) | >0.99 | 75252 | 1.01 (0.89, 1.14) | 0.87 |
| AKI | 2709/36700 (7.4) | 2811/37933 (7.4) | 1.00 (0.93, 1.06) | >0.99 | 74633 | 1.00 (0.91, 1.09) | 0.88 |
| 30-day mortality | 872/39098 (2.2) | 893/40462 (2.2) | 1.01 (0.90, 1.14) | >0.99 | 79560 | 1.01 (0.87, 1.18) | 0.83 |
| Antibiotic redosing | 5532/5829 (94.9) | 5617/5901 (95.2) | 1.00 (0.99, 1.01) | 0.85 | 11730 | 1.00 (0.98, 1.01) | 0.48 |
| Normothermia | 27038/34156 (79.2) | 27461/34895 (78.7) | 1.01 (0.99, 1.02) | 0.52 | 69051 | 1.01 (0.99, 1.02) | 0.16 |
| Fraction time hypotensive | 0.04 (0.08) | 0.04 (0.08) | -0.00 (-0.00, 0.00) | 0.46 | 78159 | -0.00 (-0.00, 0.00) | 0.10 |
| Fraction time low PIP | 0.92 (0.22) | 0.92 (0.21) | -0.00 (-0.01, 0.00) | 0.85 | 60879 | -0.00 (-0.01, 0.01) | 0.56 |
| Hyperglycemia | 1846/31826 (5.8) | 1788/33046 (5.4) | 1.07 (0.98, 1.17) | 0.20 | 64872 | 1.07 (0.96, 1.20) | 0.03 |
| Anesthetic delivery | 29599/29966 (98.8) | 30339/30669 (98.9) | 1.00 (1.00, 1.00) | 0.46 | 60635 | 1.00 (1.00, 1.00) | 0.10 |
| Efficient gas flow > 90% | 19594/26429 (74.1) | 20144/27147 (74.2) | 1.00 (0.98, 1.01) | 0.87 | 53576 | 1.00 (0.98, 1.02) | 0.88 |

**Sup Table 10: Exploratory stratification by "emergency" status**. ASA physical status with "E" modifier. This analysis was unplanned and requested by a reviewer.

| Outcome | Intervention | Usual Care | GEE coef (95% CI) | P uncorrected |
|---|---|---|---|---|
| **Emergency** | | | | |
| Delirium | 115/226 (50.9) | 115/214 (53.7) | 0.95 (0.76, 1.19) | 0.55 |
| Respiratory Failure | 102/458 (22.3) | 97/437 (22.2) | 1.00 (0.74, 1.37) | 0.98 |
| AKI | 85/586 (14.5) | 77/565 (13.6) | 1.06 (0.74, 1.53) | 0.67 |
| 30-day mortality | 81/620 (13.1) | 67/606 (11.1) | 1.18 (0.80, 1.74) | 0.28 |
| Antibiotic redosing | 85/90 (94.4) | 89/94 (94.7) | 1.00 (0.91, 1.10) | 0.94 |
| Normothermia | 448/535 (83.7) | 416/508 (81.9) | 1.02 (0.95, 1.10) | 0.43 |
| Fraction time hypotensive | 0.06 (0.12) | 0.05 (0.09) | 0.01 (-0.00, 0.03) | 0.04 |
| Fraction time low PIP | 0.93 (0.20) | 0.91 (0.22) | 0.01 (-0.02, 0.05) | 0.26 |
| Hyperglycemia | 50/516 (9.7) | 47/514 (9.1) | 1.06 (0.63, 1.79) | 0.77 |
| Anesthetic delivery | 490/505 (97.0) | 463/483 (95.9) | 1.01 (0.98, 1.05) | 0.32 |
| Efficient gas flow > 90% | 290/432 (67.1) | 274/414 (66.2) | 1.01 (0.89, 1.16) | 0.77 |
| **Non-emergency** | | | | |
| Delirium | 1149/3647 (31.5) | 1183/3830 (30.9) | 1.02 (0.94, 1.11) | 0.57 |
| Respiratory Failure | 969/33538 (2.9) | 1033/34799 (3.0) | 0.97 (0.87, 1.09) | 0.54 |
| AKI | 2231/32665 (6.8) | 2355/33876 (7.0) | 0.98 (0.91, 1.06) | 0.54 |
| 30-day mortality | 549/34682 (1.6) | 582/36019 (1.6) | 0.98 (0.84, 1.14) | 0.73 |
| Antibiotic redosing | 5055/5312 (95.2) | 5166/5415 (95.4) | 1.00 (0.99, 1.01) | 0.56 |
| Normothermia | 24034/30612 (78.5) | 24490/31369 (78.1) | 1.01 (0.99, 1.02) | 0.21 |
| Fraction time hypotensive | 0.03 (0.07) | 0.04 (0.08) | -0.00 (-0.00, 0.00) | 0.08 |
| Fraction time low PIP | 0.92 (0.22) | 0.92 (0.21) | -0.00 (-0.01, 0.00) | 0.64 |
| Hyperglycemia | 1619/27953 (5.8) | 1569/29085 (5.4) | 1.07 (0.98, 1.18) | 0.04 |
| Anesthetic delivery | 26477/26773 (98.9) | 27225/27493 (99.0) | 1.00 (1.00, 1.00) | 0.14 |
| Efficient gas flow > 90% | 17678/23586 (75.0) | 18272/24336 (75.1) | 1.00 (0.98, 1.01) | 0.77 |

**Sup Table 11: Sensitivity analysis: un-clustered analysis.** All other factors as in Table 2.

| Outcome | Intervention | Usual care | GEE coef (95% CI) | p value | N | GEE coef (99.5% CI) | uncorrected p |
|---|---|---|---|---|---|---|---|
| Delirium | 1483/4615 (32.1) | 1525/4777 (31.9) | 1.01 (0.92, 1.10) | >0.99 | 9392 | 1.01 (0.89, 1.13) | 0.86 |
| Respiratory Failure | 1273/36964 (3.4) | 1310/38288 (3.4) | 1.01 (0.91, 1.11) | >0.99 | 75252 | 1.01 (0.89, 1.14) | 0.87 |
| AKI | 2709/36700 (7.4) | 2811/37933 (7.4) | 1.00 (0.93, 1.07) | >0.99 | 74633 | 1.00 (0.91, 1.09) | 0.88 |
| 30-day mortality | 872/39098 (2.2) | 893/40462 (2.2) | 1.01 (0.90, 1.14) | >0.99 | 79560 | 1.01 (0.87, 1.18) | 0.83 |
| Antibiotic redosing | 5532/5829 (94.9) | 5617/5901 (95.2) | 1.00 (0.95, 1.05) | 0.88 | 11730 | 1.00 (0.94, 1.06) | 0.88 |
| Normothermia | 27038/34156 (79.2) | 27461/34895 (78.7) | 1.01 (0.98, 1.03) | 0.70 | 69051 | 1.01 (0.98, 1.04) | 0.49 |
| Fraction time hypotensive | 0.04 (0.08) | 0.04 (0.08) | -0.00 (-0.00, 0.00) | 0.18 | 78159 | -0.00 (-0.00, 0.00) | 0.07 |
| Fraction time low PIP | 0.92 (0.22) | 0.92 (0.21) | -0.00 (-0.01, 0.00) | 0.70 | 60879 | -0.00 (-0.01, 0.00) | 0.53 |
| Hyperglycemia | 1846/31826 (5.8) | 1788/33046 (5.4) | 1.07 (0.98, 1.17) | 0.14 | 64872 | 1.07 (0.96, 1.20) | 0.04 |
| Anesthetic delivery | 29599/29966 (98.8) | 30339/30669 (98.9) | 1.00 (0.98, 1.02) | 0.86 | 60635 | 1.00 (0.97, 1.03) | 0.85 |
| Efficient gas flow > 90% | 19594/26429 (74.1) | 20144/27147 (74.2) | 1.00 (0.97, 1.03) | 0.88 | 53576 | 1.00 (0.97, 1.03) | 0.93 |

**Sup Table 12: Main Analysis before correction for multiple testing.** All other factors as in Table 2.

| Outcome | Intervention | Control | GEE coef (95% CI) | N | GEE coef (99.5% CI) | uncorrected p |
|---|---|---|---|---|---|---|
| Delirium | 1264/3873 (32.6) | 1298/4044 (32.1) | 1.02 (0.95, 1.08) | 7917 | 1.02 (0.93, 1.11) | 0.61 |
| Respiratory Failure | 1071/33996 (3.2) | 1130/35236 (3.2) | 0.98 (0.90, 1.07) | 69232 | 0.98 (0.87, 1.11) | 0.68 |
| AKI | 2316/33251 (7.0) | 2432/34441 (7.1) | 0.99 (0.93, 1.04) | 67692 | 0.99 (0.91, 1.07) | 0.63 |
| 30-day mortality | 630/35302 (1.8) | 649/36625 (1.8) | 1.01 (0.90, 1.13) | 71927 | 1.01 (0.86, 1.18) | 0.9 |
| Antibiotic redosing | 5140/5402 (95.1) | 5255/5509 (95.4) | 1.00 (0.99, 1.01) | 10911 | 1.00 (0.99, 1.01) | 0.56 |
| Normothermia | 24482/31147 (78.6) | 24906/31877 (78.1) | 1.01 (1.00, 1.01) | 63024 | 1.01 (0.99, 1.02) | 0.18 |
| Fraction time hypotensive | 0.03 (0.08) | 0.04 (0.08) | -0.00 (-0.00, 0.00) | 70913 | -0.00 (-0.00, 0.00) | 0.18 |
| Fraction time low PIP | 0.92 (0.22) | 0.92 (0.21) | -0.00 (-0.00, 0.00) | 55479 | -0.00 (-0.01, 0.00) | 0.74 |
| Hyperglycemia | 1669/28469 (5.9) | 1616/29599 (5.5) | 1.07 (1.00, 1.15) | 58068 | 1.07 (0.98, 1.18) | 0.04 |
| Anesthetic delivery | 26967/27278 (98.9) | 27688/27976 (99.0) | 1.00 (1.00, 1.00) | 55254 | 1.00 (1.00, 1.00) | 0.22 |
| Efficient gas flow > 90% | 17968/24018 (74.8) | 18546/24750 (74.9) | 1.00 (0.99, 1.01) | 48768 | 1.00 (0.98, 1.01) | 0.78 |

**Sup Table 13: Exploratory per-protocol analysis**. See supplemental methods “Per-protocol” exploratory subgroup analysis page S.6.

| Outcome | Intervention | Usual care | GEE coef (95% CI) | p value | N | GEE coef (99.5% CI) | uncorrected p |
|---|---|---|---|---|---|---|---|
| Delirium | 162/561 (28.9) | 175/549 (31.9) | 0.91 (0.72, 1.14) | 0.45 | 1110 | 0.91 (0.68, 1.22) | 0.28 |
| Respiratory Failure | 117/5623 (2.1) | 138/5599 (2.5) | 0.84 (0.62, 1.15) | 0.42 | 11222 | 0.84 (0.57, 1.26) | 0.17 |
| AKI | 449/5420 (8.3) | 449/5452 (8.2) | 1.01 (0.86, 1.18) | 0.93 | 10872 | 1.01 (0.82, 1.24) | 0.93 |
| 30-day mortality | 89/5775 (1.5) | 61/5775 (1.1) | 1.46 (0.96, 2.21) | 0.10 | 11550 | 1.46 (0.85, 2.49) | 0.02 |
| Antibiotic redosing | 1143/1194 (95.7) | 1060/1101 (96.3) | 0.99 (0.97, 1.02) | 0.96 | 2295 | 0.99 (0.97, 1.02) | 0.50 |
| Normothermia | 4614/5584 (82.6) | 4606/5539 (83.2) | 0.99 (0.97, 1.02) | 0.96 | 11123 | 0.99 (0.96, 1.02) | 0.47 |
| Fraction time hypotensive | 0.02 (0.05) | 0.02 (0.05) | 0.00 (-0.00, 0.00) | >0.99 | 11399 | 0.00 (-0.00, 0.00) | 0.92 |
| Fraction time low PIP | 0.90 (0.23) | 0.91 (0.23) | -0.01 (-0.02, 0.01) | 0.89 | 10521 | -0.01 (-0.02, 0.01) | 0.24 |
| Hyperglycemia | 331/4850 (6.8) | 322/4774 (6.7) | 1.01 (0.83, 1.24) | >0.99 | 9624 | 1.01 (0.78, 1.31) | 0.88 |
| Anesthetic delivery | 5170/5262 (98.3) | 5123/5194 (98.6) | 1.00 (0.99, 1.00) | 0.72 | 10456 | 1.00 (0.99, 1.00) | 0.12 |
| Efficient gas flow > 90% | 3736/4569 (81.8) | 3699/4477 (82.6) | 0.99 (0.96, 1.02) | 0.92 | 9046 | 0.99 (0.96, 1.02) | 0.31 |

**Adherence to ACT recommendations.** To characterize adherence to ACT recommendations, we randomly selected 50 pre-emptive case reviews and 50 alert responses which indicated that they had been communicated to the OR clinicians. We manually extracted any actionable recommendations and reviewed the intraoperative record for evidence that those recommendations had been followed. The times that these actions were taken was compared to the timestamp of the form indicating that it had been communicated to the OR to exclude OR actions that preceded rather than followed the ACT intervention. Reviews were performed by author TB and EK. Rates of any actionable requests (requests before similar strategies had been taken) and rates of adherence to those requests were tabulated and 95% confidence intervals calculated.

**Preemptive Case Review Results**

| message | Action-able Request | Action Done | OR Action Detail | Action Pre-request | Comment |
|---|---|---|---|---|---|
| Target MAP >= 70; Redose antibiotics at drug-specific intervals; Consider multimodal analgesia therapies; Use of the following fresh gas flow rates for volatile anesthesia to reduce cost and pollution | yes | yes | MAP >70 majority of case; multimodal pain regimen | no | |
| Check glucose at least Q 2 hour; Patient on chronic steroid treatment, consider stress dose if persistent hypotension | yes | yes | glucose monitred q2h | no | |
| Consider proactive warming due to a higher patient-specific risk of hypothermia; Patient with history of PONV, consider multimodal antiemetic prophylaxis; patient is receiving 4mg of dexamethasone q6h, she got the first dose this morning 5:40 am. Consider giving the next dose around noon. | yes | yes | proactive warming therapies initated; ondansetron administered; dexamethasone administered | no | |
| Phenylephrine as a first line vasopressor for this patient; Tranfusing to keep Hgb above 7.0 g/dL; Check glucose at least Q 2 hour; Consider proactive warming due to a higher patient-specific risk of hypothermia; assessing for full reversal and/or minimize non-depolarizing NMBD use due to patient risk from | yes | yes | phenylephrine initated; warming therapies | yes | |

| | | | | |
|---|---|---|---|---|
| residual blockade; Consider subcutaneous heparin for DVT prophylaxis; Redose antibiotics at drug-specific intervals; Consider multimodal analgesia therapies | | | | |
| Check glucose at least Q 2 hour; recommend entering OSA risk orders for the post-op period due to known OSA or high OSA risk status; Initiate temperature reading if not already done; Mild to mod global LV systolic dysfunction with EF 41% | yes | yes | temperature monitred; glucose measured | no |
| Consider administering ondansetron intra-operatively; Our machine learning predicion algorithms show a risk of postop aki 4.7%. Other postop complications are low at <1.0% | yes | yes | ondansetron administered | no |
| recommend maintaining higher MAP during periods of significant head up positioning; Check glucose at least Q 2 hour; Patient with Biotronik ICD/PM with magnet placement disabling anti-tachycardia function. Recent TTE suggestive of RV-pacing, recommend careful monitoring of arterial line for bradycardia during periods of electrocautery | yes | yes | measured glucose | no |
| Initiate insulin infusion if glucose > 180 mg/dL; assess and document TOF during case; assessing for full reversal and/or minimize non-depolarizing NMBD use; Consider EEG/BIS monitoring to assist in awareness prevention; Redose antibiotics at drug-specific intervals | yes | yes | insulin infusion; BIS monitoring | yes |
| Patient is at very high risk of postop respiratory failure (specifically, inability to extubate at case end and postop reintubation). If you get close to case end and respiratory mechanics are still suboptimal, consider calling the SICU triage fellow early so they can save you a bed just in case you can't extubate; I would | yes | yes | no toradol administered; DC to floor with OSA precautions, oximetry, and telemetry | no |

| | | | | | |
|---|---|---|---|---|---|
| recommend avoiding toradol in this patient, because its most significant risk is GIB. In patients with poor renal clearance and liver disease, the risk of GIB is likely higher than in the general population. | | | | | |
| Check glucose at least Q 2 hour; Consider administering ondansetron intra-operatively; consider reducing the use of opioids by using regional or local anesthesia or non-opioid analgesics | yes | yes | glucose measured q2h; ondansetron administered | no | |
| Consider using dexamethasone and ondansetron intra-operatively; consider reducing the use of opioids by using regional or local anesthesia or non-opioid analgesics | yes | yes | dexamethasone and ondansetron administered | yes | |
| recommend entering OSA risk orders; Check glucose at least Q 2 hour; Consider using dexamethasone and ondansetron intra-operatively; consider administering at least 10ml/kg of fluid to decrease baseline risk of PONV; consider reducing the use of opioids by using regional or local anesthesia or non-opioid analgesics | yes | yes | glucose measured q2h; dexamethasone and ondansetron adminstered | no | |
| recommend avoiding excessive volume administration due to patient-specific risk of volume overload; Consider multimodal analgesia therapies | no | n/a | n/a | n/a | case was short scope procedure and messege sent at end of case, no time to implement recomendations. |
| We recommend a target MAP >= 65; Consider proactive warming due to a higher patient-specific risk of hypothermia; Consider multimodal analgesia therapies | yes | yes | fluid warmer, forced air wamer, blankets; multimodal analgesics initiated | yes | |
| Check glucose at least Q 1 hour; consider multimodal anti-emetic prophylaxis | yes | yes | glucose monitred q2h; ondansetron administered | no | |

| | | | | |
|---|---|---|---|---|
| We recommend a target MAP >= 70; Check glucose at least Q 1 hour; Initiate insulin infusion if glucose > 180 mg/dL | yes | yes | MAP >70 majority of case; q1h glucose measures; insulin initiated | no |
| recommend avoiding excessive volume administration due to patient-specific risk of volume overload; Due to a perceived risk for problematic LV or RV systolic dysfunction, this patient may require inotropic support; recommend entering OSA risk orders | yes | yes | no excess volume administered | yes |
| Consider administering ondansetron intra-operatively; onsider reducing the use of opioids by using regional or local anesthesia or non-opioid analgesics | yes | no | n/a | n/a |
| We recommend a target MAP >= 65; Remember to assess and document TOF during case; recommend assessing for full reversal and/or minimize non-depolarizing NMBD use; Redose antibiotics at drug-specific intervals; Consider multimodal analgesia therapies; recommend using no anti-emetic or one anti-emetic agent intra-operatively | yes | yes | elevated MAP maintained; multimodal analgesics; TOF monitoring throughout case | no |
| We recommend phenylephrine as a first line vasopressor for this patient; Consider proactive warming due to a higher patient-specific risk of hypothermia; Consider EEG/BIS monitoring to assist in awareness prevention; Redose antibiotics at drug-specific intervals | yes | no | n/a | n/a |
| We recommend a target MAP >= 75; recommend assessing for full reversal and/or minimize non-depolarizing NMBD use; Redose antibiotics at drug-specific intervals; recommend using no anti-emetic or one anti-emetic agent intra-operatively | yes | yes | elevated MAP maintained; appropriate antiemetic | yes |
| We recommend a target MAP >= 75; Check glucose at least Q 2 hour; Consider EEG/BIS monitoring to assist | yes | yes | glucose measured q2h | no |

| | | | | |
|---|---|---|---|---|
| in awareness prevention; Redose antibiotics at drug-specific intervals;Consider multimodal analgesia therapies; recommend using no anti-emetic or one anti-emetic agent intra-operatively | | | | |
| recommend entering OSA risk orders for the post-op period; Remember to assess and document TOF during case | yes | yes | TOF assessed | no |
| We recommend a target MAP >= 75; recommend entering OSA risk orders for the post-op period due to known OSA; Consider multimodal analgesia therapies | yes | yes | elevated MAP maintained | yes |
| We recommend a target MAP >= 70; recommend avoiding excessive volume administration due to patient-specific risk of volume overload; we recommend considering transferring this patient to a higher level of care than normal after surgery such as an OU or ICU; recommend assessing for full reversal and/or minimize non-depolarizing NMBD use; Redose antibiotics at drug-specific intervals; Consider multimodal analgesia therapies | yes | yes | elevated MAP maintained; discharged to floor with telemetry; TOF prior to reversal | no |
| Consider multimodal antiemetic prophylaxis; receiving 4mg of dexamethasone every 6 hours. Next dose is at 11 am | yes | yes | multimodal antiemetic; dexamethasone time of administration as suggested | no |
| Consider multimodal analgesia therapies; Patient with A1C of 8.0 and no preoperative glucose. Recommend checking BG as soon as possible; Although unlikely to happen, it may be prudent to place a magnet over the CRT-D to disable the anti-tachycardia function, in the event of monopolar electrocautery. This will not disrupt the pacing (resynchronization) function. | yes | no | n/a | n/a |

| | | | | | |
|---|---|---|---|---|---|
| n/a | n/a | n/a | n/a | n/a | No recommendations listed in review file |
| n/a | n/a | n/a | n/a | n/a | No recommendations listed in review file |
| We recommend a target MAP >= 65; ecommend the use of the following fresh gas flow rates for volatile anesthesia to reduce cost and pollution | yes | yes | elvated MAP maintained | no | |
| Sent UpToDate link regarding NMBA use; Consider stress dose steroids if hypotensive; Could consider remifentanil to spare neuromuscular blocking drugs while connected to the robot; Drugs that might exacerbate MG: aminoglycosides, macrolides, fluoroquinolones, CCBs, BBs, gabapentin, diuretics, magnesium | yes | yes | remifentanil used | yes | |
| We recommend tranfusing to keep Hgb above 7.0 g/dL.; Consider checking hemoglobin during the case due to anticipated anemia risk; Remember to assess and document TOF during case; Consider subcutaneous heparin for DVT prophylaxis; Redose antibiotics at drug-specific intervals; Consider multimodal analgesia therapies | yes | yes | TOF assessed | yes | |
| Consider proactive warming due to a higher patient-specific risk of hypothermia | yes | yes | forced air wamer | yes | |
| Consider checking hemoglobin during the case due to anticipated anemia risk; Check glucose at least Q 2 hour; recommend entering OSA risk orders for the post-op period; Consider multimodal analgesia therapies; Dexamethasone has been administered. Consider administering ondansetron intra-operatively | yes | yes | multimodal analgesics administered | no | |

| | | | | |
|---|---|---|---|---|
| Due to a perceived risk for problematic LV or RV systolic dysfunction, this patient may require inotropic support; recommend norepinephrine as a first line vasopressor for this patient; Equipment for emergency defibrillation should be made available while defibrillator is programmed off; recommend considering the use of continued mechanical ventilation in the early post-op period; Initiate insulin infusion if glucose > 180 mg/dL and initiate Q 1 hour glucose checks at that time; Consider EEG/BIS monitoring to assist in awareness prevention; Redose antibiotics at drug-specific intervals | yes | yes | defibrillator programmed off; norepinephrine as first line | yes |
| We recommend considering the use of continued mechanical ventilation in the early post-op period; Consider proactive warming due to a higher patient-specific risk of hypothermia; Remember to assess and document TOF during case; Consider subcutaneous heparin for DVT prophylaxis; Redose antibiotics at drug-specific intervals; Consider multimodal analgesia therapies | yes | yes | TOF assessed; forced air warmer | yes |
| We recommend maintaining higher MAP during periods of significant head up positioning; recommend using at least two anti-emetic agents | yes | yes | elevated MAP; multimodal antiemetic | no |
| We recommend administering volume as needed due to high likelihood of patient hypovolemia; We recommend tranfusing to keep Hgb above 7.0 g/dL; Initiate insulin infusion if glucose > 180 mg/dL and initiate Q 1 hour glucose checks at that time; Consider proactive warming due to a higher patient-specific risk of hypothermia; Consider multimodal analgesia | yes | yes | volume as neeeded; PRBC transfusion; multi-modal analgesics | yes |

| | | | | | |
|---|---|---|---|---|---|
| therapies; recommend using at least two anti-emetic agents | | | | | |
| n/a | n/a | n/a | n/a | n/a | No recommendations listed in review file |
| We recommend a target MAP >= 85; We recommend maintaining higher MAP during periods of significant head up positioning; We recommend administering volume as needed due to high likelihood of patient hypovolemia; We recommend phenylephrine as a first line vasopressor for this patient; Check glucose at least Q 2 hour; initiate insulin infusion if glucose > 180 mg/dL; Consider proactive warming due to a higher patient-specific risk of hypothermia; Consider multimodal analgesia therapies; recommend using a scopolamine patch along with at least two anti-emetics | yes | yes | glucose measured q2h; phenylephrine as first line | no | |
| We recommend a target MAP >= 70; recommend phenylephrine as a first line vasopressor; recommend tranfusing to keep Hgb above 7.0 g/dL; Consider proactive warming due to a higher patient-specific risk of hypothermia; Consider EEG/BIS monitoring to assist in awareness prevention; Consider multimodal analgesia therapies; recommend the use of the following fresh gas flow rates for volatile anesthesia to reduce cost and pollution | yes | yes | elevated MAP maintained; BIS monitoring; | yes | |
| recommend entering OSA risk orders for the post-op period; Check glucose at least Q 2 hour; consider reducing the use of opioids by using regional or local anesthesia or non-opioid analgesics | yes | no | n/a | n/a | |

| | | | | |
|---|---|---|---|---|
| Consider proactive warming due to a higher patient-specific risk of hypothermia; Consider EEG/BIS monitoring to assist in awareness prevention | yes | yes | forced air wamer | no |
| Consider proactive warming due to a higher patient-specific risk of hypothermia; Remember to assess and document TOF during case; Redose antibiotics at drug-specific intervals | yes | yes | forced air wamer; TOF assessed; antibiotic redosed | no |
| Check glucose at least Q 2 hour; Redose antibiotics at drug-specific intervals; monitor K trend and assess for EKG changes | yes | yes | glucose measured q2h | yes |
| Consider proactive warming due to a higher patient-specific risk of hypothermia; Avoid hypercarbia, acidosis and hypoxemia to prevent increase in PVR | yes | no | n/a | n/a |
| We recommend a target MAP >= 75; Check glucose at least Q 2 hour | yes | yes | elevated MAP maintained | yes |
| Check glucose at least Q 2 hour; Initiate temperature measurement when feasible | yes | yes | temperature monitred; glucose measured | no |
| With moderate AS and CAD disease management, consider higher maps greater that 70-75, with a slow HR; Depending on time since paralytic given, and how the case progresses, could consider checking train of four at the end of the case to help facilitate quick extubation in ICU | yes | yes | elvated MAP with low HR maintained; TOF assessed | yes |
| Consider using dexamethasone and ondansetron intra-operatively | yes | yes | dexamethasone and ondansetron administered | no |

Alert response Results

| **Alert message** | **Action-able Request** | **Request List** | **Action Done** | **OR Action Detail** | **Action Pre-request** | **Comment** |
|---|---|---|---|---|---|---|

| | | | | | | |
|---|---|---|---|---|---|---|
| Cumulative time for MAP < 60 = 12 minutes. | yes | CA3 Resident requested to go to OR to assist; Place a-line | yes | resident went to room; a-line placed | no | a-line placed prior to resident arriving to room |
| Glucose = 203. | yes | Initiate Insulin infusion; measure glucose | yes | initiated insulin | no | |
| Diabetes. No glucose recorded in last 2.9 hours. | yes | measure glucose | yes | glucose measured | no | |
| Low Temperature = 34.9 °C | yes | Increase warming therapies | yes | started forced air warming | no | |
| Diabetes. No glucose recorded in last 4.6 hours. | yes | measure glucose | yes | glucose measured | no | |
| Diabetes. No glucose recorded in last 182 minutes. | yes | measure glucose | yes | glucose measured | yes | |
| Diabetes. No glucose recorded in last 191 minutes. | yes | measure glucose | yes | glucose measured | no | |
| Diabetes. No glucose recorded in last 170 minutes. | yes | measure glucose | no | n/a | n/a | |
| No BP measured [15m] | yes | measure blood pressure | yes | blood pressure measured; monitor set to repeat bp measures | no | |
| Hypotension: MAP = 57 | yes | reduce anesthetic dose | yes | expired sevo reduced from ~2.3 to ~1.5 | no | |
| Antibiotic (ampicillin-sulbactam (UNASYN) injection) overdue by 9 minutes. | yes | redose antibiotic | no | n/a | n/a | |

| | | | | | | |
|---|---|---|---|---|---|---|
| Diabetes. No glucose recorded in last 180 minutes. | yes | measure glucose | yes | glucose measured | no | |
| High MAC = 1.7 | yes | reduce anesthetic dose | no | n/a | n/a | expired sevo maintained > ~2.0 for majority of case |
| No BP Measured [11m] | yes | measure blood pressure | yes | blood pressure measured | no | |
| Low Temperature = 34.7 °C | yes | Increase warming therapies | yes | forced air warmer | yes | |
| Diabetes. No glucose recorded in last 2.8 hours. | yes | measure glucose | yes | glucose measured | no | |
| Temperature not measured. | yes | measure temperature | yes | temperature monitoring initiated | yes | |
| Antibiotic ceFAZolin (ANCEF) injection (14:28) documented 4 minutes before surgical incision (14:32) outside the 60-5 minute window before incision. | yes | assess documentation for accuracy | no | n/a | n/a | |
| Tidal volume = 664 (13.9 cc/kilo of ideal body weight). Consider changing tidal volume to 300 to 375 (6-8 cc/kilo). | yes | reduce tidal volume | no | n/a | n/a | |
| Low SpO<sub>2</sub> = 78 | yes | requested attending POD leader to assist with low room SpO2 | yes | Attending went to room; SpO2 increased | no | |

| | | | | | | |
|---|---|---|---|---|---|---|
| Low Temperature = 34.9 °C | yes | Increase warming therapies | yes | forced air warmer | yes | |
| Low CO<sub>2</sub> = 19.6 | yes | possible PE, assistance to room | yes | OR conducted TEE at bedside | yes | Already concerned with possible PE prior to alert; Bedside TEE conducted |
| Diabetes. No glucose recorded in last 2 hours. | yes | measure glucose | yes | glucose measured | no | |
| Low Temperature = 33.4 °C | yes | Increase warming therapies | yes | warming therapies increased; temperature monitoring location changed | yes | |
| Temperature not measured. | yes | Initiate temperature monitoring | yes | temperature monitoring initiated | no | |
| Antibiotic overdue by 29 minutes. | yes | redose antibiotic | no | based on creatinine clearance, was redosing q4h | n/a | ACT request based on incorrect data |
| Diabetes. No glucose recorded in last 2.5 hours. | yes | measure glucose | yes | glucose measured | no | |
| Diabetes. No glucose recorded in last 169 minutes. | yes | measure glucose | yes | glucose measured | no | |
| Diabetes. No glucose recorded in last 156 minutes. | yes | measure glucose | no | n/a | n/a | measured glucose in PACU ~30 min after contact by ACT. |
| No glucose recorded | yes | measure glucose | yes | glucosed measured | yes | glucose measured ~1h before surgery and q2h from that point. |

| | | | | | | |
|---|---|---|---|---|---|---|
| Temperature not measured. | yes | Initiate temperature monitoring | yes | temperature monitoring initiated | no | |
| Glucose = 276. Consider starting insulin infusion 1.0 units / h | yes | measure glucose; initiate insulin infusion | yes | initiated insulin; glucose measured q1h | no | |
| Insulin infusion. Consider measuring glucose. | yes | measure glucose | yes | glucose measured q1h | no | had previously measured (initiated insulin use), did not re-check in q1h window |
| Diabetes. No glucose recorded in last 16.7 days. | yes | measure glucose | yes | glucose measured | yes | |
| FGF = 2.4 L/min. Wasting anesthetic vapor. Consider decreasing fresh gas flow. | yes | reduce fresh gas flow | no | n/a | n/a | |
| | yes | increase/maximize warming therapies | no | n/a | n/a | |
| Cumulative time for MAP < 60 = 18 minutes. | yes | administer volume; administer vasopressor | yes | phenylephrine administered | no | |
| Diabetes. No glucose recorded in last 2.5 hours. | yes | measure glucose | yes | glucose mesured | no | |
| Temperature not measured. | yes | Initiate temperature monitoring | yes | temperature monitoring initiated | no | |
| High Potassium = 6.2 | yes | review ekg; re-measure potassium; | yes | re-measured potassium; administered calcium chloride | no | |

| | | | | | | |
|---|---|---|---|---|---|---|
| | | administer stabilizing measures for hyperkalemia | | | | |
| Lung Compliance (dynamic) = 7.2 | no | n/a | n/a | n/a | n/a | patient in prone position |
| | yes | measure glucose | no | n/a | n/a | |
| Low Temperature = 33.7 °C | yes | assess monitoring for accuracy | no | n/a | n/a | |
| Glucose = 243. Consider starting insulin infusion 1.0 units / h | yes | initiate insulin infusion | yes | initiated insulin | yes | |
| Hct = 20.9. Consider transfusion | yes | transfuse prbc | yes | prbc transfused | no | |
| Cumulative time for MAP < 60 = 15 minutes. | yes | administer volume | no | n/a | n/a | case ended shortly after message sent |
| Reversal administered without TOF documentation. | yes | measure and document TOF | yes | TOF measured and documented | no | |
| Diabetes. No glucose recorded in last 162 minutes. | yes | measure glucose | no | n/a | n/a | |
| Temperature not measured. | yes | Initiate temperature monitoring | yes | temperature monitoring initiated | no | |
| Antibiotic (ceFAZolin (ANCEF) injection) overdue by 10 minutes. | yes | redose antibiotic | yes | antibiotic redosed | no | |

### CONSORT Harms 2022 integrated into CONSORT 2010 items checklist of information to include when reporting a randomised trial

| Section/Topic | Item No | Checklist item | Reported on page No |
|---|---|---|---|
| **Title and abstract** | | | |
| | 1a | Identification as a randomised trial in the title | 1 |
| | 1b | Structured summary of trial design, methods, results of outcomes of benefits and harms, and conclusions (for specific guidance see CONSORT for abstracts) | 3 |
| **Introduction** | | | |
| Background and objectives | 2a | Scientific background and explanation of rationale | 4 |
| | 2b | Specific objectives or hypotheses for outcomes benefits and harms | 4 |
| **Methods** | | | |
| Trial design | 3a | Description of trial design (such as parallel, factorial) including allocation ratio | 5 |
| | 3b | Important changes to methods after trial commencement (such as eligibility criteria), with reasons | NA – No methods changes |
| Participants | 4a | Eligibility criteria for participants | 5 |
| | 4b | Settings and locations where the data were collected | 4 |
| Interventions | 5 | The interventions for each group with sufficient details to allow replication, including how and when they were actually administered | 4-5 |
| Outcomes | 6a | Completely defined pre-specified primary and secondary outcome measures for both benefits and harms, including how and when they were assessed | 5 |
| | 6b | Any changes to trial outcomes after the trial commenced, with reasons | Statistical analysis plan |
| | 6c | Describe if and how non-prespecified outcomes of benefits and harms were identified, including any selection criteria, if applicable | NA – no harms were measured |
| Sample size | 7a | How sample size was determined | 6 |
| | 7b | When applicable, explanation of any interim analyses and stopping guidelines | None |

| Section/Topic | Item No | Checklist item | Reported on page No |
|---|---|---|---|
| Randomisation: | | | |
| Sequence generation | 8a | Method used to generate the random allocation sequence | 5 |
| | 8b | Type of randomisation; details of any restriction (such as blocking and block size) | 5 |
| Allocation concealment mechanism | 9 | Mechanism used to implement the random allocation sequence (such as sequentially numbered containers), describing any steps taken to conceal the sequence until interventions were assigned | 5 |
| Implementation | 10 | Who generated the random allocation sequence, who enrolled participants, and who assigned participants to interventions | 5 |
| Blinding | 11a | If done, who was blinded after assignment to interventions (e.g., participants, care providers, those assessing outcomes of benefits and harms) and how | 5 |
| | 11b | If relevant, description of the similarity of interventions | NA |
| Statistical methods | 12a | Statistical methods used to compare groups for primary and secondary outcomes of both benefits and harms | 6 |
| | 12b | Methods for additional analyses, such as subgroup analyses and adjusted analyses | Supplemental methods |
| **Results** | | | |
| Participant flow (a diagram is strongly recommended) | 13a | For each group, the numbers of participants who were randomly assigned, received intended treatment, and were analysed for outcomes of benefits and harms | Fig 1 |
| | 13b | For each group, losses and exclusions after randomisation, together with reasons | Fig 1 |
| Recruitment | 14a | Dates defining the periods of recruitment and follow-up for outcomes of benefits and harms | 5 |
| | 14b | Why the trial ended or was stopped | 7 |
| Baseline data | 15 | A table showing baseline demographic and clinical characteristics for each group | Table 1 |
| Numbers analysed | 16 | For each group, number of participants (denominator) included in each analysis and whether the analysis was by original assigned groups and if any exclusions were made | Table 2 |
| Outcomes and estimation | 17a | For each primary and secondary outcome of benefits and harms, results for each group, and the estimated effect size and its precision (such as 95% confidence interval) | Table 2 |
| | 17a2 | For outcomes omitted from the trial report (benefits and harms), provide rationale for not reporting and indicate where the data on omitted outcomes can be accessed | NA |
| | 17b | Presentation of both absolute and relative effect sizes is recommended, for outcomes of benefits and harms | 9 |
| | 17c | Report zero events if no harms were observed | No harms measured |
| Ancillary analyses | 18 | Results of any other analyses performed, including subgroup analyses and adjusted analyses, distinguishing pre-specified from exploratory | Supplement |
| Harms | 19 | All important harms or unintended effects in each group (for specific guidance see CONSORT for harms) | NA |

| Section/Topic | Item No | Checklist item | Reported on page No |
|---|---|---|---|
| **Discussion** | | | |
| Limitations | 20 | Trial limitations, addressing sources of potential bias related to the approach to collecting or reporting data on harms, imprecision, and, if relevant, multiplicity or selection of analyses | 13 |
| Generalisability | 21 | Generalisability (external validity, applicability) of the trial findings | 14 |
| Interpretation | 22 | Interpretation consistent with results, balancing benefits and harms, and considering other relevant evidence | 13 |
| **Other information** | | | |
| Registration | 23 | Registration number and name of trial registry | 5 |
| Protocol | 24 | Where the full trial protocol and other relevant documents can be accessed, including additional data on harms | 5 |
| Funding | 25 | Sources of funding and other support (such as supply of drugs), role of funders | 14 |

Note: Adapted from Schulz (2010) to integrate items of CONSORT Harms 2022 (Junqueira 2022) [https://creativecommons.org/licenses/by/2.0/]. CONSORT items 1b, 2b, 6a, 11a, 12a, 13a, 14a, 16a, 17a, 17b, 18, 20 and 24 of were modified to incorporate elements relevant to the reporting of harms. Two new items were added (item 6c and 17a2). Please see the CONSORT Harms 2022 statement for additional details (Junqueira 2022).

We strongly recommend reading the CONSORT 2010 statement (Schulz 2010) in conjunction with the CONSORT Harms 2022 statement (Junqueira 2022) for important clarifications on all the items. If relevant, we also recommend reading CONSORT extensions for cluster randomised trials, non-inferiority and equivalence trials, non-pharmacological treatments, adaptive designs, pilot and feasibility studies, multi arm trials, cross-over and pragmatic trials. Additional extensions are forthcoming: for those and for up-to-date references relevant to this checklist, see EQUATOR Network.

ACTFAST Author Group

| First Name | Last Name | Research Role | Degree | Clinician Role |
|---|---|---|---|---|
| Michael | Avidan | PI | MD | Attending |
| Joanna | Abraham | Investigator | PHD | N/A |
| Maureen | Arends | Support | | N/A |
| Kara | Battig | Clinician | BSN, RN | CRNA |
| Arbi | Ben Abdallah | Investigator | DES, PHD | N/A |
| Danielle | Benematti | Clinician | BS, BSN | CRNA |
| George | Benzinger | Clinician | MD, PHD | Attending |
| Mara | Bollini | Investigator | BA, BSN, MHA | N/A |
| Margaret | Bradley | Clinician | BSN | CRNA |
| Thaddeus | Budelier | Investigator | MD, MSF | N/A |
| Kathryn | Cass | Clinician | CRNA | CRNA |
| Yixin | Chen | Investigator | PHD | N/A |
| Daniel | Eddins | Clinician | BS, BSN, RN | CRNA |
| David | Eisenbath | Clinician | BSN, MSN | CRNA |
| Daniel | Emmert | Clinician | PHD, MD | Attending |
| Ellen | Fischbach | Support | BS, CCRP | N/A |
| Bradley | Fritz | Investigator | MD | Attending |
| Jason | Gillihan | Clinician | MD | Attending |
| Marie | Goez | Clinician | RN, DNP | CRNA |
| Shreya | Goswami | Coordinator | MBBS, DNB | N/A |
| Thomas | Graetz | Clinician | MD | Attending |
| Stephen | Gregory | Clinician | MD | Attending |
| Ryan | Guffey | Clinician | MD | Attending |
| Charles | Hantler | Clinician | MD | Attending |
| Peter | Haw | Clinician | BSN | CRNA |
| Daniel | Helsten | Clinician | MD | Attending |
| Bernadette | Henrichs | Clinician | PHD | CRNA |
| Erin | Herrera | Clinician | MSN | CRNA |
| Omokhaye | Higo | Clinician | MD | Attending |
| Robert | Hovis | Clinician | CRNA | CRNA |
| Gary | Hubbard | Clinician | MS | CRNA |
| Rocco | Hueneke | Clinician | MD | Attending |
| Ivan | Kangrga | Clinician | MD, PHD | Attending |

| | | | | |
|---|---|---|---|---|
| Thomas | Kannampallil | Investigator | PHD | N/A |
| Menelaos | Karanikolas | Clinician | MD, MPH | Attending |
| Christopher | King | Investigator | MD, PHD | Attending |
| Holly | Kirkpatrick | Clinician | BSN | CRNA |
| Justin | Knittel | Clinician | MD | Attending |
| Helga | Komen | Clinician | MD | Attending |
| Alexander | Kronzer | Support | BA | N/A |
| Anand | Lakshminarasimhachar | Clinician | MD, MD | Attending |
| Alyssa | McClellan | Clinician | BSN | CRNA |
| Sherry | McKinnon | Support | BS, MS, CCRP | N/A |
| Alicia | Meng | Support | BA | N/A |
| Alexander | Mohrmann | Clinician | BSN | CRNA |
| David | Monks | Clinician | MBChB FRCA MSc | Attending |
| Mary | Politi | Investigator | PHD | N/A |
| Debra | Pulley | Clinician | MD | Attending |
| Rashmi | Rathor | Clinician | MD | Attending |
| Andrea | Reidy | Clinician | MD | Attending |
| Cameron | Ritter | Clinician | BS, BSN, MS | CRNA |
| Elvira | Sayfutdinova | Clinician | BSN | SRNA |
| Craig | Schadler | Clinician | BSN | CRNA |
| Elizabeth | Schappe | Clinician | MS | CRNA |
| Tracey | Stevens | Clinician | MD | Attending |
| Marko | Todorovic | Clinician | MD | Attending |
| Brian | Torres | Clinician | MS, DNP, CRNA | CRNA |
| Bradley | Uding | Clinician | BS, BSN | CRNA |
| Brandon | Ufert | Clinician | RN | SRNA |
| Swarup | Varaday | Clinician | MBBS,FRCA(UK),FCARSI(IRELAND), MD | Attending |
| Troy | Wildes | Investigator | MD | Attending |
| William | Wise | Clinician | MSN | CRNA |
| Rachel | Wolfe | Investigator | PharmD | N/A |
| Sennaraj | Balasubramanian | Clinician | MD | Attending |
| Anuradha | Borle | Clinician | MD | Attending |
| Jamie | Brown-Shpigel | Clinician | MD | Attending |
| Jane | Exler | Clinician | BS | CRNA |
| Khatera | Najrabi | Clinician | BSN | CRNA |

| | | | | |
|---|---|---|---|---|
| David | Potter | Clinician | BSN, MBA | CRNA |
| Sarah | Sillery | Clinician | MSN | CRNA |
| Kate | Silver | Clinician | MSN | CRNA |
| Preet | Mohinder Singh | Clinician | MD | Attending |
| Melanie | Somercik | Clinician | MSN | CRNA |
| Bradley | Gerlach | Clinician | MSN | CRNA |
| Gregory | Miller | Clinician | MD | Resident |
| Adrienne | Nations | Clinician | BSN | CRNA |
| Lindsey | Schurter | Clinician | BSN | SRNA |
| Dean | Thorsen | Clinician | MD | Resident |
| Brian | Weber | Clinician | MD | Resident |
| Mark | Willingham | Clinician | BS, MD | Attending |
| Allison | Yu | Clinician | MD | Resident |
| Joseph | Balassi | Clinician | BSN | CRNA |
| Yunwei | Chen | Clinician | MD | Attending |
| Mathew | Gielow | Clinician | BSN | CRNA |
| Arianna | Montes de Oca | Coordinator | MD | N/A |
| Melissa | Milbrandt | Support | MBA, MHA | N/A |
| Mary | Zerlan | Clinician | DNP | CRNA |
| Courtney | Hardy | Clinician | MD | Attending |
| Divya | Mehta | Coordinator | MD | N/A |
| Ben | Snyders | Clinician | MSN | CRNA |
| Mohamed | Abdelhack | Support | PhD | N/A |
| Gregory | Feilner | Clinician | MSN,DNP | CRNA |
| Elyse | Kaestner | Clinician | MSN | CRNA |
| Molly | Barry | Clinician | MSN | CRNA |
| Vanya | Tumati | Clinician | MD | Resident |
| David | Carr | Clinician | MD | Resident |
| Andrew | Tran | Clinician | MD | Resident |
| Phillip | Taylor | Clinician | MD | Resident |
| Elizabeth | Schroeder | Clinician | MSN | CRNA |
| Jonathan | Jocum | Clinician | MD | Attending |
| Bruno | Maranhao | Clinician | MD, PHD | Resident |
| Hawa | Abubakar | Clinician | MD | Resident |
| Daniel | Fernandez | Clinician | MD | Resident |
| Danish | Jaffer | Clinician | MD | Resident |

| | | | | |
|---|---|---|---|---|
| Andrew | Pauszek | Clinician | MD | Resident |
| Ellen | Dekleva | Clinician | NA | SRNA |
| Catherine | Foster | Clinician | MD | Attending |
| Victoria | Koke | Clinician | MSN | CRNA |
| Megan | Taylor | Clinician | DNAP | CRNA |
| Benjamin | French | Clinician | MD | Resident |
| Benjamin | Fuller | Clinician | MD | Resident |
| Melissa | Milnor | Clinician | BSN | SRNA |
| Jessica | Sanford | Clinician | MD | Resident |
| Sandhya | Tripathi | Support | PhD | N/A |
| Fillmore | Gerardson Almiron | Clinician | NA | SRNA |
| Tamanna | Chang | Clinician | MD | Resident |
| Cynthia | Clark-Bumgarner | Clinician | NA | SRNA |
| Megan | Dewey | Clinician | MD | Resident |
| Ashley | Harrell | Clinician | BSN | SRNA |
| Claira | Sousa | Clinician | BSN | SRNA |
| Aldin | Turan | Clinician | BSN | SRNA |
| Jessica | Shah | Clinician | MSN | CRNA |
| Furqaan | Sadiq | Clinician | MD | Resident |
| Mingchun | Liu | Clinician | MD | Resident |
| Jing | Zhong | Clinician | MD | Resident |
| Jason | Han | Clinician | MD | Resident |
| Austin | Lohse | Clinician | MD | Resident |
| Jared | Wilmoth | Clinician | MD | Resident |
| James | Wirthlin | Clinician | MD | Resident |
| Mayank | Agarwal | Clinician | MD | Resident |
| Jooyoung | Maeng | Clinician | MD | Resident |
| Brennan | McMillan | Clinician | MD | Resident |
| David | Moquin | Clinician | MD | Resident |
| Shiv | Rawal | Clinician | MD | Resident |
| Alexander | Scott | Clinician | MD | Resident |
| Sara | Fisher | Support | MHA | N/A |
| Frank | Bougher | Clinician | BSN | SRNA |
| Suzanne | Frattini | Clinician | MHS | CRNA |
| Mallory | Light | Clinician | MSN | SRNA |
| Katherine | Mette | Clinician | BS | SRNA |

| | | | | |
|---|---|---|---|---|
| Benjamin | Thomas | Clinician | BSN | SRNA |
| India | Johnson | Clinician | DNAP | CRNA |
| Ramya | Baddigam | Clinician | MD | Resident |
| Emily | Davenport | Clinician | BSN | SRNA |
| Bethany | Geisler | Clinician | RN | SRNA |
| Robert | Graham | Clinician | RN | SRNA |
| Hunter | Niemeyer | Clinician | BSN | SRNA |
| Britni | Tharp | Clinician | BSN | SRNA |
| Christian | Guay | Clinician | MD | Resident |
| Micael | Bethel | Clinician | MD | Resident |
| Steven | Hedgecorth | Clinician | MD | Resident |
| Rida | Chaudhry | Clinician | MD | Resident |
| Krishna | Bhat | Clinician | MD | Resident |
| Michael | McLean | Clinician | BSN | SRNA |
| Jonathan | Ford | Clinician | MD | Resident |
| Joseph | Avery | Clinician | MD | Resident |
| Kiran | Kamath | Clinician | MD | Resident |
| Stephan | Diljak | Clinician | MD | Resident |
| Jiaxin | Huang | Clinician | MD | Resident |
| Zachary | Jergensen | Clinician | MD | Resident |
| Sheila | Sullivan | Clinician | MD | Resident |
| Bethany | Pennington | Investigator | PharmD | N/A |
| Kim | My Li | Clinician | MD | Resident |
| David | Rasche | Clinician | MD | Resident |
| Ryan | Pieterick | Clinician | MD | Resident |
| Ziyan | Song | Clinician | MD | Resident |
| Taylor | Crittenden | Clinician | BSN | SRNA |
| Paul | Kerby | Clinician | MBBS | Attending |
| Leander | Lee | Clinician | MD | Resident |
| Lingshu | Liu | Clinician | MD | Resident |
| Ruchik | Patel | Clinician | MD | Resident |
| Jonathan | Rich | Clinician | MD | Resident |
| Manjaap | Sidhu | Clinician | MD | Resident |
| Peter | Szatkowski | Clinician | MD | Resident |
| Brandon | White | Clinician | MD | Resident |
| Meghan | Woodham | Clinician | BSN | SRNA |

| | | | | |
|---|---|---|---|---|
| Lauren | Schoolfield | Clinician | BSN | SRNA |
| Benjamin | Stivers | Clinician | MD | Resident |
| Madeline | Wilson | Clinician | BSN | SRNA |
| Neil | Frydrych | Clinician | BSN | SRNA |
| Amanuel | Gebeyehu | Clinician | BSN | SRNA |
| Ottavia | Green | Clinician | MD | Resident |
| Matthew | Marten | Clinician | BSN,DNP | CRNA |
| Jennifer | Stephan | Clinician | BSN | SRNA |
| Paul | Winson | Clinician | BSN | SRNA |
| Mary | Kirsch | Clinician | DNAP | CRNA |
| Meghan | Marshall | Clinician | BSN | SRNA |
| Sean | Pottenger | Clinician | BSN | SRNA |
| Todd | Rackohn | Clinician | MD | Resident |
| Evangelos | Karanikolas | Coordinator | MD | N/A |
| Jessica | Morrow | Clinician | DNP | CRNA |
| Mark | Hanak | Clinician | MD | Resident |
| Lauren | Lagrimas | Clinician | MD | Resident |
| Grace | Huang | Support | BS | N/A |
| Miguel | Valdez | Coordinator | MD | N/A |
| Maria | Angelica Baquero | Clinician | NA | CRNA |
| Abby | Bisch | Clinician | NA | CRNA |
| Belma | Kulovac | Clinician | NA | CRNA |
| Monika | Martinek | Support | BS | N/A |
| Jonathan | Zoller | Clinician | MD | Attending |
| Leon | Du Toit | Clinician | MD | Attending |
| Anchal | Bansal | Clinician | MD | Resident |
| Sarah | Perez | Support | MSN | N/A |
| Micaela | Clark | Clinician | MD | Resident |
| Jordan | Nations | Clinician | BSN | SRNA |
| Christine | Johnson | Clinician | RN | SRNA |
| Kate | Pozzo | Clinician | CRNA | CRNA |
| Julia | Downey | Clinician | DNAP | CRNA |
| Jackie | Yu | Clinician | MD | Resident |
| Mohanapriya | Arumugam | Coordinator | MD | N/A |
| Natalie | Profumo | Clinician | DNP | CRNA |
| Chelsea | Curless | Clinician | CRNA | CRNA |

| | | | | |
|---|---|---|---|---|
| Eleanor | Bilhorn | Clinician | CRNA | CRNA |
| Lokesh | Sharma | Clinician | MD | Resident |
| Pratyush | Sontha | Support | BA | N/A |
| Laura | Cavallone | Clinician | MD | Attending |
| William | Varnum | Clinician | MSN | CRNA |
| Martha | Sabino | Clinician | BSN | SRNA |
| Muthuraj | Kanakaraj | Clinician | MD | Attending |
| Muhan | Zhang | Support | PhD | |
